# Application of multi-theory model(MTM)health behavior change: A scoping review

**DOI:** 10.1101/2025.01.27.25321021

**Authors:** Panpan Huai, Linghui Zhang, Bo Zhang, Yao Li, Bing Wu, Huimei Lv, Hui Yang

**Author notes:** Corresponding author at: No.56, Xinjian South Road, Yingze District, Taiyuan City, Shanxi Province, China. (HML). Panpan Huai and Linghui Zhang were contributed to the work equally and should be regarded as co-first authors. Huimei Lv was the first corresponding author.

## Abstract

**Background:** Since its proposal in 2015, MTM has received a lot of attention in health behavior change research both at home and abroad, but the model is still in the exploratory stage. Furthermore, the majority of current health promotion research focuses on a particular health habit, using a rather simple theoretical model. Learning from the evolution of MTM, the study of various behavioral changes is an area of health promotion research that requires careful cultivation. The purpose of this review is to discuss the application of MTM theory in health behavior change. By synthesizing relevant literature, we can improve the understanding of multi-theory model of health behavior change, make up for the shortcomings of existing studies, and provide suggestions for future studies.

**Methods:** We conducted a scoping review of the literature using the Joanna Briggs Institute (JBI) framework and followed PRISMA-ScR guidelines to report findings. Databases such as PubMed, Embase, The Cochrane Liberary, Web of Science, Ovid, CNKI, Wanfang, Vip and Sinomed. In addition to the traditional scoping review, we also evaluated the quality of the included literature.

**Results:** MTM is summarized by literature review, and the application status of MTM is reviewed. A total of 68 studies were included in this study, including 50 quantitative investigations, 12 quantitative interventions and 6 qualitative studies.

**Conclusion:** The field of application for MTM is extensive, and it demonstrates a relatively high level of prediction accuracy and intervention effectiveness. Consequently, it can be effectively utilized to advance health behavior promotion and health education initiatives.

## Introduction

The World Health Organization (WHO) and Elsevier’s Public Health Practice (2024.11) published a joint call to action calling for a more evidence-based and people-centred approach to health behaviour. Health behavior change is the process by which an individual gradually shifts from a behavior pattern or habit that is detrimental to physical and mental health to a behavior pattern or habit that is beneficial to health in order to prevent disease and maintain his or her health, including the behavior change manifested and the change in his or her psychological consciousness[1]. The empirical study demonstrates that an intervention based on the theory of health behavior change can effectively promote health behavior change[2, 3]. It has undergone four generations of change, and the focus of the first three generations of theory has changed from focusing on information therapy[4] to skill development and awareness building to evidence-based technology[3]. However, fourth-generation trends integrate multiple constructs from multiple theories for precise interventions, utilizing technology and focusing on specific behavior change[5]. Nowadays, the focus of the modern medical model has changed from the treatment of the disease itself to the management of health behavior, and the development and reform of the theory has been in line with this reform trend. Multi-theory model (MTM), as a fourth-generation theory, is a behavioral theoretical model developed in recent years and first proposed by Sharma in 2015. It splits health behavior change into two stages: initiation of health behavior change and sustenance (or continuation) of health behavior change[6].

Three main constructs influence this initiation of health behavior change. The first is the participatory dialogue derived from Freire’s (1970) model of adult education[7]. Participatory dialogues are bilateral conversations initiated by health educators about the pros and cons of health behavior change to assess subjects’ perceptions of the benefits and barriers associated with such behaviors. This two-way communication or mutual exploration enables patients to fully understand the benefits of changing their health behaviors and to viscerally identify with those benefits based on their current situation. The second construct is behavioral confidence, derived from Bandura’s (1986) self-efficacy and Ajzen’s (1991) perceived behavioral control[8]. Behavioral confidence can stem from internal sources or external influences, such as influential individuals or groups in a person’s life, and health educators. At the same time, behavioral confidence can be used as an indicator of an individual’s certainty about future involvement in changing their health behaviors. It is distinct from perceived behavioral control, which focuses on assessing obstacles associated with performing the intended behavior. The third and final construct for initiation of a behavior is the changes in the physical environment, which is derived from Bandura’s (1986) construct of the environment, Prochaska’s (1979) construct of environmental reevaluation, and environmental factors in Fishbein’s (2009) integrative model, among others[9, 10]. Changes in the physical environment require changes in the availability, accessibility, convenience, and readiness of resources in the physical environment associated with promoting desired behaviors. Behavioral change to maintain health includes long-term manifestations of behavioral change (such as lifelong participation in physical activity). In order to sustain behavior change, we need to channel our emotions into goals, constantly strive for change, and need the support of the social environment.

Three main constructs influence the sustenance of health behavior change. The first construct is derived from the self-motivation construct of emotional intelligence theory (Goleman, 1995)[11]. Emotional transformation is overcoming self-doubt, inertia, and impulsivity by focusing your feelings and emotions on healthy behavioral changes that lead you to your goals. Guide emotions to assist the change of healthy behaviors, so that patients’ emotions do not easily retreat, do not act rashly, and try to maintain a relaxed and happy state to change healthy behaviors. The second construct is derived from Freire’s (1970) adult education model’s praxis and is called practice for change[5]. The practice of behavior change emphasizes reflective behavior, including continuous and thoughtful consideration of behavior change, combined with continuous correction to eliminate ineffective strategies and address barriers. Such as recording the improvement of patients at various stages, so that patients can quantitatively see their own changes, encourage and support patients. The third and final construct is derived from constructs of the environment (Bandura, 1986), helping relationships (Prochaska, 1979), social support (House, 1981), and so on[12–14]. Social environment change refers to the establishment of social support in the environment. Changes in the social environment may be natural or man-made, and health educators can help in this process.

There are six steps in the analysis of a theory: identification of theoretical roots; Explore the meaning of the theory; Analyze the logical sufficiency of theory to determine the usefulness of theory; Clarify the degree of generalization and minimalism of the theory; Determine the testability of the theory. Previous studies of MTM theory mainly focused on the first three points of theoretical analysis, mainly on the theory itself. However, we want to conduct a comprehensive theoretical analysis by way of scope review: the practicability and help degree that MTM theory can provide, the scope that MTM theory can be extended, and whether MTM theory can be supported by empirical data[15]. In addition, as far as we know from the scientific literature we have searched, there are very few scoped reviews specifically focused on MTM. Therefore, we want to review the application research of MTM in relevant fields at home and abroad through a comprehensive review of this method, discuss the progress of MTM in the practice research of health behavior change, and have a more comprehensive and in-depth understanding of the MTM theory of health behavior change. Doing so can provide a fair reference for future empirical research in this field, lay a foundation for future optimization of the theory and identify future applied research needs, thereby promoting a better understanding and more flexible application of MTM.

Therefore, in this paper, we use the method of scope review (the first article) to discuss the research application of MTM theory in health behavior change. By synthesizing relevant literature, we can improve the understanding of multi-theoretical models of health behavior change, make up for the shortcomings of existing studies, and provide suggestions for future studies. The ultimate aim is to enhance comprehension of these components, enabling informed decision-making, adaptation, and development of effective strategies tailored to specific situations by decision makers, practitioners, and researchers. The sub-questions constituting the specific objectives of this study are as follows: What are the research groups and associated behavior types within the MTM theory of health behavior change? Which evaluation tools are employed for assessing the MTM theory of health behavior change? What research methods does the MTM theory utilize to evaluate changes in health behavior?

## Methods

### Study design

Scope review, also known as scoping review, is a method of synthesizing knowledge and identifying evidence based on the principles of evidence-based practice. It serves as the initial step in literature research. Conducting a scope review necessitates systematic methodological design and standardized reporting of research findings[16]. Simultaneously, it does not impose strict requirements on the type or quality of included information resources, thereby addressing issues that cannot be analyzed through systematic reviews and bridging gaps where evidence needs summarization but may not align with systematic review applicability. Furthermore, compared to traditional reviews, scope reviews offer more comprehensive and systematic retrieval processes. This study aims to analyze the target population and behavior types related to MTM theory of health behavior change by summarizing the evidence in all different design types of studies in this field, as well as the assessment tools and research methods used to evaluate the impact of MTM theory on health behavior change. Therefore, a scope review is selected for detailed examination. A systematic scoping review was conducted to examine studies that reported theories, models, or frameworks related to health systems integration, following the JBI guidance recommended by Cochrane[17]. The review process for scoping reviews, based on PRISMA-ScR[18] and Arksey and O’Malley’s five-stage approach[19], was employed. Additionally, Peters’ guidance methods[20] and Pollock et al.’s recommendations[15] for scope reviews were consulted during data extraction and analysis. We also registered the protocol with PROSPERO International Prospective Register of Systematic Reviews.

Systematically searched all relevant literature published in PubMed, Embase, The Cochrane Liberary, Web of Science, Ovid, China National Knowledge Infrastructure (CNKI), Wanfang, VIP Database for Chinese Technical Periodicals (VIP) and Sinomed (5 English databases and 4 Chinese databases) since their establishment. Two members of the research team developed a search strategy based on keywords, which was endorsed by a third research member, mainly using the following keywords for searching “Multi-Theory Model”, “MTM”, “multi-theory model”, Boolean logic operators and or for combining these words. Take Pubmed as an example, the search method is as follow:

(Multi-Theory Model[Title/Abstract]) OR (MTM[Title/Abstract]).

### Inclusion and Exclusion Criteria

In order to make the literature research more systematic and comprehensive, we adopt two frameworks for comprehensive research. The literature inclusion criteria were formulated based on the SPIDER (Sample, Phenomenon of Interest, Design, Evaluation, Study Type)[21] and PCC (Population, Concept, Context) frameworks[22] as outlined in Table 1. The SPIDER framework encompasses diverse population groups and focuses on the application of MTM theory for health behavior change. Methodological approaches include questionnaire adjustments, scale evaluations, interviews, and dialogues among others. Outcome indicators evaluated encompass participatory dialogue outcomes, behavioral confidence levels, changes in material environment and emotions experienced by individuals undergoing behavior change practices along with alterations in social environments. Both quantitative and qualitative research methodologies were employed. Within the PCC framework contextually applied to this study’s scope; population refers to all individuals involved while concept primarily describes MTM theory for health behavior change.

**Table 1.**
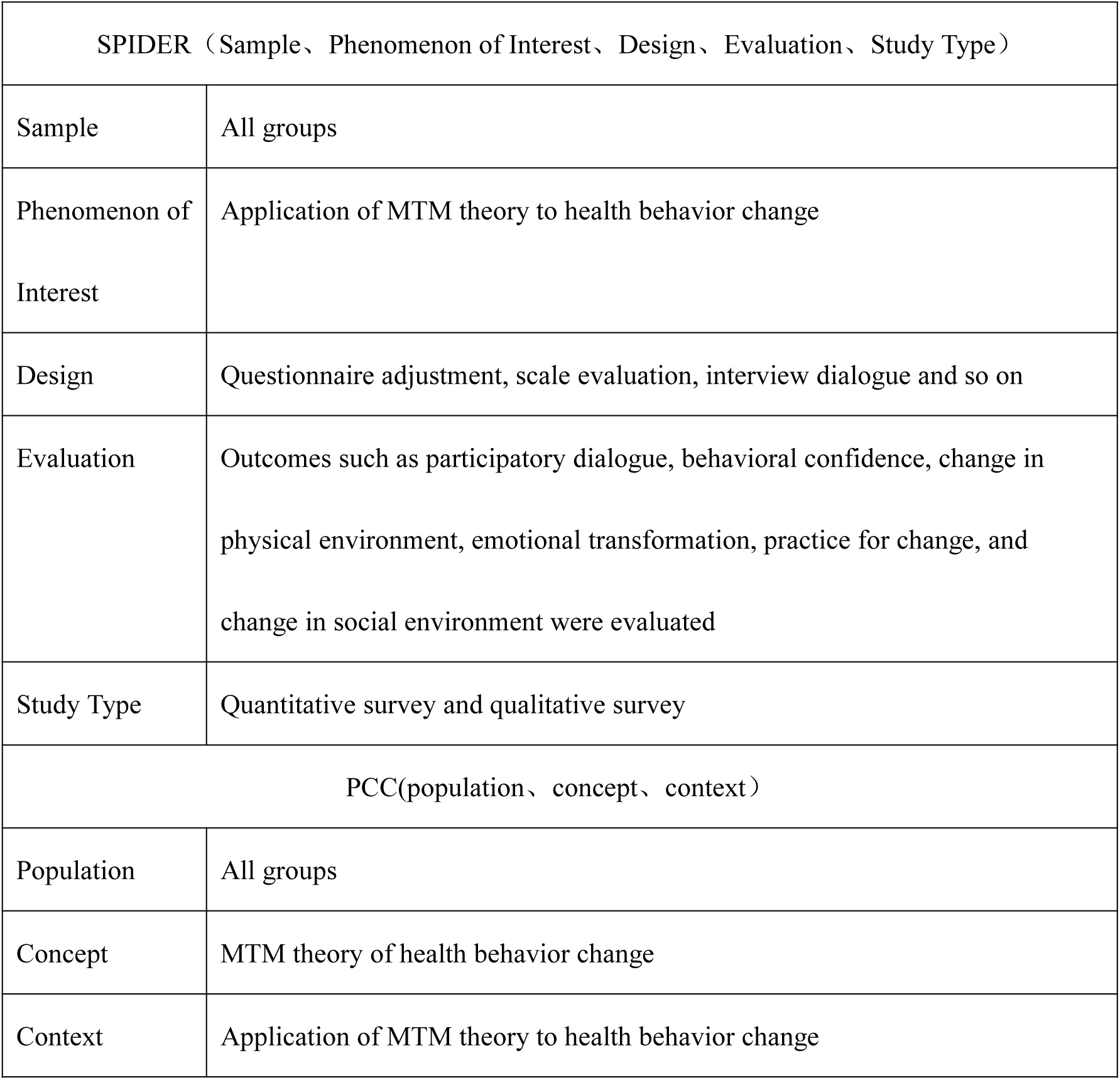
SPIDER framework and PCC framework.

The search excluded studies that did not adhere to the principles of MTM theory, those that were categorized as multi-theory rather than solely MTM theory and articles for which full-text access could not be obtained. At the same time, exclusion criteria for literature selection included:

(1) Incomplete data or unattainable outcome measures either directly or through conversion; (2) Duplicate publications reporting data from the same year, region, and population; (3) The article excludes grey literature; (4) Article search excluded relevant information from newspapers; (5) Non-chinese and non-English articles are excluded.

In order to enhance the scientific rigor of the study, we meticulously traced all references and relevant citations in the included literature to comprehensively gather research populations investigating the MTM theory of health behavior change, as well as pertinent literature on behavior types, assessment tools, and evaluation research methods.

### Study selection

In order to enhance the comprehensiveness and scientific rigor of the results, this scoping review encompassed all relevant literature published to date (Last retrieved on January 5, 2025.). Following a thorough familiarization with the existing body of literature, specific inclusion and exclusion criteria were established to ensure their applicability across all pertinent studies. The inclusion criteria comprised qualitative, quantitative, and interventional studies grounded in multiple theoretical models.

Two researchers independently extracted data from the included literature based on a pre-designed data extraction table, encompassing: (1) fundamental information of the literature such as title, author, impact factor, year of publication; (2) General study details such as study type, sample size, subjects, intervention and control measures; (3) Outcome indicators including measurement methods, tools, and time. Subsequently, retrieved documents were meticulously screened and assessed in strict adherence to predetermined inclusion and exclusion criteria. Any differences of opinion were resolved through consultation. In case of persistent disputes, a final decision was reached after discussion or consultation with a third party.

We imported the literature into NoteExpress literature management software for organization, reviewed the article titles, removed any duplicate references, further screened the literature that potentially met the requirements, and ultimately selected the final set of relevant articles.

## Data analysis

Based on the research questions, the literature content was organized, encompassing author and country information, publication year, journal, research objectives, population characteristics, time frame, study type, tools and methods employed for data collection and analysis techniques used. Furthermore, the research findings including main discoveries were extracted from two relevant articles by two independent researchers to create a preliminary data extraction table. After thorough discussion among the team members, necessary modifications were made to refine the contents of the table. Subsequently reviewed by the entire research team for accuracy and completeness, a final version of the data extraction table was established to extract all pertinent information presented in each paper. Additionally, an assessment of article quality or risk of bias was conducted.

The data were presented in the form of descriptive analysis, including: (1) Basic information: title, first author, corresponding author, country of origin, publication year, journal name and impact factor; (2) Research population characteristics: demographic details and target subjects; (3) Research methods employed: quantitative investigation research, quantitative intervention research, qualitative research etc.; (4) Research content addressed the following questions: 1) What are the populations studied and behavior types associated with MTM theory of health behavior change? 2) What evaluation tools are utilized for assessing health behavior change based on MTM theory? 3) What research methodologies are used to evaluate health behavior change according to MTM theory?

The methodological quality of all eligible studies was assessed using the JBI critical appraisal tools[15]. Two independent reviewers conducted the critical appraisal, and any discrepancies were resolved by a third reviewer following Lockwood et al.’s (2020) recommendation. The final appraisal outcome was reached through consensus. No studies were excluded.

## Results

### Study characteristics

A total of 13,759 articles were retrieved from various databases: PubMed (1,744), Embase (2,475), The Cochrane Library (207), Web of Science (2,100), Ovid (1,475), CNKI (3,366), Wan Fang (1,648), VIP (474) and Sinomed (266).

After removing duplicates and irrelevant literature based on title and abstract screening, a total of 8,681 articles remained for further analysis. After reading the full text and excluding studies that did not align with the research theory or main focus on types of health behaviors or had inaccessible full texts or conference abstracts only available, a final set of 68 relevant literatures were included in this study. The PRISMA screening process is illustrated in Fig.1.

**Fig 1.**
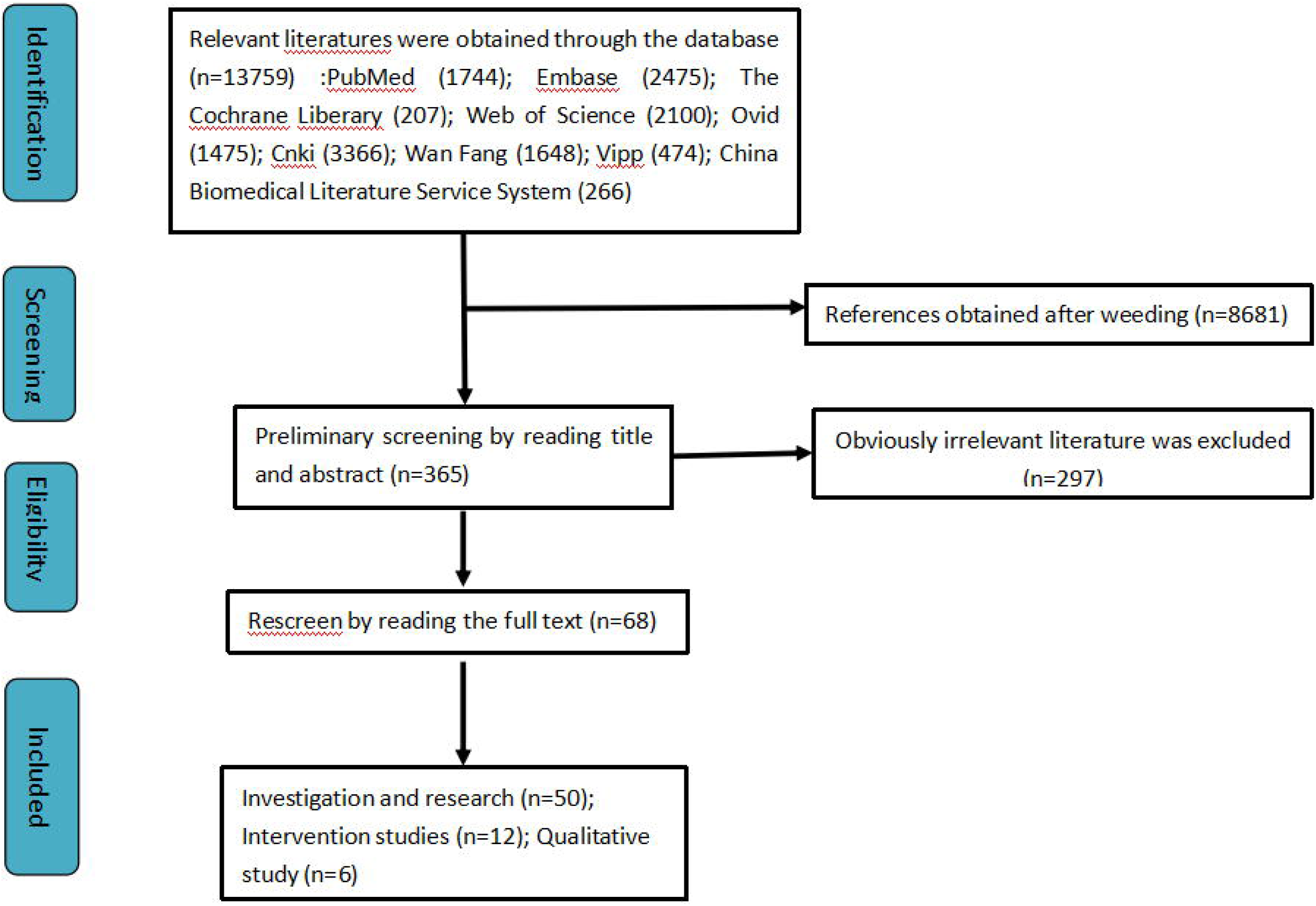
Flow chart. During the literature search, a total of 68 articles pertaining to this research question were identified. The majority of these articles were published in 2021 (n=16), followed by 15 in 2022, 10 in 2023, and 9 in 2024. Additionally, there were eight publications in 2020 and four in 2019. Three articles were retrieved from the year 2018, while one each was found for the years 2016 and 2017. Notably, no relevant publications were obtained for the year 2015. Fig.2 illustrates the annual publication trend over recent years. Manoj Sharma introduced the multi-theory model of health behavior change back in 2015 and has since conducted extensive research on it[5].

**Fig 2.**
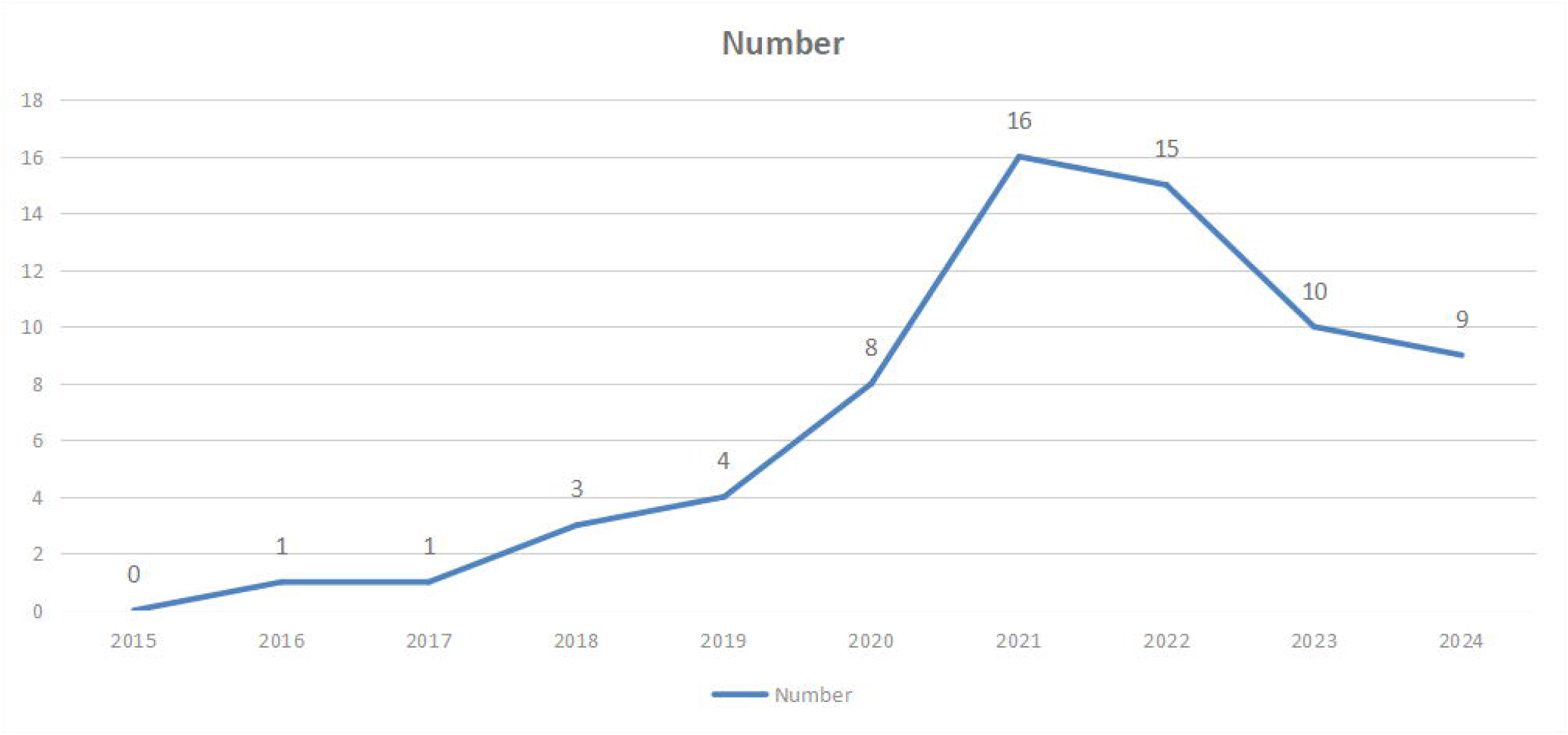
Article per year. Studies on the MTM theory of health behavior change have been conducted extensively, with the United States having the highest number of publications (n=40), followed by Iran (n=9), China (n=9), India (n=5), Ghana (n=2). Additionally, Germany and the United Kingdom have also contributed to this international trend in research content. Sharma, a renowned scholar specializing in health education and promotion in the United States, has dedicated several years to developing and evaluating interventions for health behavior change based on multiple theories. Therefore, the results of the study show that the United States has more research on this. The distribution of documents issued by each country is illustrated in Fig.3 below.

**Fig 3.**
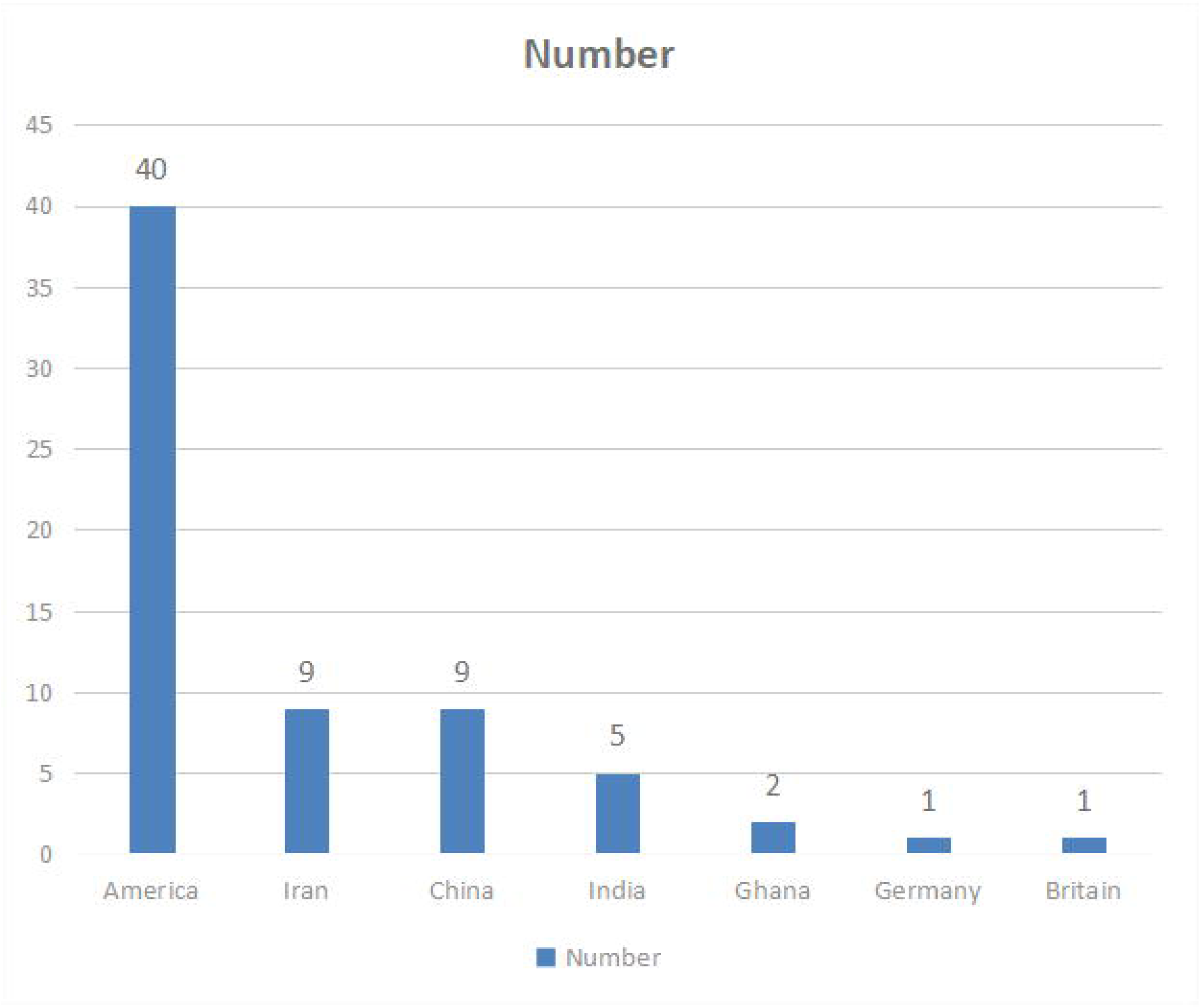
Number of publications by country. The study included a majority of usage survey categories (n=50) and quantitative interventions (n=12), while the remaining were qualitative in nature (n=6). The article types are presented in Fig.4.

**Fig 4.**
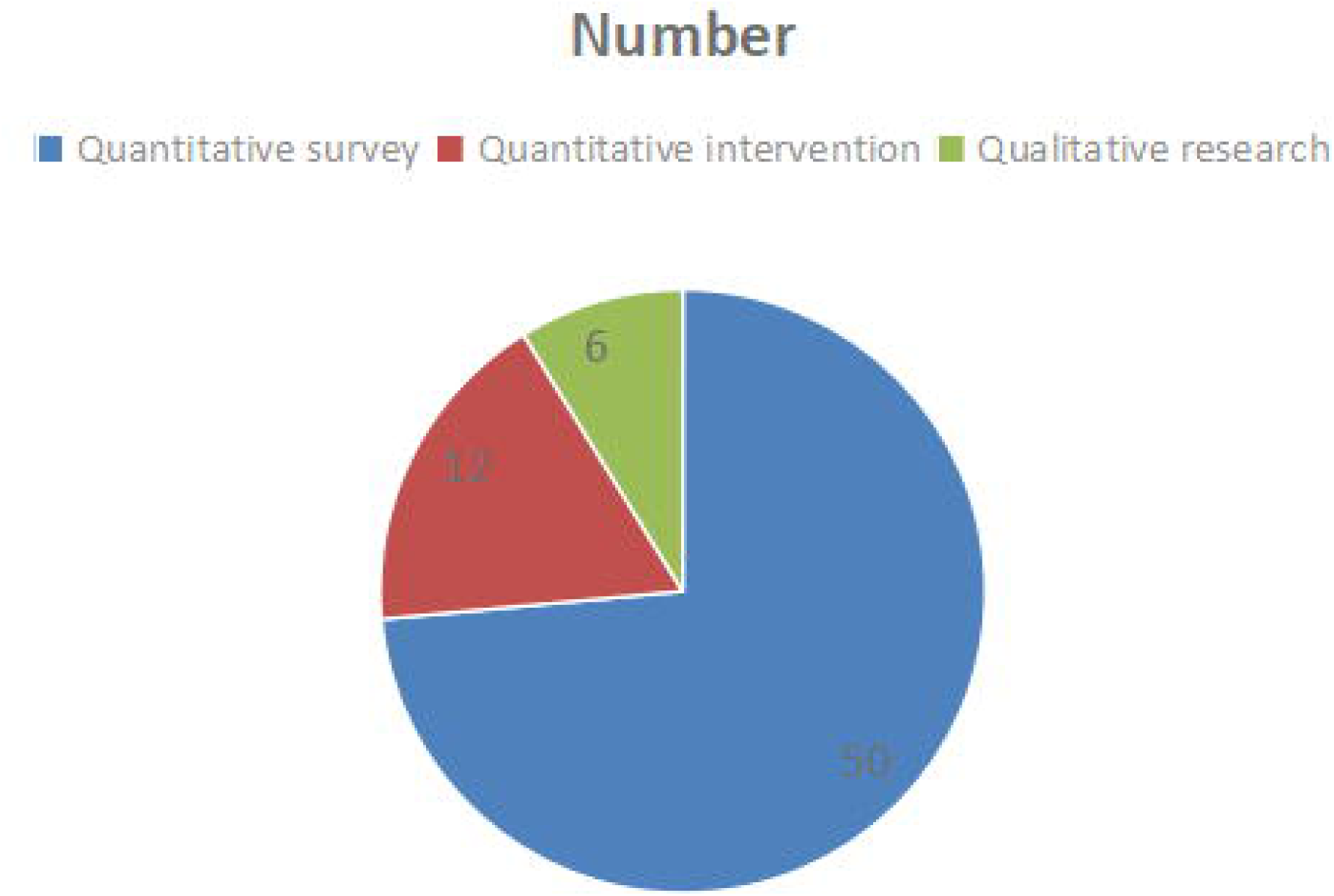
Article type. In the search of the articles, we found that the authors of the articles appeared most frequently: Manoj Sharma (n=27), Kavita Batra (n=15), Vinayak K. Nahar (n=5), Traci Hayes (n=4). These scholars have done more research on the MTM theory of health behavior change.

Details of the data are shown in **File 2. Characteristics of included studies.** and **File 3.**

**Information for intervention articles.** are enclosed in the Supporting Information.

### The populations studied and behavior types associated with MTM theory of health behavior change

The sample size of quantitative survey is large. The sample size of the quantitative survey ranges from 70 to 28,000 people, as shown in Fig.5 below. The number of samples in a quantitative intervention article is 400,150,125,100,94,80(intervention group is 40, regular group is 40),56,54(intervention group is 28, regular group is 26),48(intervention group is 25, regular group is 23) and so on. The sample size of the qualitative class was 79,34,16 and so on.

**Fig 5.**
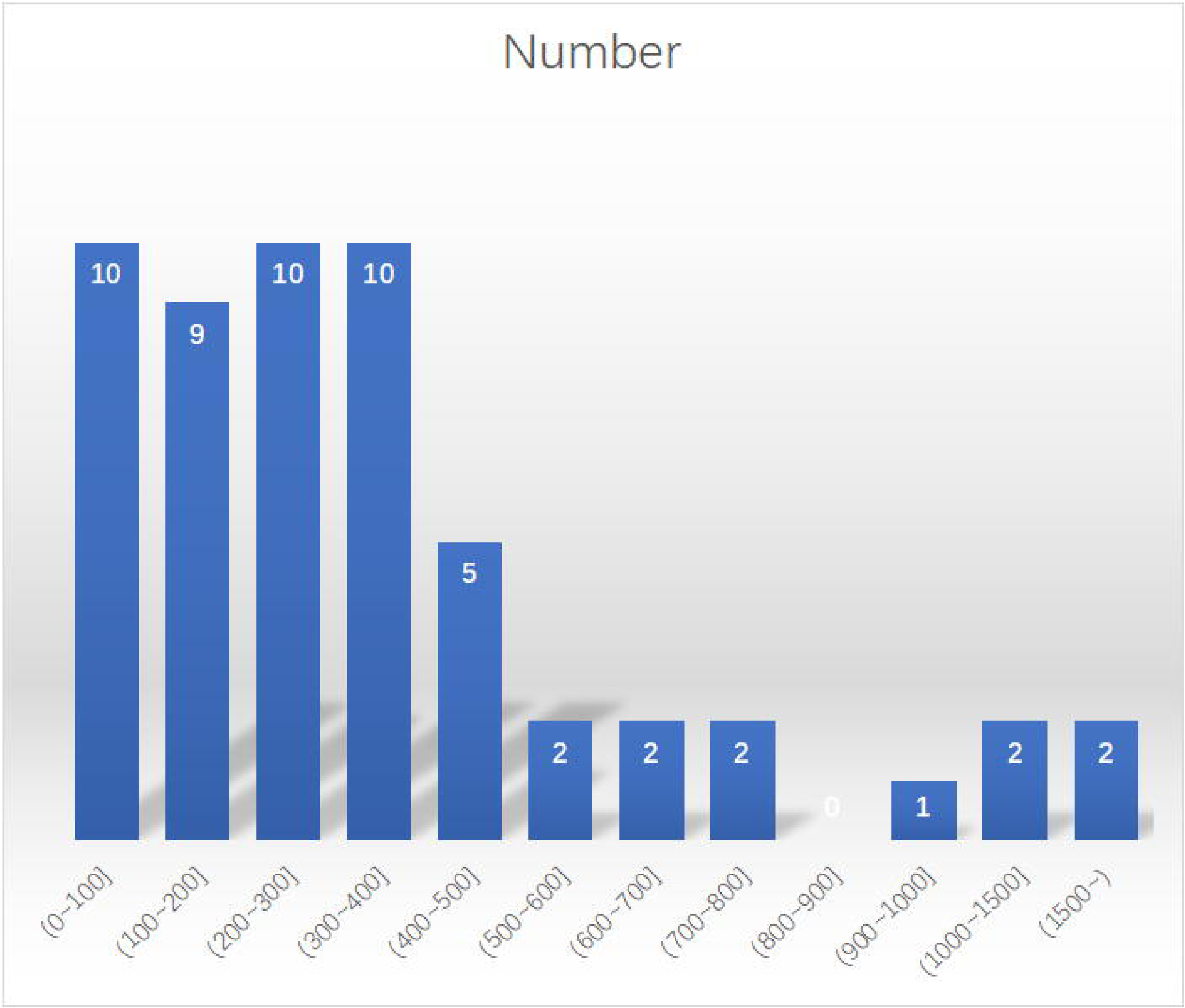
The number of samples in a quantitative intervention article. Of the 68 literature sources, most were studies that focused on promoting health behaviors. Among them, there are 25 basic health behaviors(n=25). They were 8 activities(n=8)[23–30], 7 diet(n=7) [31–37], 3 consumption(n=3)[38–40], 2 hygiene behaviors(n=2)[41, 42] and 1 sleep(n=1)[43, 44]. There were 16 health care behaviors, including 8 vaccine acceptance behaviors(n=8)[45–52] and 8 physical activity behaviors(n=8) [23–30]. There were 4 early warning behaviors:4 physical examination behavior(n=4)[53–56]. There were 3 avoid environmental hazards behaviors:3 behavior to improve quality of life(n=3)[57–59]. There were 6 behaviour to get rid of bad habits:6 smoking cessation behavior(n=6)[60–65]. A number of other behaviors were mentioned in the retrieved articles. However, the number of articles devoted to disease is limited, with only two publications identified (n=2). Studies include treatment for sleep apnea[66] and chemotherapy for gastrointestinal malignancies[67].

### An evaluation tool for MTM theory of health behavior change

In cross-sectional research, questionnaires serve as the primary research instruments. A total of 28 questionnaires were developed based on the theoretical framework of MTM, with each questionnaire comprising approximately 40 to 50 questions. Additionally, other research tools employed in this study included a self-designed MTM instrument, a survey tool devised by university students, an instrument adapted from a previous US study, a questionnaire focusing on children’s sleep habits, an open-ended questionnaire guide, an online survey software Qualtrics-generated questionnaire, a PA (physical activity) questionnaire and a validated questionnaire. Two studies utilized scales (n=2), while four studies employed hybrid research tools combining the MTM questionnaire with other questionnaires (n=4).

The main research methods employed in this study encompassed reliability and validity analysis, with a total of ten methods being evaluated, verified, analyzed, assessed, and investigated by expert groups (n=10). Additionally, four methods utilized Cronbach’s alpha coefficient (n=4), while the remaining approaches involved confirmatory factor analysis and alpha evaluation. Two of these techniques incorporated a five-point Likert scale (n=2). Furthermore, one article conducted a reading level assessment using the Flesh-Kincaid readability index (n=1). Lastly, one method applied the Integrated Marketing Communications (IMC) approach (n=1).

The intervention group generally received MTM-based interventions while the comparison group received conventional interventions. The intervention was designed in a rigorous way, through group discussion or expert validation. The type of intervention was a randomized controlled trial. The process of intervention is both online and offline. The results of the intervention proved to be effective. Through the retrieval of intervention articles, it is found that the effective intervention based on the MTM theory can effectively promote the change of health behavior of the intervention subjects compared with the comparison group.

### Research methods of MTM theory to evaluate health behavior change

The study variables were subjected to descriptive analysis, followed by modeling of the data using structural equation modeling method. The models employed included multiple linear regression model (n=17), stepwise multiple regression model (n=8), hierarchical regression model (n=7), structural equation modeling (n=4), Logit (OL model) (n=1), and path analysis model (n=1). Survey data was primarily analyzed using SPSS software package (n=41); however, SAS software was used in four papers, AMOS software in three papers, MSXQDA software and R Core Team in one paper each. Variable statistics encompassed univariate, bivariate, and multivariate methods.

Data testing methods comprised independent sample t-test (n=24), Pearson bivariate correlation test (n=15), Chi-square test (n=13), paired sample t physical examination test (n = 5), and Spearman correlation coefficient test (n = 2) across 24 papers. Other testing methods utilized included Logistic regression test, Shapiro-Wilk test, Fisher Pearson r-test, Welch t-test, Levene variance equality test, two-tailed F-test of econometric variables, Bonferron double comparison tests, Friedman tests, and Smirnov-Kolmogorov tests. The statistical techniques employed in this study involved calculating the frequency and percentage for categorical variables as well as computing the mean and standard deviation for measurement variables to describe the data.

### The result of literature quality evaluation

We evaluated the quality of all the 68 literatures included, and the specific results are as follows. In general, the quality of the included articles was above medium, including 22 articles of high quality, 45 articles of medium quality and 1 article of low quality.

## Discussion

The objective of this review is to examine the application of MTM theory in health behavior change. The findings demonstrate that the MTM serves as a theoretical framework for understanding the factors influencing the initiation and maintenance of health behavior change. Initiation of health behavior change involves transitioning from one behavior to another, encompassing one-time changes such as receiving a single dose vaccination or commencing regular physical exercise. At this stage, individuals must perceive the benefits of behavior change and possess behavioral confidence supported by environmental factors. Maintenance of healthy behavior change entails long-term adherence, such as lifelong engagement in physical exercise, necessitating emotional transformation into goals, continuous striving for improvement, and social support.

In the application research of MTM theory of health behavior change, logistic regression method is commonly employed for data analysis. However, the Analytic Network Process (ANP) can be utilized to analyze complex interdependencies and feedback loops among decision factors. ANP offers a more comprehensive and flexible approach by constructing a network model that captures dynamic relationships between decision elements[68]. It can elucidate the relationship between initiation and maintenance phases of the MTM theory of health behavior change as well as among its six dimensions: participatory dialogue, behavioral self-confidence, material environment change, emotional transformation, behavior change practice, and social environment change. This enhances precision in applying the MTM theory of health behavior change.

Alternatively, Latent Profile Analysis (LPA), a latent variable model used to identify latent or mixed categories based on continuous variables in a dataset, could be considered. When applied to studying MTM theory of health behavior change, LPA facilitates classification of subjects according to explicit variables using a small number of latent variables (class variables) to explain associations with continuous variables[69]. This aids in identifying or approximating potentially meaningful subject subgroups and contributes to better understanding sample heterogeneity.

Lifestyle intervention is an important part of chronic disease management. Studies in intervention articles have shown that effective interventions can promote changes in health behaviors. The intervention method can be carried out by means of online media, and PPT is used in two of the retrieved articles[28, 40]. With the development of Internet, comprehensive intervention scheme based on mobile medical technology has become a new trend of chronic disease management model research. Faced with the challenge of increasing complexity and comprehensiveness of intervention, a standard, detailed and comprehensive framework is conducive to deconstruction and analysis of intervention programs, so as to promote the improvement of the quality and effect of intervention[70]. For intervention methods, in the future, we advocate the use of the Internet platform and information means to achieve accurate intervention.

Health related behavior refers to any behavior of human individuals or groups related to disease prevention, health promotion, health maintenance and health restoration. According to the influence of the behavior on the health of the actor and others, it can be divided into two categories: promoting health behavior[71] and endangering health behavior[72]. Health-promoting behaviors are objectively beneficial to health, while harmful behaviors are objectively detrimental to health. Health promotion behaviors can be divided into five categories: (1) Basic health behaviors: refers to a series of basic health behaviors in daily life, such as active rest and sleep, reasonable nutrition and balanced diet[73]; (2) Health care behavior: refers to the reasonable and correct use of health care services to maintain their own health behavior, such as vaccination, regular physical examination, etc.[74]; (3) early warning behavior: prevention of accidents and correct disposal after accidents, including self-rescue and other rescue; (4) Avoid environmental hazards: Environmental hazards are broad, including the natural environment and psychosocial environment in which people live and work that are harmful to the body[75]. Such as leaving the polluted environment, taking measures to reduce environmental pollution, and actively dealing with those stressful life events that cause people’s psychological stress; (5) Get rid of bad habits: bad habits mainly refer to smoking, alcoholism, drug abuse and drug abuse[76]. There are two kinds of harmful behaviors to health: bad life style and habit and pathogenic behavior pattern.

According to the retrieved articles, it is found that the current research field of this theory is relatively shallow and the number of studies is relatively small. In fact, the MTM theory of health behavior change has been effectively applied to disease management, especially chronic diseases, and has produced significant benefits. The disease application of MTM health behavior change theory should be the focus of future research, and it has high research value for future research. Therefore, the MTM theory should be encouraged to be flexibly applied in the treatment of diseases in the future, giving full play to its advantages in precise intervention.

The research objects of the MTM health behavior change theory are mainly concentrated in the community, which is the limitation of our research object, and it is necessary to expand the research scope to make the research benefit more widely. At the same time, the application of MTM theory in health behavior change is an ongoing study, and attention should be paid to the dynamic nature of the study. In the study of behavior, it mainly involves more motor behavior, which needs comprehensive research, such as diet, mood, etc. At present, the multi-theory model of health behavior change is in the stage of development and improvement. Foreign scholars have optimized it through theoretical testing, theory-based intervention, qualitative research and other methods, and achieved certain results. However, the research on the multi-theory model of health behavior change in China is still in its infancy. In the future, there is an urgent need to explore assessment tools that are developed based on MTM framework under Chinese cultural background or Chinese and suitable for China’s national conditions, add Chinese elements to the development of multi-theory model of health behavior change, reasonably explain the intention status quo and influencing factors of health behavior change of Chinese people, and further enrich and expand the connotation and extension of MTM. In order to play a greater value in the field of health management[77].

### Strengths and limitations

This paper gives a comprehensive and systematic introduction to the MTM theory of health behavior change, which provides a valuable reference for future research in this field. The search strategy employed in this study adhered to JBI Methodology for Scoping Review, incorporating detailed steps to ensure inclusion of all relevant studies and conducting quality appraisal for each included study. All authors reviewed, discussed, and reached consensus on every step of the review process. However, due to language limitations, only English and Chinese articles were considered while important studies in other languages might have been overlooked, potentially impacting our findings negatively. Furthermore, grey literature, commentaries, books, and newspaper reports were excluded as part of the limitations.

## Conclusion

This study found that the MTM framework has great potential in predicting the initiation and maintenance of health behavior changes as a comprehensive, two-stage theoretical model. Since its inception, MTM has been validated in various populations abroad and has shown good effects in predicting and intervening in different types of health behavior changes. Currently, the research and application of MTM theory are mainly focused on promoting healthy behaviors, and it has been proven to be effective. However, as one of the theories in the field of health management, MTM can also be of great help in treating diseases, so it can be worth considering applying it to research on diseases. Meanwhile, there is an urgent need to explore the development of comprehensive and integrated assessment tools based on the MTM framework that are suitable for various types of applications in the future. MTM clarifies the intentions behind behavior change and its long-term sustainability, and is a highly promising theoretical model. This study provides strong evidence for the effective application of MTM theory in the field of medicine.

## Supporting information

Literature quality evaluation data.

PRISMA-ScR checklist

## Data Availability

All data produced in the present work are contained in the manuscript

## Acknowledgements

We would like to thank all the workers who worked hard on this article. It is our communication efforts that made this article possible.

Panpan Huai: Methodology, Writing – original draft, Writing – review & editing. Linghui Zhang: Methodology, Writing –original draft, Writing – review & editing. Bo Zhang: Writing – original draft, Writing – review & editing, Methodology. Yao Li: Conceptualization, Data curation, Writing – original draft, Writing – review & editing. Bing Wu: Conceptualization, Data curation, Writing – original draft, Writing – review & editing. Huimei Lv: Writing – original draft, Writing – review & editing, Methodology, Supervision. Hui Yang: Writing – original draft, Writing – review & editing, Methodology, Supervision.

**Supporting information**

**S1 Fig. Flow chart.**

**S2 Fig. Article per year.**

**S3 Fig. Number of publications by country.**

**S4 Fig. Article type.**

**S5 Fig. The number of samples in a quantitative intervention article.**

**S1 Table. SPIDER framework and PCC framework.**

**S1 File. Characteristics of included studies.**

**S2 File. Information for intervention articles.**

## References

1. Xiaopeng, Z., et al., Review on the theory of health behavior change in chronic diseases. 2022.

2. S, M. and W. JC, Health Behaviour Theory in Health Informatics: Support for Positive Change. Studies In Health Technology And Informatics, 2019: p. 146–158.

3. Brooks, S.P., et al., A framework to guide storytelling as a knowledge translation intervention for health-promoting behaviour change. Implementation Science Communications, 2022(No.1): p. 1-13.

4. Manoj Sharma, A.A.S.K., Mini review: possible role of the multi-theory model of health behavior change in designing substance use prevention and treatment interventions. Frontiers in public health, 2024: p. 1298614.

5. Martin, B. and V.K. Nahar, Theoretical Foundations of Health Education and Health Promotion. Perspectives in public health, 2017(No.6): p. 348.

6. Barati, M. and H. Abbasi, Multi-Theory Model for Changing Health Behavior: Evolutionary Process and Strengths and Weak Points. Journal of Education and Community Health, 2019(No.1): p. 1-2.

7. Gharouni, K., et al., Application of Freire’s adult education model in modifying the psychological constructs of health belief model in self-medication behaviors of older adults: a randomized controlled trial. BMC Public Health, 2020(No.1): p. 1350.

8. Matera, C., et al., Perceived Economic Uncertainty and Fertility Intentions in Couples: A Dyadic Extension of the Theory of Planned Behaviour. Journal of Family and Economic Issues, 2023(No.4): p. 790-806.

9. Ambrose-Spano, C.L.T.U., Teacher self-efficacy: Including students with emotional behavior disability in mainstream elementary classrooms. Dissertation Abstracts International: Section B: The Sciences and Engineering, 2022(No.12-B).

10. Social-cognitive predictors of doping use: an integrative approach LAMBROS LAzURAS. The Psychology of Doping in Sport, 2015: p. 70–88.

11. Salovey, P., et al., 19 - Emotional Intelligence. Feelings and Emotions, 2012: p. 321–340.

12. Valle, M., K.M. Kacmar and S. Zivnuska, Understanding the Effects of Political Environments on Unethical Behavior in Organizations. Journal of Business Ethics, 2019(No.1): p. 173-188.

13. Nicholls, G.K., Relationship—The heart of helping people: by Helen Harris Perlman. Univ. Chicago Press, Ill., 1979. 236 pp. $10.50. Social Science & Medicine. Part C: Medical Economics, 1979(No.4): p. 242.

14. Kern, A.P., et al., URBAN, ENVIRONMENTAL AND HABITABILITY PARAMETERS FOR SOCIAL HOUSING. Revista de Gestao Social e Ambiental, 2023(No.10).

15. Hughes, J., A. Randazzo and R. Ajja, MAINTENANCE OF HEALTH BEHAVIOR CHANGE: RECOMMENDATIONS FOR FUTURE RESEARCH. Innovation in Aging, 2023(Suppl 1): p. 390.

16. Qiu, R. and Y. Gu, Interpretation of the PRISMA extension for scoping review (PRISMA-ScR). Chinese Journal of Evidence-Based Medicine, 2022(No.6): p. 722-730.

17. Ernesto Calderon Martinez, J.R.F.V., Ten Steps to Conduct a Systematic Review. Cureus, 2023(No.12): p. e51422.

18. Danielle Pollock, E.L.D.M., Undertaking a scoping review: A practical guide for nursing and midwifery students, clinicians, researchers, and academics. Journal of advanced nursing, 2021(No.4): p. 2102-2113.

19. Tricco, A.C., et al., PRISMA Extension for Scoping Reviews (PRISMA-ScR): Checklist and Explanation. Annals of Internal Medicine, 2018(No.7): p. 467-473.

20. Buus, N., et al., Arksey and O’Malley’s consultation exercise in scoping reviews: A critical review. J Adv Nurs, 2022(No.8): p. 2304-2312.

21. Cooke, A.O.C.M., D.U.O.M. Smith and A.O.S.O. Booth, Beyond PICO: The SPIDER tool for qualitative evidence synthesis. Qualitative Health Research, 2012(No.10): p. 1435-1443.

22. Sze Ling Chan, C.Z.H.H., Frameworks for measuring population health: A scoping review. PloS one, 2024(No.2): p. e0278434.

23. Hayes, T., V.K. Nahar and M. Sharma, Predicting Physical Activity Behavior in African American Females: Using Multi Theory Model. J Res Health Sci, 2018. 18(2): p. e00410.

24. Zhang, W., et al., Predicting Physical Activity in Chinese Pregnant Women Using Multi-Theory Model: A Cross-Sectional Study. Int J Environ Res Public Health, 2022. 19(20).

25. Nahar, V.K., et al., Testing multi-theory model (MTM) in predicting initiation and sustenance of physical activity behavior among college students. Health Promot Perspect, 2016. 6(2): p. 58–65.

26. Hayes, T., et al., Using the Multi-Theory Model (MTM) of Health Behavior Change to Explain Yoga Practice. Altern Ther Health Med, 2022. 28(1): p. 12–17.

27. Dai, C.L., C.C. Chen and M. Sharma, Exploring Yoga Behaviors among College Students Based on the Multi-Theory Model (MTM) of Health Behavior Change. Int J Environ Res Public Health, 2023. 20(14).

28. Hayes, T., et al., The evaluation of a fourth-generation multi-theory model (MTM) based intervention to initiate and sustain physical activity. Health Promot Perspect, 2019. 9(1): p. 13–23.

29. Sharma, M., et al., Can the Multi-Theory Model (MTM) of Health Behavior Change Explain the Intent for People to Practice Meditation? J Evid Based Integr Med, 2021. 26: p. 2515690X211064582.

30. Sharma, M., et al., Introspective Meditation before Seeking Pleasurable Activities as a Stress Reduction Tool among College Students: A Multi-Theory Model-Based Pilot Study. Healthcare (Basel), 2022. 10(4).

31. Brown, L., et al., Efficacy testing of the SAVOR (Sisters Adding Fruits and Vegetables for Optimal Results) intervention among African American women: A randomized controlled trial. Health Promot Perspect, 2020. 10(3): p. 270–280.

32. Joveini, H., et al., The effect of empowerment program to reduce Sugar Consumption based on the Multi-Theory Model on Body Mass Index and abdominal obesity in Iranian women. BMC Womens Health, 2023. 23(1): p. 207.

33. Xhakollari, V., M. Canavari and M. Osman, Why people follow a gluten-free diet? An application of health behaviour models. Appetite, 2021. 161: p. 105136.

34. Sharma, M., et al., Applying Multi-Theory Model (MTM) of Health Behavior Change to Predict Water Consumption Instead of Sugar-Sweetened Beverages. J Res Health Sci, 2017. 17(1): p. e00370.

35. Sharma, A., et al., Determining predictors of change in sugar sweetened beverage consumption behaviour among university students in India. Int J Adolesc Med Health, 2020. 34(1).

36. Sharma, M., et al., Explaining the Correlates of Eating Outside-of-Home Behavior in a Nationally Representative US Sample Using the Multi-Theory Model of Health Behavior Change: A Cross-Sectional Study. Int J Environ Res Public Health, 2024. 21(1).

37. Yoshany, N., et al., Predictors in Initiating and Maintaining Nutritional Behaviors to Deal With Menopausal Symptoms Based on Multi-Theory Model. Int Q Community Health Educ, 2021: p. 272684X21991010.

38. Sharma, M., et al., Using a Multitheory Model to Predict Initiation and Sustenance of Fruit and Vegetable Consumption Among College Students. J Am Osteopath Assoc, 2018. 118(8): p. 507–517.

39. Williams, J.L., et al., Using multi theory model (MTM) of health behavior change to explain intention for initiation and sustenance of the consumption of fruits and vegetables among African American men from barbershops in Mississippi. Health Promot Perspect, 2020. 10(3): p. 200–206.

40. Gupta, A., et al., Effect of health promotion interventions on small portion size consumption behavior among college students. Indian J Public Health, 2023. 67(3): p. 435–441.

41. Sharma, M., et al., Explaining Handwashing Behavior in a Sample of College Students during COVID-19 Pandemic Using the Multi-Theory Model (MTM) of Health Behavior Change: A Single Institutional Cross-Sectional Survey. Healthcare (Basel), 2021. 9(1).

42. Panjwani, D., et al., Novel behavioral model in evaluating initiation and sustenance of teeth brushing behavior among students pursuing health sciences: a cross-sectional study. F1000Res, 2022. 11: p. 389.

43. Chandra, A., M. Sharma and V. Nahar, Exploring the efficacy of the multi-theory model (MTM) in understanding the intention for PAP adherence among recently diagnosed sleep apnea patients. Sleep Medicine, 2024(Suppl 1): p. 104.

44. Sharma, A., et al., Predictors of behaviour change for unhealthy sleep patterns among Indian dental students. Int J Adolesc Med Health, 2020. 33(5).

45. Nerida, T.M., et al., COVID-19 Vaccine Acceptance Behavior among Hispanics/Latinxs in Nevada: A Theory-Based Analysis. Healthcare (Basel), 2023. 11(5).

46. Popelsky, B.K., et al., Assessing Attitudes and Beliefs Toward HPV Vaccination among Ghanaian Parents with Unvaccinated Adolescents: Application of Multi-Theory Model of Behavior Change. Asian Pac J Cancer Prev, 2022. 23(6): p. 1901–1911.

47. Batra, K., et al., COVID-19 Booster Vaccination Hesitancy in the United States: A Multi-Theory-Model (MTM)-Based National Assessment. Vaccines (Basel), 2022. 10(5).

48. Batra, K., et al., COVID-19 vaccination hesitancy for children: A pilot assessment of parents in the United States. Health Promot Perspect, 2022. 12(4): p. 391–398.

49. Mohamed, A.A., et al., Hesitancy in COVID-19 Vaccine Uptake and Its Correlated Factors Using Multi-Theory Model among Adult Women: A Cross-Sectional Study in Three States of Somalia. Vaccines (Basel), 2023. 11(9).

50. Su, Y., et al., Exploring the Influencing Factors of COVID-19 Vaccination Willingness among Young Adults in China. Int J Environ Res Public Health, 2023. 20(5).

51. Asare, M., et al., Multi-Theory Model and Predictors of Likelihood of Accepting the Series of HPV Vaccination: A Cross-Sectional Study among Ghanaian Adolescents. Int J Environ Res Public Health, 2020. 17(2).

52. Sharma, M., K. Batra and R. Batra, A Theory-Based Analysis of COVID-19 Vaccine Hesitancy among African Americans in the United States: A Recent Evidence. Healthcare (Basel), 2021. 9(10).

53. Sharma, M., et al., A multi-theory model based analysis of correlates for initiating and sustaining mammography screening behavior among Hispanic American women in the United States. Health Promot Perspect, 2022. 12(1): p. 110–119.

54. Sharma, M., et al., Explaining Correlates of Cervical Cancer Screening among Minority Women in the United States. Pharmacy (Basel), 2022. 10(1).

55. Sharma, M., et al., Using the Multi-Theory Model (MTM) of Health Behavior Change to Explain the Seeking of Stool-Based Tests for Colorectal Cancer Screening. Int J Environ Res Public Health, 2023. 20(16).

56. Sharma, M., et al., Using the Multi-Theory Model (MTM) of Health Behavior Change to Explain the Correlates of Mammography Screening among Asian American Women. Pharmacy (Basel), 2021. 9(3).

57. Yoshany, N., et al., Effect of the fourth generation multi-theory model intervention on the quality of life in Iranian postmenopausal women: A randomized controlled trial. Post Reprod Health, 2021. 27(4): p. 189–197.

58. Morowatisharifabad, M.A., et al., Effects of an educational intervention based on the multi-theory model on promoting the quality of life in postmenopausal women: a protocol. Prz Menopauzalny, 2019. 18(3): p. 153–160.

59. Yoshany, N., et al., Predictors of regular physical activity behavior and quality of life in post-menopausal Iranian women based on the multi-theory model. J Med Life, 2022. 15(3): p. 408–414.

60. Bashir, M., F.A. Shokravi and A.K. Nezhad, Applying Multi-Theory Model (MTM) in Determining Intentions to Smoking Cessation among male Health Worker Smokers in Kabul, Afghanistan. 2024.

61. Sharma, M., et al., Assessing the Testability of the Multi-Theory Model (MTM) in Predicting Vaping Quitting Behavior among Young Adults in the United States: A Cross-Sectional Survey. Int J Environ Res Public Health, 2022. 19(19).

62. Nahar, V.K., et al., Utilizing Multi-Theory Model in Determining Intentions to Smoking Cessation Among Smokers. Tob Use Insights, 2019. 12: p. 1179173X19843524.

63. Bashirian, S., et al., Water Pipe Smoking Reduction in the Male Adolescent Students: An Educational Intervention Using Multi-Theory Model. J Res Health Sci, 2019. 19(1): p. e00438.

64. Bashirian, S., et al., Male students’ experiences on predictors of waterpipe smoking reduction: A qualitative study in Iran. Tob Prev Cessat, 2019. 5: p. 30.

65. Kumar, V., et al., Effectiveness of tobacco cessation counselling and behavioural changes Using Multi Theory Model (MTM): A follow-up study. Indian J Dent Res, 2021. 32(1): p. 56–60.

66. Sharma, M., et al., Utility of Multi-Theory Model (MTM) to Explain the Intention for PAP Adherence in Newly Diagnosed Sleep Apnea Patients. Nat Sci Sleep, 2021. 13: p. 263–271.

67. Qin, G., C. Limei and W. Xiujian, Effect of diet guidance based on the MTM model on taste change in patients with gastrointestinal cancer undergoing chemotherapy. Nursing Practice and Research, Dec.2022, Vol.19, No.24, 2022(第24期): p. 3762-3768.

68. Ginting, R., et al., Analysis of Quality Function Deployment (QFD) and Analytical Network Process (ANP) Methods at PT. XYZ. IOP Conference Series: Materials Science and Engineering, 2021: p. 012044.

69. Johnson, A.E., et al., Uncertainty and risk during the COVID-19 pandemic: A latent profile analysis. Social & Personality Psychology Compass, 2023(No.12): p. 1.

70. Xu, D., et al., Construction and Application of Health Behavior Change Intervention Ontology. Chinese Journal of Biomedical Engineering, 2023(No.1): p. 74-81.

71. Al-Matalka, S.K., et al., Health-Promoting Lifestyle Behaviors Among Jordanian University Students. AMERICAN JOURNAL OF OCCUPATIONAL THERAPY, 2023.

72. Junger, M. and M. Dekovid, Crime as Risk-Taking: Co-occurrence of Delinquent Behavior, Health-Endangering Behaviors, and Problem Behaviors. Control Theories of Crime and Delinquency, 2017: p. 213–248.

73. Katalin Fusz, Z.K.A.P., Health behavior, sleep quality and subjective health status among Hungarian nurses working varying shifts. Work (Reading, Mass.), 2021(No.1): p. 171-180.

74. LIAO Yanming, Z.X.X.Y., Differences in Health-related Behaviors and Quality of Life among Older Adults with Multimorbidity Based on Latent Class Analysis. Zhongguo quanke yixue, 2024(No.17): p. 2124-2129.

75. Peter D. Mills Ph. D., M.S., et al., Inpatient suicide on mental health units in Veterans Affairs (VA) hospitals: Avoiding environmental hazards. General Hospital Psychiatry, 2013(No.5): p. 528-536.

76. Selman Kesicia, S.Y.A.B., Get rid of the bad first. Proceedings of the National Academy of Sciences of the United States of America, 2020(No.23): p. 12526-12527.

77. Zhang, W., et al., Research progress on the application of Multi-Theory Model for health behavior change in health management. Chinese Journal of Nursing, 2022(No.15): p. 1893-1898.

