## Supplementary material for "Application of multi-theory model(MTM)health behavior change: A scoping review": Literature quality evaluation data.: Literature quality evaluation data..docx

**Results of the evaluation of the quality of qualitative research literature.**

|  |  | Q1 | Q2 | Q3 | Q4 | Q5 | Q6 | Q7 | Q8 | Q9 | Q10 | Grade |
| --- | --- | --- | --- | --- | --- | --- | --- | --- | --- | --- | --- | --- |
| 1 | (Yue Su et al., 2023) | Yes | Yes | Yes | Yes | Yes | Yes | Yes | Yes | Yes | Yes | A |
| 2 | (Saeed Bashirian et al., 2019) | Yes | Yes | Yes | Yes | Yes | Unclear | Yes | No | Yes | Yes | B |
| 3 | (Zhu et al., 2023) | Yes | Yes | Yes | Yes | Yes | No | Yes | Unclear | Yes | Yes | B |
| 4 | (Chia-Liang Dai et al., 2023) | Yes | Yes | Yes | Yes | Yes | No | Yes | Yes | Yes | Yes | B |

There are 10 evaluation criteria, each of which is evaluated by “yes,” “no,” “unclear,” and “not applicable,” and grade A is for meeting all criteria with the lowest probability of bias; grade B is for meeting part of the criteria with a moderate probability of bias; and grade C is for not meeting the criteria at all with the highest probability of bias. Grade A is to satisfy all criteria with the least possibility of bias; grade B is to satisfy part of the criteria with a moderate possibility of bias; and grade C is to not satisfy the criteria at all with the highest possibility of bias.

Q1. Is there congruity between the stated philosophical perspective and the research methodology?

Q2. Is there congruity between the research methodology and the research question or objectives?

Q3. Is there congruity between the research methodology and the methods used to collect data?

Q4. Is there congruity between the research methodology and the representation and analysis of data?

Q5. Is there congruity between the research methodology and the interpretation of results?

Q6. Is there a statement locating the researcher culturally or theoretically?

Q7. Is the influence of the researcher on the research, and vice- versa, addressed?

Q8. Are participants, and their voices, adequately represented?

Q9. Is the research ethical according to current criteria or, for recent studies, and is there evidence of ethical approval by an appropriate body?

Q10. Do the conclusions drawn in the research report flow from the analysis, or interpretation, of the data?

**Quai-experimental studies Quality Evaluation Form.**

|  |  | Q1 | Q2 | Q3 | Q4 | Q5 | Q6 | Q7 | Q8 | Q9 |
| --- | --- | --- | --- | --- | --- | --- | --- | --- | --- | --- |
| 1 | Atul Gupta et al.,2023 | Yes | No | Yes | Yes | Yes | Yes | Yes | Yes | Yes |
| 2 | Hamid Joveini et al.,2023 | Unclear | Yes | Yes | Yes | Yes | Yes | Yes | Yes | Yes |

The JBI Quality Assessment Tool for Quai-experimental studies consists of nine entries that evaluate the overall quality of classical experimental studies in terms of causality of study variables, baseline, control, measurement of outcome indicators, and analysis of the data, each of which is adjudicated using Yes, No, Unclear, and Not Applicable.

Note: Q1: Were the causal relationships in the study clearly stated?Q2: Was the baseline comparable between groups?Q3: Did the groups receive the same measures other than the intervention to be validated?Q4: Was a control group established?Q5: Were multidimensional measurements of the outcome indicators made pre- and post-intervention?Q6: Was the follow-up complete, and, if incomplete, was the loss of follow-up reported and measures taken to Q7: Were the outcome indicators measured in the same way for each group of study participants? Q8: Were the outcome indicators measured credible way? Q9: Were the data analyzed appropriately?

**Results of Literature Quality Evaluation of Non-Randomized Experiments.**

|  |  | Q1 | Q2 | Q3 | Q4 | Q5 | Q6 | Q7 | Q8 | Q9 | Q10 | Q11 | Q12 | Grade |
| --- | --- | --- | --- | --- | --- | --- | --- | --- | --- | --- | --- | --- | --- | --- |
| 1 | Vijay Kumar et al.,2021 | 2 | 1 | 2 | 2 | 0 | 1 | 0 | 2 | 0 | 0 | 0 | 2 | Moderate |
| 2 | Mohammad Ali Morowatisharifabad et al.,2020 | 2 | 2 | 2 | 2 | 2 | 1 | 1 | 2 | 2 | 2 | 1 | 2 | High |
| 3 | Manoj Sharma et al., 2022 | 1 | 1 | 2 | 2 | 2 | 0 | 2 | 2 | 0 | 0 | 0 | 2 | Moderate |

The risk of bias of the included non-randomized controlled trials was assessed using the methodological evaluation indexes for non-randomized controlled trials (MINORS), which consisted of 12 entries, each of which was scored from 0 to 2, with 0 indicating that the trial was not reported, 1 indicating that the trial was reported but with insufficient information, and 2 indicating that the trial was reported and provided sufficient information. 0 to 8 was classified as low quality literature, 9 to 16 as moderate quality literature, 17 to 24 as high quality literature.

Q1.A clearly stated aim: the question addressed should be precise and relevant in the light of available literature.

Q2.Inclusion of consecutive patients: all patients potentially fit for inclusion (satisfying the criteria for inclusion) have beenincluded in the study during the study period (no exclusion or details about the reasons for exclusion).

Q3.Prospective collection of data: data were collected according to a protocol established before the beginning of the study.

Q4.Endpoints appropriate to the aim of the study: unambiguous explanation of the criteria used to evaluate the main outcome which should be in accordance with the question addressed by the study. Also, the endpoints should be assessed on an intention-to-treat basis.

Q5.Unbiased assessment of the study endpoint : blind evaluation of objective endpoints and double-blind evaluation of subjective endpoints. Otherwise the reasons for not blinding should be stated.

Q6.Follow-up period appropriate to the aim of the study: the follow-up should be sufficiently long to allow the assessment of the main endpoint and possible adverse events.

Q7. Loss to follow up less than 5% : all patients should be included in the follow up. Otherwise, the proportion lost to follow up should not exceed the proportion experiencing the major endpoint.

Q8. Prospective calculation of the study size: information of the size of detectable difference of interest with a calculation of 95% confidence interval, according to the expected incidence of the outcome event, and information about the level for statistical significance and estimates of power when comparing the outcomes.

Q9.An adequate control group: having a gold standard diagnostic test or therapeutic intervention recognized as the optimal intervention according to the available published data.

Q10.Contemporary groups: control and studied group should be managed during the same time period (no historical comparison).

Q11. Baseline equivalence of groups: the groups should be similar regarding the criteria other than the studied endpoints. Absence of confounding factors that could bias the interpretation of the results.

Q12. Adequate statistical analyses: whether the statistics were in accordance with the type of study with calculation of confidence intervals or relative risk.

**Quality assessment of included cross-sectional studies (Using the AHRQ Cross-Sectional/Prevalence Study Quality Scale ).**

| Study | Item | | | | | | | | | | | Total scores | Quality assessment |
| --- | --- | --- | --- | --- | --- | --- | --- | --- | --- | --- | --- | --- | --- |
|  | A | B | C | D | E | F | G | H | I | J | K |  |  |
| Tara Marie Nerida et al.,2023 | Yes | Yes | Yes | Yes | No | Yes | No | Yes | Unclear | Yes | Unclear | 7 | Moderate |
| Laleh Hassani et al., 2024 | Yes | Yes | Unclear | Yes | Unclear | Yes | No | Unclear | Yes | No | No | 5 | Moderate |
| Mousa Bashir et al., 2021 | Yes | No | Yes | Yes | No | Yes | No | No | No | No | Unclear | 4 | Moderate |
| Manoj Sharma et al., 2017 | Yes | No | No | Unclear | No | Unclear | No | Yes | No | Yes | No | 3 | Low |
| Braden K Popelsky et al., 2022 | Yes | Yes | No | Yes | No | Yes | No | Yes | No | No | Yes | 6 | Moderate |
| Manoj Sharma et al., 2022(1) | Yes | Yes | Yes | Yes | Yes | Yes | Unclear | Yes | No | Yes | No | 8 | High |
| Manoj Sharma et al., 2021(1) | Yes | Yes | No | Yes | Unclear | Yes | No | Unclear | No | Yes | Yes | 6 | Moderate |
| Traci Hayes et al., 2021 | Yes | Yes | Yes | Yes | No | Unclear | No | No | No | No | No | 4 | Moderate |
| Kavita Batra et al., 2022 | Yes | Yes | Yes | Yes | Unclear | Yes | No | Yes | No | No | Unclear | 6 | Moderate |
| Manoj Sharma et al., 2022(2) | Yes | Yes | Yes | Yes | Yes | Yes | No | Yes | No | Yes | No | 8 | High |
| Manoj Sharma et al., 2021(2) | Yes | Yes | Yes | Yes | No | Yes | No | Yes | No | No | No | 6 | Moderate |
| Manoj Sharma et al., 2021(3) | Yes | Yes | No | Yes | Unclear | Yes | No | No | No | No | No | 4 | Moderate |
| Manoj Sharma et al., 2024(1) | Yes | Yes | Yes | Yes | Yes | Yes | No | Yes | No | No | No | 7 | Moderate |
| Vinayak K. Nahar et al., 2020 | Yes | Yes | No | Yes | Yes | Yes | No | Unclear | No | No | No | 5 | Moderate |
| Adam A. Mohamed et al., 2023 | Yes | Yes | No | Yes | No | Yes | No | Yes | No | No | No | 6 | Moderate |
| Matthew Asare et al.,2020 | Yes | Yes | No | Yes | Unclear | Yes | Yes | Yes | No | Yes | Yes | 8 | High |
| Dhiraj Panjwani et al., 2022 | Yes | Yes | No | Yes | No | Yes | No | No | Yes | Yes | Yes | 7 | Moderate |
| Manoj Sharma et al., 2022(3) | Yes | Yes | Yes | Yes | No | Yes | No | Yes | No | No | Unclear | 6 | Moderate |
| Traci Hayes et al., 2018 | Yes | Yes | Yes | Yes | Unclear | Yes | No | Unclear | No | No | Yes | 6 | Moderate |
| Wei Zhang et al., 2022 | Yes | Yes | Yes | Yes | No | Yes | Yes | Yes | Yes | Yes | Yes | 10 | High |
| Nooshin Yoshany et al., 2022 | Yes | Yes | Yes | Yes | No | Yes | No | No | No | No | Yes | 6 | Moderate |
| Ankur Sharma et al., 2021 | Yes | Yes | No | Yes | No | Yes | Yes | Unclear | No | Yes | Yes | 7 | Moderate |
| Nooshin Yoshany et al., 2022 | Yes | Yes | Yes | Yes | Unclear | Yes | No | No | No | No | No | 5 | Moderate |
| Manoj Sharma et al., 2021(4) | Yes | Yes | Yes | Yes | Yes | Yes | No | Yes | Yes | Yes | Yes | 10 | High |
| Vinayak K. Nahar et al., 2016 | Yes | Yes | Yes | Yes | No | Yes | No | No | No | Yes | Yes | 7 | Moderate |
| Manoj Sharma et al., 2021(5) | Yes | Yes | Yes | Yes | No | Yes | No | Yes | Yes | Yes | Yes | 10 | High |
| Manoj Sharma et al., 2024(2) | Yes | Yes | Yes | Yes | Unclear | Yes | No | Yes | No | No | Yes | 7 | Moderate |
| Amanda H. Wilkerson et al., 2023 | Yes | Yes | Yes | Unclear | No | Yes | Yes | No | No | No | Yes | 6 | Moderate |
| Manoj Sharma et al., 2018(1) | Yes | Yes | Yes | Unclear | No | Yes | No | No | No | Yes | Yes | 6 | Moderate |
| Jaelrbreiret L. Williams et al., 2020 | Yes | Yes | Yes | Yes | Unclear | Yes | No | No | No | Yes | Yes | 7 | Moderate |
| Manoj Sharma et al., 2023 | Yes | Yes | No | Yes | No | Yes | No | Yes | No | Yes | No | 6 | Moderate |
| Manoj Sharma et al., 2020 | Yes | Yes | No | Yes | Unclear | Yes | No | No | No | Yes | No | 5 | Moderate |
| Traci Hayes et al., 2022 | Yes | Yes | Yes | No | No | Yes | No | Unclear | No | No | No | 4 | Moderate |
| Manoj Sharma et al., 2021(6) | Yes | Yes | Yes | Yes | Yes | Yes | No | Yes | No | No | Unclear | 7 | Moderate |
| Edith Claros et al., 2020 | Yes | Yes | No | Yes | No | Yes | No | Unclear | No | Yes | Yes | 6 | Moderate |
| Manoj Sharma et al., 2021(6) | Yes | Yes | No | Unclear | No | Yes | No | Yes | No | No | Yes | 5 | Moderate |
| Vinayak K Nahar et al., 2019 | Yes | Yes | Yes | No | No | Yes | No | Yes | No | No | Yes | 6 | Moderate |
| Julia Wells et al., 2023 | Yes | Yes | Yes | Unclear | No | Yes | No | No | No | No | No | 4 | Moderate |
| Vilma Xhakollari et al., 2021 | Yes | Yes | Yes | Yes | No | Yes | No | Yes | No | No | Yes | 7 | Moderate |
| Shasha Li et al., 2021 | Yes | Yes | Yes | Unclear | No | No | No | No | No | Yes | No | 4 | Moderate |
| Jingwen Meng et al., 2024 | Yes | Yes | Yes | Yes | No | Yes | No | Yes | No | Yes | No | 7 | Moderate |
| Robert E. Davis et al., 2021 | Yes | Yes | Yes | Unclear | Unclear | Yes | No | Yes | Yes | Yes | Yes | 8 | High |
| Sidath Kapukotuwa et al., 2023 | Yes | Yes | Yes | Unclear | No | Yes | No | Yes | No | No | No | 5 | Moderate |
| Ankur Sharma1 et al., 2020 | Yes | Yes | Yes | No | No | Yes | No | Yes | No | Yes | No | 6 | Moderate |
| Kavita Batra et al., 2022 | Yes | Yes | Yes | Yes | No | Yes | No | Yes | No | Yes | Yes | 8 | High |
| Manoj Sharma et al., 2022(4) | Yes | Yes | No | Unclear | No | Yes | No | Yes | No | Yes | Yes | 6 | Moderate |
| Manoj Sharma et al., 2018(2) | Yes | Yes | Yes | Yes | No | Yes | No | Yes | No | Yes | Yes | 8 | High |
| Manoj Sharma et al., 2021(7) | Yes | Yes | Yes | Yes | Yes | Yes | No | Yes | No | Yes | Unclear | 8 | High |

The cross-sectional study evaluation criteria proposed by the Agency for Healthcare Research and Quality (AHRQ) for the assessment of bias risk. These standards consist of 11 items. For each item, the answer “Yes” received 1 point, and the answers “No” or “Unclear” received 0 points, for a total possible score of 11 points. A higher total score indicated better quality of the literature. A score ≤3 was classified as low quality, 4–7 as moderate quality, and ≥8 as high quality.

* A: Define the source of information (survey, record review); B: List inclusion and exclusion criteria for exposed and unexposed subjects (cases and controls) or refer to previous publications; C: Indicate time period used for identifying patients; D: Indicate whether or not subjects were consecutive if not population-based; E: Indicate if evaluators of subjective components of study were masked to other aspects of the status of the participants; F: Describe any assessments undertaken for quality assurance purposes (e.g., test/retest of primary outcome measurements); G: Explain any patient exclusions from analysis; H:Describe how confounding was assessed and/or controlled; I: If applicable, explain how missing data were handled in the analysis; J: Summarize patient response rates and completeness of data collection; K: Clarify what follow-up, if any, was expected and the percentage of patients for which incomplete data or follow-up was obtained.

**Risk of Bias in RCT.**
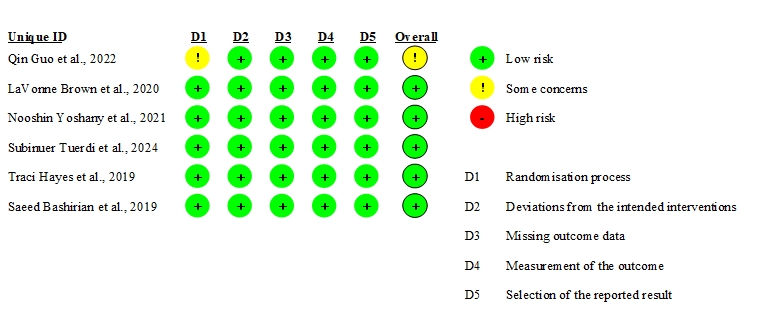
