## Supplementary material for "Application of multi-theory model(MTM)health behavior change: A scoping review": Table 1: S1_File.pdf

**Table 1. SPIDER framework and PCC framework.**

| SPIDER（Sample、Phenomenon of Interest、Design、Evaluation、Study Type） | |
|---|---|
| Sample | All groups |
| Phenomenon of Interest | Application of MTM theory to health behavior change |
| Design | Questionnaire adjustment, scale evaluation, interview dialogue and so on |
| Evaluation | Outcomes such as participatory dialogue, behavioral confidence, change in physical environment, emotional transformation, practice for change, and change in social environment were evaluated |
| Study Type | Quantitative survey and qualitative survey |
| PCC(population、concept、context） | |
| Population | All groups |
| Concept | MTM theory of health behavior change |
| Context | Application of MTM theory to health behavior change |
