## Supplemental Table 2 for "Application of multi-theory model(MTM)health behavior change: A scoping review": S2_File.pdf

### File 1. Characteristics of included studies.

| Article Title | Author, year, journal | Country | Research purpose | Participants | Behavior type | Research type | Research method | Data analysis | Research limitation |
|---|---|---|---|---|---|---|---|---|---|
| COVID-19 Vaccine Acceptance Behavior among Hispanics/Latinxs in Nevada: A Theory-Based Analysis | Tara Marie Nerida, Manoj Sharma.(2023).HEALTHCARE | America | The study was designed to use a multi-theoretical model of Health Behavior Change (MTM) to explain the intent to initiate and maintain COVID-19 vaccination behavior in Hispanic and Latino populations in Nevada who expressed and did not express hesitation about the vaccine. | 263 | COVID-19 Vaccine Acceptance Behavior. | A quantitative cross-sectional and survey-based research study design | Using a quantitative cross-sectional and survey-based research study design data were collected using a 50-item questionnaire and analyzed using multiple linear regression modeling. | The survey data from Qualtrics were further analyzed in SPSS . Descriptive statistical analysis was conducted for all study variables. A zero-order correlation matrix was conducted among the construct variables to identify if there were any significant, simple bivariate relationships between the theoretical constructs and both the initiation and sustenance for the hesitant and non-hesitant groups. Hierarchical multiple regression was used to “control” for certain variables among different groups to see if adding variables improved the model’s capacity to predict the likelihood of getting the COVID-19 vaccine and/or the second dose/booster dose; this was used to study the hesitant and non-hesitant groups and their relationship with the two outcome variables of initiation and sustenance, which formed four models. | The study utilized a cross-sectional study which may not determine if an association equals causation or the directionality of the outcome. Additionally, as with any self-reported survey study design, one limitation was response bias. Recruitment bias may have occurred due to the difficulty of obtaining participants early in the recruitment stages. Another limitation was that the survey instrument had a Flesch Reading Ease score of 52.3 and a Flesch–Kincaid Grade Level of 9.9, which made the survey fairly difficult to read. The sample collected contained responses from predominantly females (69%) and people of Mexican identity (63.2%), all of whom resided in Nevada. This limits the generalizability of the study findings to all genders and other Hispanic/Latinx identities outside of Nevada. Another quantitative study design that would help to generalize the study’s findings could be conducted using a larger population sample; however, a qualitative study design utilizing interviews and focus groups may help to gain a deeper understanding of the participatory dialogue and behavioral confidence that would affect the initiation of the vaccine, as well as the emotional transformation of the |

| Article Title | Author, year, journal | Country | Research purpose | Participants | Behavior type | Research type | Research method | Data analysis | Research limitation |
|---|---|---|---|---|---|---|---|---|---|
| | | | | | | | | | sustenance of the vaccine. |
| A multi-theory model-based analysis of correlates for initiating and sustaining mammography screening behavior among Hispanic American women in the United States | Manoj Sharma, Kavita Batra.(2022) .HEALTH PROMOTION PERSPECTIVES | America | Cross-sectional study aimed to explore correlates of mammography screening behavior among a sample of Hispanic women aged 45-54 years living in the United States using the multi-theory model (MTM). | 370 | Initiating and sustaining mammography screening behavior. | Non-random sampling procedures | A 50-item web-based survey consisting of psychometrically valid tools based on MTM theoretical framework was administered through non-random sampling procedures using Qualtrics. Univariate, bivariate, and multivariate statistics were used to analyze the data. | SPSS software v.26 was used to analyze the data. All types of analytic methods, including univariate, bivariate, and multivariate statistics were utilized. First, univariate statistics were calculated to describe the characteristics of the sample. As an uncertainty measure, 95% confidence intervals of proportion were calculated through normal approximation to the binomial distribution. Initial model and assumptions (e.g. independence of residuals, linearity, equal error variance, multicollinearity, and normality of residuals) were tested prior to the predictive modelling. Comparisons across categorical and continuous outcomes were derived through Chi-square and independent sample t-tests respectively (bivariate tests). Pearson correlation was utilized to investigate bivariate relationships between the observed variables. Two separate (one for initiation and one for sustenance) hierarchical regression models were used to explain the change in variance in the dependent variables (initiation and sustenance of mammography screening) attributed to the sequential addition of independent variables. | / |
| An intervention to improve antibiotic | Laleh Hassani. (2024).HEA | Iran | The present study aims to develop, implement, and evaluate the effectiveness of an | / | Antibiotic prescription behavior in | A cross-sectional study | The present study will include four phases including a qualitative phase, an instrument design and | The data will be analyzed in MAXQDA 10. In the second phase, the face and content validity will be tested by a panel of experts as field specialists. A confirmatory factor analysis | Although the random assignment of students to two intervention and control groups leads to the same distribution of confounding variables and prevents selection |

| Article Title | Author, year, journal | Country | Research purpose | Participants | Behavior type | Research type | Research method | Data analysis | Research limitation |
|---|---|---|---|---|---|---|---|---|---|
| prescription behavior in veterinary students: A protocol based on the multi-theory model to tackle antimicrobial resistance | LTH SCIENCE REPORTS | | educational program based on the multi-theoretical model (MTM) in improving antibiotic prescription behavior in veterinary students of Iran. | | veterinary students. | | psychometric test phase, and a cross-sectional, and an interventional phase. In the first phase, the sampling will be purposive with a maximum variety. The interviews will be conducted with a sample of veterinarians. | will be used to test construct validity, and Cronbach's alpha coefficient and intracluster correlation coefficient will be used to determine the internal consistency of the instrument. Then, at this stage, a number of veterinary students will be selected through a multi-stage sampling method. In the cross-sectional phase, another sample of veterinary students will complete a researcher-made questionnaire. Then, Spearman's correlation coefficient test will be used to test the relationship between the two stages of behavior initiation and behavior continuation. The data will be analyzed in SPSS 22. In the third phase, some veterinary students will be selected through a census and will be randomly divided into a control and an intervention group. To collect data in the final phase, the researcher-made questionnaire that was designed in the second phase of the study based on a multi-theory model will be used to extract data. To compare demographic characteristics, compare the correlation between the constructs of the multi-theory model with antibiotic prescribing behavior in the cross-sectional phase and compare the scores of the constructs of the MTM in two intervention and control groups paired-samples T test and independent-samples T test will be used. | error, there is a possibility that some students will have more willingness and participation in the current research with more study. Therefore, it will not be possible to generalize this study to all students. Due to data collection by online questionnaire, it is not possible to explain the questions to the participants. Therefore, at the beginning of the questionnaire, the main researcher will provide his contact number for any possible questions, and in addition, in the explanation section of the questionnaire, he will try to give a brief explanation for some questions. For the participation of the students and their cooperation to adhere to the study, a small booklet will be given to all of them as a gift. Future investigations can include multilevel interventions to provide solutions to combat global threats. |
| Applying | Mousa | Iran | A multi-theory model (MTM) | 170 | Smoking cessation | A cross-sectional | By visiting different Health Centers, | A descriptive analysis was performed on all study variables. | This study has limitations that the authors would like to |

| Article Title | Author, year, journal | Country | Research purpose | Participants | Behavior type | Research type | Research method | Data analysis | Research limitation |
|---|---|---|---|---|---|---|---|---|---|
| Multi-Theory Model (MTM) in Determining Intentions to Smoking Cessation among male Health Worker Smokers in Kabul, Afghanistan | Bashir, Farkhondeh Amin Shokravi. (2021). medRxiv | | was used to determine cessation intentions among male health worker smokers in Kabul, Afghanistan. | | intentions of male health worker smokers. | research design | a convenience sample of male health worker smokers from west part of Kabul city, was invited to participate in this cross-sectional study. A valid and reliable 37-item MTM-based survey instrument was administered to the male participants who smoked. To explain smoking cessation behavior, stepwise multiple regressions were conducted. The entire value of the Cronbach alpha coefficient (α) of the subscales and the scale for the initiation of MTM variables was 0.80 and for the sustenance of MTM variables was 0.79. | As appropriate, we calculated Spearman's r to evaluate the variables related to demographics (covariates) and dependents variables (Smoking cessation behavior initiation and maintenance). Since there were two dependent variables, in order to determine how predictive MTM constructs are, we conducted two stepwise multiple regression models in two blocks apart from factors influencing demographics. Each model began with block 1 entering demographic variables in bivariate analysis. Every MTM construct was added to block 2 for each model. A priori, 0.05 was set as the statistical significance level. The IBM SPSS V.25 is used for analyzing the data. | acknowledge. In this study, due to target population study limitation, convenience sampling method was used for male health workers who smoke. Therefore, the findings cannot be generalized beyond the sample population of the study. Also there is a need for future research to use randomized controlled designs to conduct interventional studies. The current study was restricted to only male health worker smokers. Future studies can be planned to investigate both genders. Finally, in this study our sample was from the west region of the Kabul city, so it is not representative of all regions, which may have influenced the results. |
| Applying multi-theory model (MTM) of health behavior change to predict water | Manoj Sharma, Hannah Priest Catalano. (2017).Journal of | America | The purpose of this study was to use the multi-theory model (MTM) in predicting initiation and sustenance of plain water consumption instead of sugar-sweetened beverages among college students. | 410 | Plain water consumption instead of sugar-sweetened beverages among college students. | A cross-sectional study | In this cross-sectional study, a 37-item valid and reliable MTM-based survey was administered to college students in 2016 via Qualtrics at a large public university in the Southeastern United States. Overall, 410 students | All data were analyzed using SPSS, version 21 . For descriptive statistics, means, standard deviations of metric variables, and frequencies and percentages for categorical variables were reported. With a sample size of 174 subjects and a large number of parameters in our model, SEM estimates are not stable23. Thus, we chose to use stepwise multiple regressions to build the initiation and sustenance | / |

| Article Title | Author, year, journal | Country | Research purpose | Participants | Behavior type | Research type | Research method | Data analysis | Research limitation |
|---|---|---|---|---|---|---|---|---|---|
| consumption instead of sugar-sweetened beverages | Research in Health Sciences | | | | | | responded to the survey; of those, 174 were eligible for the study and completed it. | models. For stepwise multiple regression, the a priori criteria of probability of F to enter the predictor in the model was set as less than or equal to 0.05 and for removing the predictor as greater than or equal to 0.10. | |
| Assessing Attitudes and Beliefs Toward HPV Vaccination among Ghanaian Parents with Unvaccinated Adolescents: Application of Multi-Theory Model of Behavior Change | Braden K Popelsky, Matt Asare. (2022).Asian Pacific Journal of Cancer Prevention | Ghana | The purpose of the study was to assess the attitudes and beliefs towards HPV vaccination among Ghanaian parents with unvaccinated adolescents using the Multi-Theory Model (MTM) of behavior change. | 380 | Parents with unvaccinated adolescents living in the Ashanti Region of Ghana. | A cross-sectional study | A 44-item validated survey was administered among parents with unvaccinated adolescents living in the Ashanti Region of Ghana. HPV vaccine initiation predictors were perceived beliefs and MTM constructs: participatory dialogue, behavioral confidence, and change in the physical environment. HPV vaccine completion predictors were emotional transformation, social environment, and practice for change. | The demographic variables were analyzed using descriptive statistics and multiple linear regression analyses were performed to assess the relationships between Multi-Theory Model constructs and the likelihood of parents allowing their adolescent to initiate and complete the HPV vaccine series. Structural Equation Modeling was performed to evaluate the fitness of the model. The significant result was set a priori at p-value $< 0.05$. All data were analyzed using the IBM Statistic Package for Social Sciences (SPSS version 25). For the structural equation modeling, we used AMOS. | Potential limitations of this study may be found in the design and sampling methods used. This study used a cross-sectional design with convenience sampling, which may not provide a representative sample of the parents in the Ashanti region of Ghana. Additionally, a majority of the study population were female, thus study findings may not reflect the beliefs of both male and female parents in the Ashanti region of Ghana. |
| Assessing the Testability of the Multi-Theory | Manoj Sharma, Kavita Batra. | America | This cross-sectional study aims to assess the testability of the contemporary multi-theory model of health behavior change | 619 | Quitting Behavior among Young Adults in the United States. | A cross-sectional survey | A nationally representative sample of 619 young adults engaged in vaping behavior and aged 18–24 years was recruited to complete a | The responses of the survey were analyzed using univariate, bivariate as well as multivariate statistical methods. Categorical variables were represented as counts and proportions, whereas continuous variables (if normally | Our study had some limitations. Our sample, though nationally representative for gender, national region, race, and ethnicity, was not representative of other dimensions, which could have influenced the results. This attributes to |

| Article Title | Author, year, journal | Country | Research purpose | Participants | Behavior type | Research type | Research method | Data analysis | Research limitation |
|---|---|---|---|---|---|---|---|---|---|
| Model (MTM) in Predicting Vaping Quitting Behavior among Young Adults in the United States: A Cross-Sectional Survey | (2022).INTERNATIONAL JOURNAL OF ENVIRONMENTAL RESEARCH AND PUBLIC HEALTH | | in predicting the vaping quitting behavior among young adults in the United States. | | | | 49-item web-based survey. A structural equation model was used to test relationships between MTM constructs. | distributed) were represented as means and standard deviations. In the univariate analysis, the 95% confidence intervals of proportions were computed through the binomial “exact” method. The intercorrelation matrix among continuous variables was ascertained using Pearson’s bivariate correlation test. Two separate models of the hierarchical multiple regression were built to predict or explain the variance in the dependent variables (e.g., initiation and sustenance) by independent variables, such as demographic characteristics, history of behaviors, and MTM constructs. A complete model-building process is described in Figure 3 as shown The responses of the survey were analyzed using univariate, bivariate as well as multivariate statistical methods. Categorical variables were represented as counts and proportions, whereas continuous variables (if normally distributed) were represented as means and standard deviations. In the univariate analysis, the 95% confidence intervals of proportions were computed through the binomial “exact” method. The intercorrelation matrix among continuous variables was ascertained using Pearson’s bivariate correlation test. Two separate models of the hierarchical multiple regression were built to predict or explain the variance in the dependent variables (e.g., | limited generalizability. Next, we only sampled individuals who vape and could not compare the characteristics with those who do not vape. Further, we did not differentiate those who vaped just for flavor, as our study focused on nicotine and cannabis. Future studies can be planned to investigate demographic differences among groups who vape vs. who do not. Our study used a cross-sectional design and hence we cannot make firm conclusions due to a lack of the ability to establish temporal associations. Finally, self-reported data have several limitations. Future research must undertake interventional work using randomized controlled designs. |

| Article Title | Author, year, journal | Country | Research purpose | Participants | Behavior type | Research type | Research method | Data analysis | Research limitation |
|---|---|---|---|---|---|---|---|---|---|
| | | | | | | | | initiation and sustenance) by independent variables, such as demographic characteristics, history of behaviors, and MTM constructs. | |
| Can the Multi-Theory Model (MTM) of Health Behavior Change Explain the Intent for People to Practice Meditation? | Manoj Sharma, Ram Lakhan.(2021).Journal of Evidence-Based Integrative Medicine | America | This study aimed to test if a fourth-generation multitheory model (MTM) could explain the intent for starting and maintaining meditation behavior in a sample of US adults. | 330 | Practice Meditation. | A cross-sectional design | A face and content valid 48-item instrument based on MTM was administered in a cross-sectional design through an online survey. | All data were analyzed using IBM-SPSS, Version 26.0. For descriptive purposes, metric demographic and study variables were summarized using means and standard deviations while categorical variables were summarized using frequencies and percentages. For structural equation modeling for construct validation, chi-square ($\chi 2$), comparative fit index (CFI), root mean square error of approximation (RMSEA), and standardized root mean square residual (SRMR) indices to assess the overall goodness of fit of the model were employed. A nonsignificant chi-square ($P > .05$) is desirable for the model to have a good fit. RMSEA values of ≤ 0.06 and CFI values approximating 1 indicate a good model fit. For SRMR, values < 0.10 are acceptable, with values < 0.08 as preferable. SRMR was included because it is the most sensitive to latent structures or miss-specified factor covariances. For modeling initiation and sustenance, only participants who spent < 140 min weekly on meditation and who indicated that they did not suffer from any medical condition including physical or mental disability that prevented them from performing | The study did have some limitations. In this study, actual behavior was not measured but rather the intent of meditation; a substitute for actual behavior. Future studies can address this limitation. Also, our sample consisted of mainly older adults and the majority was White so the results have to be interpreted with caution for generalizability purposes across diverse Sharma et al 9populations. The cross-sectional design and use of self reports are also methodological limitations of our study. |

| Article Title | Author, year, journal | Country | Research purpose | Participants | Behavior type | Research type | Research method | Data analysis | Research limitation |
|---|---|---|---|---|---|---|---|---|---|
| | | | | | | | | meditation were included. The perceived stress score, gender, age, race/ethnicity, education level, and employment status were used as covariates. To assess statistically significant associations between the aforementioned covariates and dependent variables (initiation and sustenance for practicing meditation behavior), Pearson product-moment correlations and one-way analysis of variance (ANOVA) were conducted. For model building, hierarchical regressions (initiation model and sustenance model) were employed in 2 blocks. In block one, only those covariates were entered which showed a statistically significant relationship with the dependent variable. Further, in the second block, MTM constructs were entered to assess their relationships with the dependent variable, after controlling for the effects of covariates. The significance level of .05 was set a priori. | |
| COVID-19 Booster Vaccination Hesitancy in the United States: A Multi-Theory-Model | Kavita Batra, Manoj Sharma, Jagdish Khubchandani. (2022).VAC | America | The purpose of the study was to investigate hesitancy, confidence, literacy, and the role of the multi-theory model (MTM) constructs in COVID-19 booster uptake. | 285 | COVID-19 Booster Vaccination Hesitancy in the United States. | A Multi-Theory-Model (MTM)-Based National Assessment | This cross-sectional study utilized a 52-item psychometric valid web-based survey conducted during the month of October 2021 to recruit a nationally representative sample of U.S. adults. Univariate, bivariate, and multivariate statistical tests were used to analyze the data. | Data were first cleaned and recoded for the analytical operations. All statistical assumptions, including the normality, homogeneity of variance, independence of residuals, and equal error variances, were assessed. Box plots were visually inspected to identify outliers in the data. Continuous variables are described as mean and standard deviation unless stated otherwise. Categorical variables were represented as frequencies and proportions. Univariate and | The results of this study should be viewed in light of several potential limitations. First, our findings are restricted by all threats to the validity and reliability inherent to crosssectional and survey study designs (e.g., socially desirable responses, non-response bias, self-selection bias, recall bias, and the inability to establish cause-and-effect relationships).Second, although the MTM is a comprehensive model, there could be other individual |

| Article Title | Author, year, journal | Country | Research purpose | Participants | Behavior type | Research type | Research method | Data analysis | Research limitation |
|---|---|---|---|---|---|---|---|---|---|
| (MTM)-Based National Assessment | CINES | | | | | | | bivariate analyses (i.e., chi-square, independent-samples t-test, and Pearson's correlation test) were used to describe the sample. In univariate statistics, 95% confidence intervals of proportion were calculated using normal approximation to the binomial distribution. Adjusted standardized residuals greater than 2 were considered significant cells for contingency tables larger than 2 × 2 chi-square analysis. A contingency table analysis using adjusted residuals (or Z scores) was performed to generate p values of multiple comparisons. Hierarchical multiple regression was run to determine Data were first cleaned and recoded for the analytical operations. All statistical assumptions, including the normality, homogeneity of variance, independence of residuals,and equal error variances, were assessed. Box plots were visually inspected to identify outliers in the data. Continuous variables are described as mean and standard deviation unless stated otherwise. Categorical variables were represented as frequencies and proportions. Univariate and bivariate analyses (i.e., chi-square, independent-samples t-test, and Pearson's correlation test) were used to describe the sample. In univariate statistics, 95% confidence intervals of proportion were calculated using normal approximation to the binomial distribution. Adjusted standardized residuals | characteristics and influential factors that could have influenced study participants' willingness to receive a booster dose for COVID-19 (e.g., side effects from previous doses of the vaccine, mandates from employers, or COVID-19 related mortality and morbidity in social networks). Third, our study sample had a higher proportion of individuals who were vaccinated with the primary series of the COVID-19 vaccine (75% in our study vs. 65% of the US population). Finally, a threat to the external validity is that the sample is limited in nature and extent (e.g., limited to those with computers or mobile phones and an understanding of the online survey environment). |

| Article Title | Author, year, journal | Country | Research purpose | Participants | Behavior type | Research type | Research method | Data analysis | Research limitation |
|---|---|---|---|---|---|---|---|---|---|
| | | | | | | | | greater than 2 were considered significant cells for contingency tables larger than 2 × 2 chi-square analysis. A contingency table analysis using adjusted residuals (or Z scores) was performed to generate values of multiple comparisons. Hierarchical multiple regression was run to determine. | |
| Applying the integrated marketing communication approach to recruit and retain African American women | Traci Hayes1, Manoj Sharma. (2021).HEALTH PROMOTION PERSPECTIVES | America | This article explores the IMC approach used to recruit and retain volunteers for a community-based intervention. | 74 | Integrated marketing communication approach. | A cross-sectional study | This is a cross-sectional study relying on extracted data from the Multi-Theory Model (MTM) of Health Behavior Physical Activity intervention. A brief multiple-choice survey was administered to a sample of African American women (n=74) to assess the effectiveness of applying an IMC approach for recruiting and retaining volunteers for the multi-week program during January - June 2018. The measures were (1) source for study information, (2) preferred method of contact, (3) primary source for health information. | The statistical analyses were conducted using IBM SPSS Version 25. Descriptive statistics including frequencies and percentages were used to explain findings. A chi-square test of independence was used to investigate whether there was an association between age groups and selection of recruitment strategy and preference of retention strategy. Statistical significance was set at alpha = 0.05. We did not control for confounding variables in the current study. | / |
| COVID-19 | Kavita | America | The purpose of this study was to | 263 | COVID-19 | A cross-sectional | To participate in this study, a | Means and standard deviation were used to present | This study is not without limitations. First, cross-sectional |

| Article Title | Author, year, journal | Country | Research purpose | Participants | Behavior type | Research type | Research method | Data analysis | Research limitation |
|---|---|---|---|---|---|---|---|---|---|
| vaccination hesitancy for children: A pilot assessment of parents in the United States | Batra. (2022).HEALTH PROMOTION PERSPECTIVES | | conduct a national assessment of parents' preferences for COVID-19 vaccination of children using the evidence-based Multi-Theory Model (MTM) and explore the predictors of vaccine hesitancy. | | vaccination hesitancy for children. | analytical study | national random sample of parents took a valid and reliable online questionnaire based on the MTM. Independent samples t test, chi-square test, multiple logistic regression was utilized to analyze data. | continuous variables, whereas categorical variables were represented as counts and percentages. Bivariate (e.g., chi-square, independent samples test, and Pearson's correlation) and multiple logistic regression tests were used to analyze the data. For the logistic regression, we used the maximum likelihood method to obtain Wald's confidence intervals and adjusted odds ratio estimates. All variables were dummy coded to allow an appropriate estimation in the regression, and probability in the logistic regression was modeled on "willingness of parents" = "yes." Statistical significance was assumed at $P < 0.05$. All data analyses were conducted using the Statistical Package for Social Sciences (SPSS) version 27 for Windows and Statistical Analysis System (SAS), version 9.4 . | nature of this study limited our ability to establish causality. Second, certain type of biases (e.g., selection, recall, and non-response bias) were inevitable. Also, some parents may have provided socially desirable responses, which might have introduced social desirability bias. Third, there could be individual characteristics and other influential factors that might have left unmeasured and could have influenced study participants' willingness to get their children vaccinated (e.g., side effects of vaccines among parents). Next, as the sample is limited in nature and extent (e.g., limited to those with computers or mobile phones and an understanding of the online survey environment), external validity will be limited. Moreover, sample was not representative, which might limit the external validity of the results. Future studies can be planned with a nationally representative sample to investigate parental hesitancy. |
| Determining predictors of change in sugar sweetened beverage consumption behaviour | Ankur Sharma. (2020).International Journal of Adolescent Medicine | India | The present study aimed to predict the SSB consumption behaviour among Indian university students by utilising a multi theory model (MTM) of health behaviour change. | 267 | Change in sugar sweetened beverage consumption behaviour among university students in India. | A cross-sectional design | In a cross-sectional design, a validated 37-item self-report questionnaire was administered to 267 participants from a mid-size university in the National Capital Region (NCR) of India. Stepwise multiple regressions were used to | All data were analysed using SPSS, version 20 analysed. For descriptive statistics, means, standard deviations of metric variables, and frequencies and percentages for categorical variables were reported. For inferential statistics, stepwise multiple regressions were used to build the initiation and sustenance models. For stepwise multiple regression, the a priori criteria of probability of F to enter the predictor in the | / |

| Article Title | Author, year, journal | Country | Research purpose | Participants | Behavior type | Research type | Research method | Data analysis | Research limitation |
|---|---|---|---|---|---|---|---|---|---|
| among university students in India | and Health | | | | | | determine predictors of change in initiation and sustenance of SSB consumption behaviour. | model was set as less than or equal to 0.05 and for removing the predictor as greater than or equal to 0.10. | |
| Explaining Correlates of Cervical Cancer Screening among Minority Women in the United States | Manoj Sharma, Kavita Batra, Christopher Johansen. (2022). pharmacy | America | This cross-sectional study attempts to examine the correlates of cervical cancer screening by Pap test using the Multi-theory Model (MTM) as a theoretical paradigm among minority women in the United States (U.S.). | 364 | Cervical cancer screening for minority women in the United States. | A cross-sectional study | Given the paucity of theory-based interventions to promote Pap smear tests among minority women, this cross-sectional study attempts to examine the correlates of cervical cancer screening by Pap test using the Multi-theory Model (MTM) as a theoretical paradigm among minority women in the United States (U.S.). Structural Equation Modelling (SEM) was done for testing the construct validity of the survey instrument. | The SPSS software v.26 was used to conduct the descriptive and inferential statistical analysis. All assumptions were tested prior to the application of statistical models. Comparison between groups for the normally distributed numeric data was conducted using the independent samples test, whereas the chi-squared test was used to compare categorical data among groups. The Pearson correlation test was used to correlate two continuous variables. Hierarchical regression was conducted by taking initiation and sustenance scores as dependent variables. Polytomous variables used in the regression were dummy-coded. The statistical significance was denoted as $p < 0.05$. Missing data analysis was not warranted as a complete dataset and was obtained from Qualtrics. | There were some limitations to this study. First, the cross-sectional nature of the design precludes making temporal causal inferences between the independent and dependent variables. Future studies must utilize longitudinal experimental designs to provide more definitive evidence. Second, the study did not collect direct data in the form of medical records of Pap tests and relied only on self-reports, which are subject to biases. While some variables such as attitudes can only be measured through self-reports, future studies must utilize more objective data for variables that can have other means of measurement. Another limitation concerns the conducting of this survey only in English, which limited our sample to only those who could read and speak English. Future studies should offer other languages, e.g., Spanish, Chinese, Japanese, etc., to potentially capture more minority women. Finally, the study did not measure the stability of the instrument by test–retest reliability coefficients. Future studies, especially those undertaking interventional research, must test the stability of the |

| Article Title | Author, year, journal | Country | Research purpose | Participants | Behavior type | Research type | Research method | Data analysis | Research limitation |
|---|---|---|---|---|---|---|---|---|---|
| | | | | | | | | | instrument. |
| Explaining Handwashing Behavior in a Sample of College Students during COVID-19 Pandemic Using the Multi-Theory Model (MTM) of Health Behavior Change: A Single Institutional Cross-Sectional Survey | Manoj Sharma, Kavita Batra. (2021).HEALTHCARE | America | The purpose of this cross-sectional study was to explore and explain the handwashing behavior among college students during the COVID-19 pandemic using a contemporary fourth-generation multi-theory model (MTM) of health behavior change. | 713 | Handwashing Behavior in a Sample of College Students during COVID-19 Pandemic. | A Single Institutional Cross-Sectional Survey | Utilizing the MTM theoretical framework, a survey instrument to assess the likelihood of initiating and sustaining handwashing behavior was developed. The instrument consisted of 36 items with six items related to participant demography. The remaining 30 items correspond to two main components of the MTM, such as initiation and sustenance, with a total of seven constructs. | Participants' responses from Qualtrics XM were exported to Microsoft Excel and then imported to IBM SPSS version 26.0. Confirmatory factor analysis using the maximum likelihood method and internal consistency diagnostic tests using Cronbach's alpha were performed to check the construct validity and internal consistency of the tool. For establishing a one-factor solution following the Kaiser criterion of Eigenvalue greater than or equal to 1.0, factor loadings on each item greater than 0.326 (after doubling the critical value for a sample size of 250 at an $\alpha = 0.01$ for a two-tailed test) were established a priori as per the generally accepted recommendations from previous literature. For establishing internal consistency reliability, a Cronbach's alpha of $\geq 0.70$ was considered acceptable. Descriptive statistics, including the frequencies, proportions, mean, and standard deviations, were generated. The likelihood of intention and sustenance of handwashing behavior were the dependent variables, while the constructs were used as independent variables. To analyze the differences in mean scores across different groups who follow or did not follow handwashing recommendations, an independent-samples t-test was utilized. Pearson's correlations test was utilized to | First, this study is based on only one large, public Southern U.S. university. Therefore, the findings may not be extrapolated to students of other institutions, and caution should be applied while inter-preting the results. However, our sample was nearly representative of the institution where study took place. University racial breakdown for previous year was White/Caucasian(73.7%), Black/African American (4.4%), Hispanic (8.6%), and Asian (2.5%).Second, the study relied on self-reported information, which can subject to measure- ment error. However, when it comes to measuring attitudes towards health behavior, this is the only method for the measurement. Third, in testing the instrument's reliability, the instrument's stability over time was not assessed. This offers a potential avenue for future research and will be especially important before conducting experimental studies. Fourth, actual availability of resources for handwashing were not measured in this study. Finally, the study used a cross-sectional design in which the independent and dependent variables are measured simultaneously, thereby preventing any causal inferences. |

| Article Title | Author, year, journal | Country | Research purpose | Participants | Behavior type | Research type | Research method | Data analysis | Research limitation |
|---|---|---|---|---|---|---|---|---|---|
| | | | | | | | | calculate correlations among the variables. p-values less than 0.05 (two-sided) were considered statistically significant, and data were reported with 95% confidence intervals. Hierarchical multiple regression was utilized to predict the likelihood of initiation and sustenance of handwashing behavior based on multiple regressors or independent variables, such as age, gender, race/ethnicity, and individual constructs of the MTM. Gender and race/ethnicity variables were dummy coded. Assumptions of independence of observations, linearity (e.g., scatterplot and partial regression plots), homoscedasticity, multicollinearity, and normality were evaluated on the full model. | |
| Explaining Screen-Time Behavior Among Preschoolers in Northern India Using Multi Theory Model: A Parental Cross-Sectional Survey | Manoj Sharma, Kavita Batra. (2021) .INTERNATIONAL QUARTERLY OF COMMUNITY | America | This study assessed the multi-theory model (MTM)'s applicability in explaining the ST behavior change among preschoolers through parents. | 72 | Screen-Time Behavior Among Preschoolers in Northern India. | A Parental Cross-Sectional Survey | A quota sample of 72 parents was drawn from Northern India. Data were analyzed using multiple regression. | Participants' responses, from Qualtrics, were exported to Microsoft Excel, and then imported to IBM SPSS version 26.0 for the statistical analysis. Descriptive statistics, including means and standard deviations for all continuous variables and frequencies and percentages for categorical variables, were computed. The correlations among the variables were computed using the independent Pearson's correlations test. The two stepwise multiple regression models were fitted with initiation and sustenance constructs as dependent variables. For the initiation model, three predictors, namely participatory dialogue, behavioral | First, the study was conducted in a small geographical area in Northern India, which may limit the generalizability of the results to other populations and regions. Second, the ST behavior was measured through a past 7-day recall, which may introduce a recall bias. Failure to correctly observe the ST behaviors in children, and all the other shortcomings of self-report can also induce bias. Future research can utilize more objective measures of measuring ST behavior such as actual recording on an app or other such means. Third, the study used a cross-sectional design which limits its ability to provide conclusive evidence regarding causality. Future |

| Article Title | Author, year, journal | Country | Research purpose | Participants | Behavior type | Research type | Research method | Data analysis | Research limitation |
|---|---|---|---|---|---|---|---|---|---|
| | HEALTH EDUCATIO N | | | | | | | confidence, and changes in the physical environment, were utilized. In the sustenance model, emotional transformation, practice for change, and changes in the social environment were used as predictors. P-values less than 0.05 (two-sided) were considered statistically significant, and data were also reported as 95% confidence intervals. | studies can utilize longitudinal designs. Fourth, the study did not establish test-retest reliability due to the lack of enough resources. Finally, in the initiation model and sustenance model, the intention for behavior was measured instead of actual behavior which is justified based on previous work with this theory and theory of planned behavior. Prospective studies addressing the above limitations can be designed to strengthen empirical evidence. |
| Explaining the Correlates of Eating Outside-of-Ho me Behavior in a Nationally Representative US Sample Using the Multi-Theory Model of Health Behavior Change: A Cross-Sectional | Manoj Sharma, Bertille Assoumou, and Kavita Batra. (2024). International Journal of Environmen tal Research and Public Health | America | The purpose of this analytical cross-sectional study was to investigate EOH behavior by using the MTM among a nationally representative sample in the United States (US). | 1474 | Eating out behavior in a nationally representative sample of the United States. | A Cross-Sectional Study | Investigation tool for 61 projects based on MTM theoretical framework. | First, the univariate analysis was performed to describe the data and also to identify any patterns in the data. Categorical variables were represented as counts and proportions, whereas continuous variables were reported as means and standard deviations, unless otherwise stated. The box plot was inspected to assess outliers in the data. The assumption of normality was assessed by Shapiro–Wilk's test ($p > 0.05$). Chi-square/Fisher's exact tests were used to compare categorical data, whereas the independent-samples t-test was used to compare the mean scores of the MTM constructs across groups. Pearson correlation analysis was performed for the intercorrelation matrix between the MTM constructs and hierarchical regression models were built to predict the variance in the initiation and sustenance by certain predictor variables beyond demographic characteristics. For the tables | There were some shortcomings of this study. This study utilized a cross-sectional design which, while being quick and inexpensive, failed to establish causality due to a lack of temporality. To address this limitation, future studies must test the MTM for EOH in experimental designs. Further, self-reports including intent for EOH instead of recording actual behavior were used in this study that lend themselves to measurement biases. While for attitudina measures, self-report is the only approach; for actual behaviors, in future experimenta studies, efforts must be made to measure these. Also, the test–retest reliability (stability of the instrument was not tested, which is imperative for future experimental research Next, our current sample size did not allow stratification analysis to unveil some possible dimensions of intersectionality, such as race, age, marital |

| Article Title | Author, year, journal | Country | Research purpose | Participants | Behavior type | Research type | Research method | Data analysis | Research limitation |
|---|---|---|---|---|---|---|---|---|---|
| Study | | | | | | | | more than 2 by 2 in the chi square, the exact p values were calculated by using adjusted residuals. For the independent-samples t-test, Levene's test for equality of variance was conducted to check the assumption of the homogeneity of variance. We also ran hierarchical multiple regression to predict the initiation (continuous variable) by a series of models, including demographic characteristics, and MTM constructs. This was to determine the R-square change and improvement in prediction after adding variables during the model-building process. A similar hierarchical model was built with sustenance as a dependent variable too. Prior to running the regression models, linearity was assessed by partial regression plots and a plot of standardized residuals against the predicted values. The independence of residuals was assessed by the Durbin–Watson statistics. The multicollinearity was assessed by the tolerance and variance inflation factor (VIF). We also performed multinominal logistic regression to model the log odds of the initiation and sustenance levels. There was a total of 5 levels, including "not at all likely (0), "somewhat likely" (1), "moderately likely" (2), "very likely" (3), and "completely likely" (4). We further recoded these levels to a total of three levels by combining "not at all likely" with "somewhat likely" and by | status, education, employment etc., which could be an important recommendation for future studies with a larger sample size. Finally, residual confounding bias could have been introduced due to some variables being left unmeasured, such as ecologically conscious purchase behavior, and opting for healthy food choices were not investigated in this study. |

| Article Title | Author, year, journal | Country | Research purpose | Participants | Behavior type | Research type | Research method | Data analysis | Research limitation |
|---|---|---|---|---|---|---|---|---|---|
| | | | | | | | | merging "very likely" with "completely likely". For the reliability diagnostics, we calculated Cronbach's alpha, as well as McDonald's Omega. The 95% confidence intervals of proportions were calculated by the normal approximation to the binomial calculation. IBM SPSS (V.28) was used to analyze the data, and the level of significance was set at 5%. | |
| Factors associated with initiation and sustenance of stress management behaviors in veterinary students: Testing of Multi-Theory model (MTM) | Vinayak K. Nahar. (2020). INTERNATIONAL JOURNAL OF ENVIRONMENTAL RESEARCH AND PUBLIC HEALTH | America | The purpose of this study was to examine the utility of the multi-theory model (MTM) of health behavior change in predicting the initiation and sustenance of stress management behaviors among veterinary students. | 342 | Stress management behavior of veterinary students. | A cross-sectional study design | A cross-sectional design was used to study the efficacy of the MTM in predicting initiation and sustenance of stress management behaviors among veterinary students at a private College of Veterinary Medicine in the Southeast United States. Researchers collected data using a 54-item valid and reliable survey. Only students who did not already engage in daily stress management behaviors were included in the study. After recruitment and exclusion, a total of 140 students remained and participated in the study. | Descriptive statistics were performed for all variables. The dependent variables, intention to initiate and to sustain relaxation behaviors, were calculated on a continuous scale. To assess statistically significant relationships between demographic covariates and MTM variables of interest, Pearson Product-Moment correlations were performed for continuous variables and independent samples t-tests were performed for categorical variables. Analyses were performed to determine the utility of MTM in predicting intention to both initiate and to sustain relaxation behaviors in two separate models, model 1 and model 2. In model 1, initiation, independent variables were participatory dialogue, behavioral confidence, and changes in the physical environment. In model 2, sustenance, independent variables were emotional transformation, practice for change, and changes in the social environment. For both models, researchers first determined statistically significant | / |

| Article Title | Author, year, journal | Country | Research purpose | Participants | Behavior type | Research type | Research method | Data analysis | Research limitation |
|---|---|---|---|---|---|---|---|---|---|
| | | | | | | | | demographic covariates and entered them into block 1. Hierarchical multiple regression was then performed among the significant covariates and the independent variables for each model. All statistical analyses of data were completed using IBM SPSS statistical software version 25.0 with a significance level of 0.05. | |
| Hesitancy in COVID-19 Vaccine Uptake and Its Correlated Factors Using Multi-Theory Model among Adult Women: A Cross-Sectional Study in Three States of Somalia | Adam A. Mohamed. (2023). VACCINES | Germany | This study aimed to understand the current level of awareness, accessibility, trust, and hesitancy toward the COVID-19 vaccine among women in Somalia. | 999 | Hesitancy to vaccinate adult women against COVID-19. | A cross-sectional study design | To assess COVID-19 vaccine uptake, acceptance, community awareness, and hesitancy rates in Somalia, we carried out a cross-sectional mixed methods study in three regions of Somalia that were selected randomly out of the 18 regions of Somalia. A multi-theory model (MTM) was developed to identify correlated factors associated with the hesitancy or non-hesitancy toward COVID-19 vaccination among women of all ages (18 years and above). | The main dependent variable of the study was willingness to get the vaccine if made available. Respondents were asked if they were willing to receive the COVID-19 vaccination for themselves. The expected response for the willingness question was 'no intention to receive vaccination', 'undecided', or 'intention to receive the vaccination'. The intent to get vaccinated was considered as 'vaccine acceptance' whereas uncertainty and unwillingness to get vaccinated were considered as 'vaccine hesitancy' and 'vaccine unacceptance', respectively. The study also captured the respondents' awareness of the availability of vaccination, knowledge and attitude toward the vaccination, and previous vaccination experience. | / |
| Introspective Meditation before Seeking | Manoj Sharma, Kavita | America | This pilot study focuses on introspective meditation performed before seeking | 9000 | Introspective meditation before seeking pleasurable | A Multi-Theory Model-Based Pilot Study | A non-probability sample of college students was recruited from a midsized Southern University of the | Survey responses were imported to IBM SPSS version 27.0 for the analyses. For assessing the construct validity of the MTM tool, Confirmator factor analysis (CFA) with a | First, the sample size of this study with college students was very small, and the sample was sourced from a single institution. Hence, there is limited generalizability of the |

| Article Title | Author, year, journal | Country | Research purpose | Participants | Behavior type | Research type | Research method | Data analysis | Research limitation |
|---|---|---|---|---|---|---|---|---|---|
| Pleasurable Activities as a Stress Reduction Tool among College Students: A Multi-Theory Model-Based Pilot Study | Batra. (2022). HEALTHCARE | | pleasurable activities, which is a self-reflection about whether to pursue a goal that will bring sensory pleasure in life. | | activities in college students. | | United States using a 52-items web-based survey built in Qualtrics. | maximum likelihood method was used. Reliability diagnostics were assessed to check the internal consistency of the MTM instrument. For establishing a one-factor solution following the Kaiser criterion of Eigenvalue greater than or equal to 1.0, factor loadings on each item greater than 0.610 (after doubling the critical value for a sample size of 65 at an ↵ = 0.01 for a two-tailed test) were established as a priori, given the generally accepted recommendations from previous studies. Categorical variables were represented as frequencies and proportions, whereas continuous variables were represented by mean and standard deviations. Anxiety, depression, and stress were coded per scoring criteria indicated by previous studies. The normal approximation to the binomial distribution method was used to calculate 95% confidence intervals of proportions in the univariate analyses. A bivariate Pearson's correlation test was performed to calculate correlations among the MTM variables. Two models of hierarchical multiple regression (HRM) were fit to predict the likelihood of initiation and sustenance of introspective meditation among students who were not currently practicing it. All assumptions of HRM, including independence of observations, linearity, homoscedasticity, multicollinearity, and normality, were assessed. A detailed | results. Future studies should collect data from multiple institutions with larger sample sizes. Second, the campus racial diversity of the college students could not be captured well due to the majority of participants in the study being White. Hence, the results of this study may not be transferable to other campuses that have broader racial diversity. In addition, some unmeasured variables, such as substance abuse, alcohol consumption, smoking, and major of the students may have introduced some residual confounding. Third, this was a cross-sectional study (snapshot) study at a specific point in the academic year. One of the limitations of using a crosssectional study design is that a temporal causal relationship between the study variables cannot be determined and the relationships are limited to correlations and associations. Fourth, since this was an online survey, it is assumed that the participants completing the survey were truly the participants involved in the study. Additionally, in survey studies using self-report, there is always an element of 'social desirability bias that happens when respondents mark responses which portray a favorable image of themselves. |

| Article Title | Author, year, journal | Country | Research purpose | Participants | Behavior type | Research type | Research method | Data analysis | Research limitation |
|---|---|---|---|---|---|---|---|---|---|
| | | | | | | | | strategy of HRM model building can be seen in Table 1. All p-values are two-sided. | |
| Multi-Theory Model and Predictors of Likelihood of Accepting the Series of HPV Vaccination: A Cross-Sectional Study among Ghanaian Adolescents | Matthew Asare. (2020) .INTERNATIONAL JOURNAL OF ENVIRONMENTAL RESEARCH AND PUBLIC HEALTH | Ghana | We used multi-theory model (MTM)constructs to predict initiation and completion of HPV vaccination series in Ghanaian adolescents. | 285 | Ghanaian teenagers receive the HPV vaccine series. | The cross-sectional study | We used multi-theory model (MTM) constructs to predict initiation and completion of HPV vaccination series in Ghanaian adolescents. Adolescents (n = 285) between the ages of 12 and 17 years old were recruited from four selected schools in Ghana to participate in the cross-sectional study. | Descriptive statistics were calculated on the demographic data to describe the sample. Univariate analysis was used to compare the mean difference of participants' likelihood of getting the first dose and completing the recommended series of HPV vaccination. Hierarchical multiple regression analyses were conducted to predict HPV vaccination behaviors. All data were analyzed using the IBM Statistical Package for Social Sciences (SPSS) version 25.0. | The limitations include the nature of the study design as a cross-sectional study because it can introduce response, recall, and selection biases. Our outcomes are based on self-reported intention to initiate and complete the HPV vaccine series. We were unable to verify vaccination status through vaccine records. The use of a convenience sample limits the generalizability of the study findings to only study participants. The small sample size of 25 adolescent boys is a limitation in the study. A future study should include a larger sample size of boys to better understand the association of sex and HPV vaccination uptake. Finally, the lack of data from parents who are key in decision making for vaccination is a major limitation. However, knowing factors that influence adolescents' likelihood of accepting the HPV vaccination can help guide a future intervention to facilitate adolescents-parent communication about HPV vaccination. Also, predicting HPV vaccination behavior among adolescents is consistent with several studies. |
| Novel behavioral model in | Dhiraj Panjwani. (2022). | India | A novel behavioral model known as the multi-theory model (MTM) was used to | 235 | Brushing behavior of health science students. | A cross-sectional study | Students pursuing Medicine and Dentistry in a University setting were included. A validated | The independent t tests and one-way analysis of variance (ANOVA) were used according to the division of groups to compare data between demographic variables, twice daily | As this is an explorative study, random sampling of subjects was not feasible hence does not define the population in general. Since more than two thirds of the study population |

| Article Title | Author, year, journal | Country | Research purpose | Participants | Behavior type | Research type | Research method | Data analysis | Research limitation |
|---|---|---|---|---|---|---|---|---|---|
| evaluating initiation and sustenance of teeth brushing behavior among students pursuing health sciences: A cross-sectional study | F1000 Research | | understand two important aspects of health behavior change: (i) Initiation and (ii) Sustenance in twice daily teeth brushing in a university setting with objectives to identify factors effecting MTM in initiation and sustenance of twice daily brushing behavior among students pursuing health sciences and correlating the MTM theory with socio-demographic and behavioral patterns. | | | | questionnaire was designed for this study. Questions were framed to evaluate the constructs of initiation and sustenance of MTM, personality, sleeping habits and demographic corelates of participants. | brushing and MTM scores. Pearson product-moment correlation was used to correlate brushing habits, MTM questionnaire scores with Ten Item Personality Inventory and the questionnaire on sleeping habits. A correlation was established between different domains of MTM, Ten Item Personality Inventory, twice daily brushing and demographic criteria. The items which showed statistical significance were then entered into the block for hierarchical multiple regression. A hierarchical linear regression is a form of a multiple linear regression analysis in which multiple variables are added to the model in separate steps. This was done to statistically control variables (Cofounders) (academic class, academic performance, professional fields), to see whether adding variables significantly improves a model's ability to predict the outcome (MTM Model) variable to investigate the model. A stepwise regression was then performed, among the significant covariates and the independent variables for these models. All statistical analyses of data were completed using IBM SPSS statistical software version 21.0 with a significance level <0.05. | were dental students, the results of the study might have been affected. But since this is the first time the MTM has been utilized in the field of dentistry, it was appropriate to have a study sample comprising of future health professionals, to confirm the validity of the instrument in the population. In future studies, a much larger sample size should be employed and interventions can be designed for participants who do not follow brushing twice daily behaviour and are willing to initiate and sustain changed brushing behaviour. Since this is a cross-sectional analysis, the responses of the participants are only pertaining to the week before they answered the questionnaire. |
| Predicting Flossing through the | Manoj Sharma, Kavita | America | This cross-sectional study aimed to conduct an exploratory behavioral research to identify | 520 | Flossing behavior among African American/Black | A cross-sectional study | A 39-item psychometrically valid web-based questionnaire was used to collect responses from a nationwide | The data were analyzed using bivariate and multivariate statistical methods. Of 520 minority adolescents, the proportion of flossing was nearly equally split in the sample. | The data were collected by self-reports, which have the potential for several biases, such as dishonesty, exaggeration, under-reporting, acquiescence bias, recall |

| Article Title | Author, year, journal | Country | Research purpose | Participants | Behavior type | Research type | Research method | Data analysis | Research limitation |
|---|---|---|---|---|---|---|---|---|---|
| Application of the Multi-Theory Model (MTM) of Health Behavior Change among Minority Adolescents in the United States | Batra. (2022). INTERNATIONAL JOURNAL OF ENVIRONMENTAL RESEARCH AND PUBLIC HEALTH | | evidence-based (theory-based) approaches to promote flossing behavior among African American/Black and Latinx/Hispanic (minority) adolescents. | | and Latino/Hispanic (minority) adolescents. | | sample of minority adolescents aged 10–17 years residing in the United States. | A significantly higher proportion of minority adolescents who were flossing had access to floss as opposed to those who were not flossing (86.8% vs. 69.8%, $p < 0.001$). A significantly higher proportion of minority adolescents who were not flossing did not visit the dentist over the past year as opposed to those who floss (25.2% vs. 14.7%, $p < 0.001$). Among the participants who were not flossing, gender, grade level, instruction in school regarding flossing, and multi-theory model (MTM) of health behavior change constructs were the significant predictors ($p < 0.001$) of initiating and sustaining flossing. | bias, and social desirability bias. However, for gauging attitudes, this is the only way to collect data. In this study, intention for flossing was used as a proxy measure of actual flossing. Future studies can utilize experimental designs with interventions to gauge actual behavior change. Further, we did not test for stability reliability in our study. Future studies can conduct the test–retest reliability of the instrument, especially before conducting interventional studies. Finally, the crosssectional study design did not allow for establishing causality, as both the independent and dependent variables were measured at the same point in time. Future work with interventional experimental studies can overcome this limitation. |
| Predicting physical activity behavior in African American females: Using multi theory model | Traci Hayes, Vinayak K. Nahar. (2018). Journal of Research in Health Sciences | America | The objective of this research was to test MTM in its ability to predict physical activity behavior in African American women. | 156 | The physical activity behavior of African American women. | A cross-sectional study | African American women aged 18 yr and older were recruited at various locations (primarily churches) of Jackson, a large city in Central Mississippi instead of southern Mississippi to participate in this cross-sectional study in 2016. The valid and reliable survey was administered to a G*Power calculated quota sample of 156 | The independent variables were the constructs of MTM operationalized as interval/ratio scores and dependent variables were initiation and sustenance of physical activity behavior also operationalized as interval/ratio score. Stepwise multiple regressions were utilized to analyze the survey data. Descriptive statistics were used to analyze the demographic information and describe the data using frequencies and percentages for categorical variables and means and standard deviations for metric variables. All data were analyzed using IBM SPSS. | / |

| Article Title | Author, year, journal | Country | Research purpose | Participants | Behavior type | Research type | Research method | Data analysis | Research limitation |
|---|---|---|---|---|---|---|---|---|---|
| | | | | | | | women either in person or via a Qualtrics link sent through an e-mail. | | |
| Predicting Physical Activity in Chinese Pregnant Women Using Multi-Theory Model: A Cross-Sectional Study | Wei Zhang, Suwen Feng. (2022) .INTERNATIONAL JOURNAL OF ENVIRONMENTAL RESEARCH AND PUBLIC HEALTH | China | This study aimed to: (1) assess the utility of Multi-Theory Model (MTM) to explain the intentions of PA behavior in Chinese pregnant women; (2) analyze the predictors in initiating and maintaining PA behavior based on MTM. | 450 | Physical activity of pregnant women in China. | A Cross-Sectional Study | A cross-sectional study including pregnant women was conducted from March to June 2022 at a university hospital in Hangzhou, Zhejiang Province, China. Participants completed measures that included a self-developed demographic questionnaire and a 29-item MTM questionnaire. | Data analysis was conducted using IBM SPSS Statistics 25 and IBM AMOS 26. Continuous variables are represented by the mean and standard deviation (SD), and categorical variables are described by percentage frequency. The stepwise multiple regression model was used to model the association between outcome and independent variables. The level of statistical significance was $p \leq 0.05$. | First of all, the participants in this study were confined to one hospital in Hangzhou, Zhejiang Province, China, with a small sample size. In the future, participants from different regions and a larger sample size should be adopted. Moreover, the study relied on subjective self-reporting rather than objective measurements of behavior, which can be biased or exaggerated. Meanwhile, in a cross-sectional design study, the independent and dependent variables were collected simultaneously, which cannot determine the temporality of the association. |
| Predictors in Initiating and Maintaining Nutritional Behaviors to Deal With | Nooshin Yoshany. (2022) .International Quarterly of Community | America | This article examines predictors of initiation and maintenance of nutritional behavior in response to menopausal symptoms based on a multi-theoretical model. | 204 | Nutritional behavior to cope with menopausal symptoms. | A Cross-Sectional Study | The participants were required to complete the demographic information questionnaire and a researcher-made questionnaire over the effective nutritional behaviors in menopause based on the MTM. | After entering the data into SPSS 22, the relationships between variables were examined by path analysis model using the AMOS software version 23. Path analysis, an extension of a multiple regression model that allows simultaneous examination of the multiple responses, was used to estimate and evaluate the causal relationships in | / |

| Article Title | Author, year, journal | Country | Research purpose | Participants | Behavior type | Research type | Research method | Data analysis | Research limitation |
|---|---|---|---|---|---|---|---|---|---|
| Menopausal Symptoms Based on Multi-Theory Model | Health Education | | | | | | | terms of the MTM. | |
| Predictors of behaviour change for unhealthy sleep patterns among Indian dental students | Ankur Sharma, Meena Jain. (2021). International Journal of Adolescent Medicine and Health | India | The present study aimed at determining predictors of sleep behaviour change among dental students using the multi-theory model (MTM) of health behaviour change in India. | 535 | Unhealthy sleep pattern behaviors of Indian dental students. | A Cross-Sectional Study | This study was conducted among 535 students of a dental college in India. Predictors of sleep behaviour change were assessed using a validated 30-item questionnaire. Theoretical predictors of sleep behaviour were modelled using multiple linear regression. | Theoretical predictors of sleep behaviour were modelled using multiple linear regression. If the probability of coefficient of regression (F) to enter a predictor in the model was 0.05 or less, it was retained, while if it was 0.10 or more, it was removed. All the data were analysed using SPSS statistical software, version 22 (IBM Corp. Released 2013. IBM SPSS Statistics for Windows, Version 22.0. Armonk, NY, USA). | / |
| Predictors of regular physical activity behavior and quality of life in post-menopausal Iranian | Nooshin Yoshany. (2022). JOURNAL of MEDICINE and LIFE | Iran | This research aims to identify the predictive factors related to the initiation and sustaining of regular physical activity behaviors and their influence in adapting to menopausal symptoms. | 200 | General physical activity behavior and quality of life in postmenopausal Iranian women. | Descriptive cross-sectional research | The study uses the multi-theory model (MTM) as the conceptual framework. The descriptive cross-sectional research was conducted on 200 post-menopausal women aged 45-55 years. All participants were referred to health centers, where they completed a three-part questionnaire involving: | Data were collected, managed, and analyzed using SPSS 20 and AMOS 23 software. | / |

| Article Title | Author, year, journal | Country | Research purpose | Participants | Behavior type | Research type | Research method | Data analysis | Research limitation |
|---|---|---|---|---|---|---|---|---|---|
| women based on the multi-theory model | | | | | | | demographic information, a questionnaire on the influence of regular physical activity on the onset and sustaining of menopause using the MTM, and a standard questionnaire of menopausal quality of life. | | |
| Predictors of Responsible Drinking or Abstinence Among College Students Who Binge Drink: A Multitheory Model Approach | Manoj Sharma. (2018). The Journal of the American Osteopathic Association | America | To use the multitheory model (MTM) of health behavior change to predict initiation and sustenance of responsible drinking or abstinence among binge-drinking college students in a sample drawn from a large southern public university. | 289 | Alcoholic college students rational drinking or abstinence behavior. | A Multitheory Model Approach | This cross-sectional survey study included a sample of college students who binge drank in the past 30 days. A 39-item face- and content-valid instrument was used. | Data were analyzed using IBM SPSS statistical software version 22.0. Descriptive statistics were calculated for all measured variables. Pearson product-moment correlation was used for continuous demographic variables, and analysis of variance was used for categorical demographic variables. Both dependent variables (intention to initiate and sustain responsible drinking/abstinence behaviors) were measured on a continuous scale. Statistical significance level of .05 was set a priori for all analyses. The following analyses were carried out to determine the utility of MTM in predicting intention to initiate and sustain responsible drinking/abstinence behaviors. | The cross-sectional design of this study limits the temporal conclusions that can be drawn from the results. Therefore, it cannot be explicitly said that the MTM constructs precede responsible drinking behavior. However the theories and models that the MTM is derived from indicate that environmental and attitudinal constructs come before health behavior change.34 More robust longitudinal and experimental study designs could be executed in future studies. Other possible limitations were that the majority of the participants were white, and data were collected at 1 southern US university. Thus, caution must be exercised when generalizing these findings to all college students. Furthermore, for the sake of feasibility, proxies for intention and sustenance behavior were measured rather than actual behavior. Future studies could look into finding more objective measures of behavior. Additionally, although |

| Article Title | Author, year, journal | Country | Research purpose | Participants | Behavior type | Research type | Research method | Data analysis | Research limitation |
|---|---|---|---|---|---|---|---|---|---|
| | | | | | | | | | choices were limited for attitudinal assessments, this was a selfreported survey study, and, therefore, measurement bias and false reporting are potential confounders. Stability testing (test-retest) reliability of the survey instrument was not performed—future studies replicating this work should include reliability assessments. |
| Testing Multi-Theory Model (MTM) in Explaining Sunscreen Use among Florida Residents: An Integrative Approach for Sun Protection | Manoj Sharma, Kavita Batra. (2021). HEALTHCARE | America | This cross-sectional study aimed to examine the correlates of initiating and sustaining sunscreen usage behavior among Florida dwellers, using the fourth-generation, multi-theory model (MTM) of behavior change. | / | Florida residents use sunscreen. | A Cross-Sectional Study | A web-based survey containing 51 questions was emailed to Florida residents aged 18 years or above, who were randomly selected from the state voter file. | Participants' responses were first preprocessed and then exported to IBM SPSS version 27.0 (IBM Corp. Armonk, NY, USA) for statistical analyses. Incomplete responses and those with invalid data entries were excluded. Mean and standard deviation were used to represent continuous variables. Counts and proportions were used to express categorical variables. Inferential statistics were conducted through independent samples-t-tests to perform group-wise comparisons. Cronbach's alpha values were computed for the entire scale and subscales to assess the internal consistency. Two hierarchical regression models (HRM) were fit to explain the variance in the likelihood of initiation and sustenance of sunscreen use behavior by MTM individual constructs, besides the demographic variables. Structural equation modeling (SEM) was utilized for the construct validation. The Participants' responses were first preprocessed and then exported to IBM SPSS version 27.0 | First, the use of a cross-sectional design has the limitation of collecting information on independent variables and dependent variables at the same time, thereby precluding causal inferences. Future studies could test the validity of MTM in experimental designs, whereby actual manipulation of the variables is done in a longitudinal manner. Furthermore, self-reports are liable to several shortcomings, such as dishonesty, exaggerated responses, and so on. However, when measuring attitudes, one cannot choose another approach and these are indeed the only means. Finally, the study was done in Florida, thereby limiting the generalizability to other parts of the country. |

| Article Title | Author, year, journal | Country | Research purpose | Participants | Behavior type | Research type | Research method | Data analysis | Research limitation |
|---|---|---|---|---|---|---|---|---|---|
| | | | | | | | | (IBM Corp. Armonk, NY, USA) for statistical analyses. Incomplete responses and those with invalid data entries were excluded. Mean and standard deviation were used to represent continuous variables. Counts and proportions were used to express categorical variables. Inferential statistics were conducted through independent samples-t-tests to perform group-wise comparisons. Cronbach's alpha values were computed for the entire scale and subscales to assess the internal consistency. Two hierarchical regression models (HRM) were fit to explain the variance in the likelihood of initiation and sustenance of sunscreen use behavior by MTM individual constructs, besides the demographic variables. Structural equation modeling (SEM) was utilized for the construct validation. The Analysis of Moment Structure, AMOS (Chicago, IL, USA) was used for SEM. We used indices such as chi-square ("2), root mean square error of approximation (RMSEA), comparative fit index (CFI), and Tucker–Lewis (TLI) to assess how well our models fit the data. Models were considered to have adequate fit if they met the less stringent, but traditionally accepted, values of 0.90 or greater for CFI and TLI, and values less than 0.08 for RMSEA. P-values less than 0.05 were considered statistically significant. | |

| Article Title | Author, year, journal | Country | Research purpose | Participants | Behavior type | Research type | Research method | Data analysis | Research limitation |
|---|---|---|---|---|---|---|---|---|---|
| Testing multi-theory model (MTM) in predicting initiation and sustenance of physical activity behavior among college students | Vinayak K. Nahar. (2016). HEALTH PROMOTION PERSPECTIVES | America | The purpose of this study was to examine the utility of the newly proposed multi-theory model (MTM) of health behavior change in predicting initiation and sustenance of PA among college students. | 495 | College students' physical activity behavior. | A Cross-Sectional Study | Using a cross-sectional design, a valid and reliable survey was administered in October 2015 electronically to students enrolled at a large Southern US University. The internal consistency Cronbach alphas of the subscales were acceptable (0.65-0.92). Only those who did not engage in more than 150 minutes of moderate to vigorous intensity aerobic PA during the past week were included in this study | Descriptive statistical analyses were conducted to describe the study variables. Using stepwise multiple regression, best possible predictors of PA behavior change (i.e., initiation and sustenance) were assessed while controlling for demographic variables. For stepwise multiple regression the a priori criteria of probability of F to enter the predictor in the model was chosen as less than or equal to 0.05 and for removing the predictor as greater than or equal to 0.10. All data analyses were performed using IBM SPSS (version 20.0). | First, the study utilized a cross sectional study design which looks at all the variables at one time thereby nothing can be said about the temporal association of variables. Or in other words strictly speaking we cannot say that the constructs come before the behavior. However, previous theories have indicated that the attitudinal and environmental constructs precede the behavior so we can also assume the same for PA behavior in college students. Second, the actual behavior has not been measured by this study but a proxy intention for initiation and sustenance of behavior has been used in measurement which is subject to criticism. However, there is evidence in previous theories, particularly theory of reasoned action and theory of planned behavior that intentions precede behavior. So the measurement of behavior the way it has been done in this study can be justified. Third, the instrument was all self-report and that too introduces measurement bias. Self-reports are prone to dishonesty, false reporting under reporting or extreme reporting and such biases. However, when it comes to attitudinal assessments there are no other choices, so this limitation must be considered in that context. Finally, the test-retest (stability) reliability of the instrument was not conducted. |

| Article Title | Author, year, journal | Country | Research purpose | Participants | Behavior type | Research type | Research method | Data analysis | Research limitation |
|---|---|---|---|---|---|---|---|---|---|
| Testing the Multi-Theory Model (MTM) to Predict the Use of New Technology for Social Connectedness in the COVID-19 Pandemic | Manoj Sharma, Kavita Batra. (2021). HEALTHCARE | America | The current study aims to test theory-based determinants in explaining the adoption of new technology in a nationally representative sample during the COVID-19 pandemic. | 382 | The use of new technologies for social linkages in the COVID-19 pandemic. | A Cross-Sectional Study | A psychometrically reliable and valid instrument based on the multi-theory model (MTM) of health behavior change was administered electronically using a cross-sectional study design. | Participants' responses to Qualtrics were exported to a spreadsheet and then imported to IBM SPSS version 27.0 for analysis. Confirmatory factor analysis (CFA) using the extraction method of maximum likelihood was utilized. Reliability diagnostics or Cronbach's alpha was computed for all the subscales. Critical values for determining one-factor solution were set according to the prespecified literature's criteria. The critical value for a correlation coefficient at α = 0.01 for a 2-tailed test for the sample size of 400 participants was 0.129. This was doubled for testing the significance of loading [26]. Hence, a critical value of 0.258 was deemed appropriate. The normality assumption of data was assessed using the Shapiro–Wilk test and normal Q-Q plots. An independent-samples- t-test was utilized to compare the mean scores across new technology users and non-user groups. A chi-square test was conducted to compare categorical variables. A post-hoc contingency table analysis using adjusted residuals (or Z scores) was performed in case of multiple comparisons. Bonferroni corrected p-values were generated. Bootstrapped significance testing for the chi-square test was conducted to examine replicability and consistency. The score of social isolation was dichotomized as low social isolation (≤6.0) and high social isolation (>6.0) | A cross-sectional study limits the establishment of causal inferences due to data on independent correlations and dependent variables being collected simultaneously. Further, reliance on self-reports introduces potential measurement bias. Unmeasured variables may prevent accountability of predictability to close to 100%. Finally, even though we collected data from a nationally representative sample in terms of gender and race, all other variables, including age and region/geographical distribution, could have introduced sampling bias, which limits the generalizability of our findings. Moreover, the sole purpose of this study was model testing and did not determine the prevalence estimates. Moreover, COVID-19 restrictions posed challenges in the sampling. |

| Article Title | Author, year, journal | Country | Research purpose | Participants | Behavior type | Research type | Research method | Data analysis | Research limitation |
|---|---|---|---|---|---|---|---|---|---|
| | | | | | | | | by using the median-split method. Categorical variables were expressed as counts and proportions, whereas continuous variables were represented as means and standard deviations. Two separate Hierarchical Regression Models (HRM) were built to predict the variance in the likelihood of initiation and sustenance of new technology behavior by multiple factors, such as demographic characteristics, social isolation, and MTM constructs. All assumptions of HRM were assessed. The significance level was set at 0.05, and 95% confidence intervals were reported wherever applicable. | |
| Theory-based antecedents of breastfeeding among pregnant women in the United States | Manoj Sharma. (2024). HEALTH PROMOTION PERSPECTIVES | America | The study examined the correlates of intention to breastfeed among US pregnant women based on the multi-theory model (MTM) of health behavior change. | 315 | Pregnant women in the United States breastfeed. | A Cross-Sectional Study | Using a cross-sectional design, a 36-item online survey was administered to a nationally representative sample of 315 pregnant women in the US. The instrument was psychometrically validated for face, content, and construct validity by a panel of six experts over two rounds. Further, construct validation was done by confirmatory factor analysis (CFA). Hierarchical regression modeling was employed to explain the | Descriptive statistics were calculated for demographic variables to characterize the study sample using the MEANS and the FREQ procedures in SAS version 9.4 (SAS Institute Inc., 2016, Cary, NC, USA). The first-order multi-factor structure of the initiation and sustenance models was validated using confirmatory factor analysis (CFA). The CFA was accomplished using the package from R Statistical Software version 4.3.0.29,30 The item responses were treated as ordinal variables in the CFA. We used the weighted least squares with mean and variance adjustments (WLSMV) as the estimator for the CFA to account for the ordinality in participant responses.31 Model fit was diagnosed using the robust estimates of the comparative fit | For example, the study utilized a cross-sectional design which, while fast and inexpensive, collects data on the independent variables and dependent variables at the same time thereby causing issues with temporality. Since the survey implemented was web based, respondents with biases may have been recruited which may affect the generalizability of the results.36 We also collected data via self-reports. We also did not conduct test-retest reliability assessments on our instrument, which is something future researchers should pursue. Finally, the study did not measure the actual behavior and only used intention as a proxy measure of breastfeeding. |

| Article Title | Author, year, journal | Country | Research purpose | Participants | Behavior type | Research type | Research method | Data analysis | Research limitation |
|---|---|---|---|---|---|---|---|---|---|
| | | | | | | | intention to start breastfeeding and sustain exclusive breastfeeding for up to six months and with complementary foods for up to 24 months. | index (CFI), root mean square error of approximation (RMSEA), standardized root mean squared residual (SRMR), and Tucker Lewis index (TLI). We used the following cutoff criteria recommended by Hu and Bentler to assess the acceptability of fit: CFI above 0.95, TLI above 0.90, RMSEA below 0.08, and SRMR below 0.08.31,32 Cronbach's alpha values were measured for the whole scale and each subscale defined by the constructs to assess internal consistency. The lower threshold for acceptable Cronbach's alpha values was set at 0.65.33,34 Confidence intervals for the Cronbach's alpha values were calculated using bootstrapping methods. The loadings were examined for each survey item to test the correlation between item responses and constructs. | |
| Use of the multi-theory model (MTM) in explaining initiation and sustenance of indoor tanning cessation among college | Amanda H. Wilkerson. (2023). ARCHIVES OF DERMATO LOGICAL RESEARC H | America | The purpose of this cross-sectional study was to explore and explain the initiation and sustenance of indoor tanning cessation among college students using the multi-theory model (MTM) of health behavior change. | 254 | Indoor tanning for college students. | A Cross-Sectional Study | Data were collected from 254 college students who reported current indoor tanning use using a validated 46-item survey to assess demographics and the MTM constructs. | Data were analyzed using SPSS Version 23 (IBM) and screened for extreme or missing values. Both the mean and standard deviation for metric variables and the percentages and frequencies for categorical variables were determined using descriptive statistics. Multiple linear regression was used to determine the ability of MTM constructs to predict the initiation and sustenance of indoor tanning cessation. Two regression models were created, one for initiation and one for the sustenance of quitting indoor tanning. The | The study used a cross-sectional research strategy. This research design restricts the interpretation of any cause-and-effect correlations as well as any interpretations of the study variables' time and temporal sequence. Thus, the authors cannot confirm that the MTM constructs assessed in the regression models preceded indoor tanning cessation behavior or intentions. Second, the data collected were self-report and are subject to measurement bias. However, due to the measurement of psychosocial variables |

| Article Title | Author, year, journal | Country | Research purpose | Participants | Behavior type | Research type | Research method | Data analysis | Research limitation |
|---|---|---|---|---|---|---|---|---|---|
| students | | | | | | | | constructs of the MTM health behavior theory served as independent variables: participatory dialogue, behavioral confidence, the physical environment changes for the initiation model, and emotional transformation, social environment, and practice for change for the sustenance model. G*Power (University of Kiel, Germany) was used for the sample size calculation, with an α of 0.05, a power of 0.80, seven predictors for each model, and an estimated effect size of 0.09 (small to medium). The resulting sample size from the power analysis was 167 participants. Core assumptions of multiple regression were not violated. Individual characteristics that exhibited a significant bivariate relationship with criterion variables were controlled for in the regression models. Statistical significance was set a priori at $p<0.05$. | in the present study, the authors were limited to the use of subjective measurement techniques. Third, the final sample of indoor tanners was overwhelmingly White and female and were recruited from two large, public universities in the south-central and southeastern US. The homogeneity in the sample limits the generalizability of the study findings to other gender and racial identities. Additionally, university environments, such as providing access to indoor tanning facilities on campus or through the use of university associated cards and accounts may vary, which further limits the generalizability of the findings. Fourth, actual indoor tanning cessation was not measured in the present study. |
| Using a Multitheory Model to Predict Initiation and Sustenance of Fruit and Vegetable | Manoj Sharma. (2018). The Journal of the American Osteopathic Association | America | To predict change in fruit and vegetable consumption behavior among college students who were not eating the recommended amount of fruits and vegetables using the multitheory model (MTM) of behavior change. | 175 | Fruit and vegetable consumption of college students. | A Cross-Sectional Study | Data were collected from 254 college students who reported current indoor tanning use using a validated 46-item survey to assess demographics and the MTM constructs. | All data were analyzed using SPSS version 23 (IBM). Descriptive statistics for sociodemographic and the MTM constructs were conducted. Frequencies and percentages were calculated for categorical variables, and mean (SD) was calculated for metric variables. Internal consistency of the MTM subscales was measured using Cronbach α, and confirmatory factor analysis was used for construct validation.27,28 The criterion for Cronbach α to be | The design cannot establish a causal relationship between the independent and dependent variables because both are being measured at the same time. Also, this study relied on self-reported data, which are subject to bias. However, many of the constructs of the MTM are attitudinal and can only be measured with self-reported information. The study also used intention of behavior change both for initiation and sustenance of behavior change as proxy measures for actual |

| Article Title | Author, year, journal | Country | Research purpose | Participants | Behavior type | Research type | Research method | Data analysis | Research limitation |
|---|---|---|---|---|---|---|---|---|---|
| Consumption Among College Students | | | | | | | | acceptable was 0.70.27 The criteria for the MTM subscales to be construct valid were eigenvalues greater than 1 and factor loadings for each item greater than 0.32.28 Pearson product moment correlation was performed to assess direction and strength of the association between initiation and sustenance and their corresponding constructs. The core assumptions of multiple regression were checked. After each assumption was tested and no violations were recognized, stepwise multiple regression was carried out to predict initiation and sustenance of behavior change based on their respective constructs. | behavior change. Another limitation of the study is that it did not establish test-retest of the survey. This type of reliability testing is especially important for experimental studies. Additionally, because the purpose of this study was theory testing, and internal validity was of prime concern, random sampling was not done. |
| Using multi theory model (MTM) of health behavior change to explain intention for initiation and sustenance of the consumption of fruits and | Jaelrbreiret L. Williams. (2020). HEALTH PROMOTI ON PERSPECT IVES | America | To predict change in fruit and vegetable consumption behavior among college students who were not eating the recommended amount of fruits and vegetables using the multitheory model (MTM) of behavior change. | 134 | Fruit and vegetable consumption by African Americans in a Mississippi barbershop. | A Cross-Sectional Study | Using a cross-sectional design a valid and reliable paper survey was administered during November and December of 2019. The target population for the study consisted of African American adult men (18 or older) that had not consumed recommended levels of fruits and vegetables within 24 hours of taking the questionnaire. A convenience quota sample of African American men from select barbershops in | The IBM SPSS Statistics, version 26 (Chicago, IL, USA) was used to analyze the data for the study. Frequencies and percentages were reported for categorical variables, and means (standard deviations [SD]) for continuous variables. Stepwise multiple regression for model building was conducted to explain behavior change while controlling for demographic variables. Two separate regression models were built for initiation model and sustenance model. The primary predictive initiation variables were participatory dialogue, behavioral confidence, and changes in the physical environment. The primary predictive sustenanc variables were emotional transformation, practice for change, and | There were some limitations to our study. First, this study utilized a cross-sectional design which cannot establish a causal relationship between the independent and dependent variables as the data was are measured at one time (snapshot). Secondly, this study utilized self-reported data, which may have been subject to recall bias, dishonesty, acquiescence bias and other such shortcomings. In addition, this study utilized the intention of behavior change for initiation and sustenance as substitution measures for actual change. The study also did not establish test-retest reliability of the survey. Lastly, random sampling was not utilized; therefore, the results of this survey are limited by the choice |

| Article Title | Author, year, journal | Country | Research purpose | Participants | Behavior type | Research type | Research method | Data analysis | Research limitation |
|---|---|---|---|---|---|---|---|---|---|
| vegetables among African American men from barbershops in Mississippi | | | | | | | Jackson, Mississippi, were asked to complete the 40-item questionnaire on preventive health screening behavior (n=134). | changes in social environment. The a priori criterion of the predictor to enter into the model was P ≤ 0.05, and for removing, the predictor was P > 0.10, as is the norm in SPSS default. | of the sample, which limit generalizability. |
| Using the Multi-Theory Model (MTM) of Health Behavior Change to Explain the Seeking of Stool-Based Tests for Colorectal Cancer Screening | Manoj Sharma, Christopher Johansen, Kavita Batra. (2023).International Journal of Environmental Research And Public Health | America | This cross-sectional study utilizes the multi-theory model (MTM) of health behavior change to explain the seeking of stool-based tests for colorectal cancer (CRC) screening. | 640 | Seek a stool based test for colorectal cancer screening. | A Cross-Sectional Study | An online 57-item questionnaire with an established psychometric validity was used to collect responses from the US-based sample (n = 640) of adults aged 45–75 years old. | Univariate, bivariate, and multivariate statistical tests were used to analyze the data. Frequencies/proportions were used to represent categorical variables, whereas continuous variables were represented as the means and standard deviations given the normal distribution. For initiation, two separate models of hierarchical multiple regression analyses were used among those participants who underwent stool-based colorectal screening and those who had not undergone these screening tests. The selection of variables to be entered in the regression was assessed based on a theoretical, conceptual, and statistical basis. All tests were performed using IBM SPSS (version 28.0), with the level of significance set at 5%. In calculating the sample size, we used the following parameters: confidence level: 95; margin of error: 5%; and population proportion: 33%. The population proportion of (100 − 67) = 33% was based on the data reported by the Centers for Disease Control and | First, we did not analyze the data pertaining to visual (structural) tests such as colonoscopy, virtual colonoscopy, or flexible sigmoidoscopy. Second, we used a cross-sectional study design, which gave us quick results but precludes us from making causal inferences due to a lack of established temporality. Third, we relied on self-reports that are amenable to several biases. However, our study was primarily based on identifying attitudes, and there could be no other means by which to do so. Next, as part of the contractual agreement with Qualtrics, partial responses were not provided, which limited our ability to analyze the characteristics of the non-responders, which is important for understanding the sources of non-response bias. Finally, while we established the face content and construct validity of our instrument, along with the internal consistency reliability, we could not establish test–retest reliability. This is something future researchers should attempt before |

| Article Title | Author, year, journal | Country | Research purpose | Participants | Behavior type | Research type | Research method | Data analysis | Research limitation |
|---|---|---|---|---|---|---|---|---|---|
| | | | | | | | | Prevention in 2018, according to which 67% of US adults aged 50–75 years met the recommendations of colorectal cancer screening. The minimal sample required was 340, and after applying a 20% non-response rate, a total of n = 340 + 68 = 408 was needed. Our sample size is sufficiently larger than estimated, which allowed us to conduct structural equation modeling too. | embarking on intervention research. |
| Using the Multi-Theory Model (MTM) of Health Behavior Change to Explain Intentional Outdoor Nature Contact Behavior among College Students | Manoj Sharma, Vinayak K. Nahar. (2020).INTERNATIONAL JOURNAL OF ENVIRONMENTAL RESEARCH AND PUBLIC HEALTH | America | The purpose of this study was to use the multi-theory model (MTM) of health behavior change, a contemporary fourth-generation behavioral theory in explaining intentional outdoor nature contact behavior among college students. | 401 | College students' outdoor natural contact behavior. | A Cross-Sectional Study | Using a cross-sectional design, 401 students completed the validated survey based on MTM. Of these, 281 met the inclusion criteria. | Data were analyzed for descriptive and inferential statistics using SPSS version 25. For descriptive statistics, frequencies and percentages were calculated for all the demographics and study variables that were categorical. Means and standard deviations were reported for those variables that were continuous. To establish the internal consistency of the scale, Cronbach’s alphas were calculated for each subscale and the entire scale. For construct validation of the scale, a confirmatory factor analysis using the maximum likelihood method was employed. Stepwise multiple regression modeling was performed for both the initiation model consisting of participatory dialogue, behavioral confidence, and changes in the physical environment as predictors; and for the sustenance model with emotional transformation, practice for change, and changes in the social environment as predictors. | First, the study used a cross-sectional design, which is a snapshot in time. The weakness of this design is the lack of ability to establish a temporal relationship, which, though vital for causal inference, has been recently debated. We chose this study design for practical reasons and future studies can use more robust longitudinal designs. Second, the study relied on self-reports which can lead to measurement error in the form of dishonest responses, bias, exaggeration, etc. While attitudinal variables can only be measured through self-report, the actual performance of the behavior can be objectively observed but is usually not feasible as was the case in this study. Future studies must attempt to measure actual behavior objectively. Third, the study used intentional outdoor nature contact behavior for both initiation and sustenance models which is only a proxy of the actual behavior. Finally, the study did not establish |

| Article Title | Author, year, journal | Country | Research purpose | Participants | Behavior type | Research type | Research method | Data analysis | Research limitation |
|---|---|---|---|---|---|---|---|---|---|
| | | | | | | | | | the test-retest (stability) reliability of the instrument used. This was due to lack of adequate resources. Future studies must attempt to do that. |
| Using the Multi-Theory Model (MTM) of Health Behavior Change to Explain Yoga Practice | Traci Hayes, Manoj Sharma. (2022).ALTERNATIVE THERAPIES IN HEALTH AND MEDICINE | America | The purpose of this study was to utilize the fourth-generation, multi-theory model (MTM) of health behavior change to explain change regarding yoga practice of asanas, shava asana, pranayama, dhyana, yama and niyama among college students. | 70 | Yoga Practice. | A Cross-Sectional Study | This cross-sectional study relied on a quota sample of students 18 years and older attending Jackson State University, a historically black college in Jackson, Mississippi, United States. | The descriptive statistics for all study variables are presented in Tables 1 and 2 and demographic variables are shown in Tables 3 and 4. For stepwise multiple regression analysis, the a priori probability levels for F to include the predictor in the model and F to remove the predictor from the model were chosen as ≤0.05 and ≥0.10, respectively. A stepwise multiple regression model was used to analyze the survey data for identifying the best possible predictors of yoga practice (i.e., initiation and maintenance) and is presented in Tables 3 and 4. A statistical significance level of 0.05 was set a priori. Data analyses were completed using IBM SPSS (version 25.0). | Study limitations include its small sample size. The low Cronbach's alpha for the disadvantages of participatory dialogue suggests that a weak relationship exists between the targeted variables that is affected by random factors, and the five questions may not represent challenges faced by yoga practitioners. The study also relied on self-reported data, which is subject to bias or false or exaggerated responses and the cross-sectional design of this study does not establish causality or directionality between MTM variables and yoga practice. Despite multiple targeted communications and previous studies utilizing similar approaches, survey participation was low, which may indicate that the topic may be of limited interest to this population. |
| Using the Multi-Theory Model (MTM) of Health Behavior Change to | Manoj Sharma, Kavita Batra. (2021). pharmacy | America | This cross-sectional study utilized the fourth-generation, multi-theory model (MTM) of health behavior change to explain the correlates of mammography screening among | 374 | Mammography for Asian American women. | A Cross-Sectional Study | A 44-item instrument was evaluated for face, content, and construct validity (using structural equation modeling) and reliability (Cronbach's alpha) and administered electronically to a nationally | We used IBM SPSS version 27.0, plus 7.11, and G* Power software packages for the analyses. Minimum sample size was estimated using the Cohen's effect sizes conventions (based on the type of statistical test) corresponding to 99% power. The significance level was set at 0.05, and 95% confidence intervals were reported wherever appropriate. | / |

| Article Title | Author, year, journal | Country | Research purpose | Participants | Behavior type | Research type | Research method | Data analysis | Research limitation |
|---|---|---|---|---|---|---|---|---|---|
| Explain the Correlates of Mammography Screening among Asian American Women | | | Asian American women between the ages of 45–54 years. | | | | representative sample of Asian American women (n = 374). | Data normality assumptions were assessed through visual inspection of normal Q-Q plots and histograms. An independent-samples t-test was utilized to compare the mean scores of MTM constructs across groups who have had mammography and those who have not. Categorical variables were expressed as counts and proportions, whereas continuous variables were represented as means and standard deviations. Two separate Hierarchical Regression Models (HRM) were built to predict the variance in the likelihood of initiation and sustenance of mammography behavior by multiple factors, such as demographic characteristics and MTM constructs. For construct validation, Structural Equation Modelling (SEM) was utilized to determine the structural relationship between measured variables and latent constructs. Weighted least squares approach (WLSMV estimator) and the 0.05 alpha level were used to test our two hypothesized structural models among the samples. | |
| Using the multi-theory model of health behavior change to identify | Edith Claros. (2020). Journal of Substance Use | America | This study utilized the Multi-Theory Model (MTM) of Health Behavior Change to identify correlates of change in substance use behavior. | 93 | Substance use behavior in a sample of mental health clinics. | A Cross-Sectional Study | Data were collected, using a cross-sectional design, from 93 participants who completed treatment at a substance use treatment facility. Participants completed a 40-item, newly | Descriptive statistics were performed to describe the study variables. We conducted stepwise multiple regressions to determine correlates of change in substance use behavior. To assess demographic covariates (i.e. age, gender, race/ethnicity, education, work status, and yearly household income), Pearson product-moment correlations, independent | Our cross-sectional design provides a snapshot of the variables at one point in time, thus limiting the ability to judge any temporal association between the variables. We cannot argue that the MTM constructs precede the behavior change in SUD. Additionally, we utilized a self-report questionnaire, which is subject to self-favoring by the |

| Article Title | Author, year, journal | Country | Research purpose | Participants | Behavior type | Research type | Research method | Data analysis | Research limitation |
|---|---|---|---|---|---|---|---|---|---|
| correlates of change in substance use behavior in a mental health clinic-based sample | | | | | | | developed, selfadministered questionnaire, grounded on relevant substance use, treatment, and health-related behavior. Cronbach's alpha of all subscales were over 0.70 and deemed acceptable. | t-tests, and analysis of variance were performed, as appropriate. Only age was found to be statistically significantly related with the intent for initiation (r = 0.209; p = .045); hence, it was included as a covariate in the stepwise multiple regression analysis for initiation model. For stepwise multiple regressions, we chose ≤0.05 as a criterion of the probability of F to enter the independent variable in the model and we chose ≥0.10 as a criterion of the probability of F to remove the independent variable from the model. Listwise deletion for missing values was used for the regression analysis. All statistics were conducted using IBM SPSS (Version 23.0). | participants. The study included a relatively small sample size and a set of participants who were relatively homogenous, with the majority being Caucasian males receiving treatment at a private treatment facility. It would be important to expand this study to other levels of care including public treatment centers. Due to the transient nature of individuals in terms of a revolving door of episodic treatment within multiple treatment centers, the outcomes of those subjects who participated in the study remain unknown. Longitudinal studies to address the efficacy of the MTM as well as answering many other fundamental questions regarding SUD treatment and recovery are necessary. |
| Utility of Multi-Theory Model (MTM) to Explain the Intention for PAP Adherence in Newly Diagnosed Sleep Apnea | Manoj Sharma. (2021). Nature and Science of Sleep | America | The objective of this study was to assess the utility of a fourth-generation multitheory model (MTM) in explaining the intention for initiation and sustenance of PAP adherence among newly diagnosed sleep apnea patients. | 138 | Patients with sleep apnea insist on PAP therapy. | A Cross-Sectional Study | For this cross-sectional study, data were collected at a private sleep center located in the Southeastern United States. A total of 138 newly diagnosed patients with sleep apnea who had been prescribed PAP therapy completed a valid and reliable 41- item MTM instrument. Stepwise multiple regression modeling was conducted to assess | All data were analyzed using SPSS, Version 25.0. Descriptive statistics for metric variables were summarized by calculating means and standard deviations while frequencies and percentages were reported for categorical variables. To assess statistically significant associations between covariates (age, gender, race ethnicity, education, work status, income, and sleep study experience) and dependent variables (intention for initiation of PAP therapy and intention for the sustenance of PAP therapy), Pearson Product-Moment correlations, one-way analysis of variance | First, as mentioned earlier, the cross-sectional design used in this study was a snapshot in time where information on both the independent and dependent variables was col lected at the same time. Hence, the temporality of association cannot be established by this study. However, it is worth noting that MTM has been used in a longitudinal randomized controlled trial and the directionality of association of its constructs has been found to withstand the empirical testing.27 The other limitation of the study was that it relied on subjective self-reporting and used a proxy measure of actual |

| Article Title | Author, year, journal | Country | Research purpose | Participants | Behavior type | Research type | Research method | Data analysis | Research limitation |
| --- | --- | --- | --- | --- | --- | --- | --- | --- | --- |
| Patients | | | | | | | MTM based explanatory variables of PAP adherence in this study sample. | (ANOVA), and independent samples t tests were performed. Sleep study experience showed statistically significant associations with both the dependent variables. As per the SPSS default, the a priori criteria in stepwise regression modeling of the probability of the F to enter the independent variable in the model was chosen as ≤ to 0.05 and for removing the independent variable as greater than and equal to 0.10. For both the initiation model and sustenance model, sleep study experience was used as a covariate. | compliance in the form of asking the respondents about their intent to initiate and sustain usage instead of actual usage. Since this study was about psychosocial attitudes and objective measurement is not possible so this was considered as the best possible approach. Another limitation was that the instrumentation used in the study had acceptable face validity, content validity, construct validity and internal consistency reliability but stability was not established. Furthermore, no attempt was made to collect information about different types of PAPs used by the participants, which might have affected the findings of this study, if the type of PAP was included in the analyses as a covariate. Finally, even though we did power analysis, the sample size was relatively small which limits generalizability of the results. |
| Utilizing Multi-Theory Model in Determining Intentions to Smoking Cessation Among | Vinayak K Nahar. (2019). TOBACCO USE INSIGHTS | America | The purpose of this study was to predict initiation and sustenance of smoking cessation among smokers using a fourth-generation behavioral framework, multi-theory model (MTM) of health behavior change. | 148 | Smokers' willingness to quit. | A Cross-Sectional Study | A convenience sample of smokers from a shopping mall in rural, Appalachian Kentucky county was invited to participate in this cross-sectional study. A 38-item, face and content validated, MTM-based survey instrument was administered to the participants. | Descriptive statistics were calculated for all study variables. To assess the relationship between demographic variables (covariates) and dependent variables (initiation and sustenance of smoking cessation behaviors), we performed Pearson’s r, independent samples t-tests, and one way analysis of variance (ANOVA) analyses, as appropriate. Since there were two dependent variables in the current study, two hierarchical multiple regression models were | First, the utilization of a crosssectional research design limits any interpretations about the temporal sequence or time sequence of the study variables. Thus, it cannot be stated that the constructs assessed in both MTM models preceded smoking cessation behavior. However, behavioral theory suggests that psychosocial constructs precede behavior, so when interpreting the study findings, we can assume that the constructs precede smoking cessation |

| Article Title | Author, year, journal | Country | Research purpose | Participants | Behavior type | Research type | Research method | Data analysis | Research limitation |
|---|---|---|---|---|---|---|---|---|---|
| Smokers | | | | | | | | conducted in two blocks to determine the predictive ability of the MTM constructs beyond the influence of the demographic variables. In block 1 for each model, only those demographic variables were entered which were statistically significant in bivariate analysis. In block 2 for each model, all of the MTM constructs were added. A statistical significance level of 0.05 was set a priori. Data analyses were completed using IBM SPSS (version 23.0). | behavior. Second, the study relied on self-report data, which are subject to measurement bias. However, due to the assessment of attitudes and perceptions, the researchers were limited to the measurement approaches available for this study. Third, actual smoking cessation behavior was not assessed as part of this cross-sectional study. Finally, the sample ascertained for this study was a convenience sample of visitors at a shopping mall in a rural, Appalachian Kentucky county, so strictly speaking the findings cannot be generalized beyond the study sample. |
| Application of the multi-theory model to explain veterinarians' intentions to use telehealth/tele medicine | Julia Wells. (2023). VETERINA RY RECORD | America | The goal of this study was to evaluate how effective the multi-theory model (MTM) of health behaviour change explains the initiation and sustenance of telehealth use among veterinarians. | 243 | Veterinarians use tele-health/telemed icine. | A Cross-Sectional Study | Tools based on MTM theoretical framework including 42 projects. | All collected data were analysed using IBM SPSS statistical software (version 22.0) with a significance level of 0.05 set a priori. The dependent variables (i.e., intentional use of telehealth in the near future and sustenance of use from now on) were measured on a continuous scale. The constructs of MTM (i.e., independent variables) were also measured on a continuous scale. The demographics and practice information (i.e., independent variables) were measured on a categorical scale. Descriptive statistical analyses (i.e., mean, standard deviation, frequency, percentage and range) were conducted on all measured variables. Independent samples t-tests were conducted to assess differences in mean between users and non-users of telehealth. In order to evaluate the relationships | The authors agree that there are some limitations to the study that should be addressed. This study was designed as a cross-sectional study; thus, researchers cannot evaluate results over time. Due to a smaller sample size (n = 243), the results of this study should not be extrapolated to the entire veterinary community. As described in section 'Methods', data were collected via a survey sent to participants. With surveys, self-reporting bias should always be considered. Due to the longer nature of the survey, participants may have become tired and started to lose focus, resulting in less accurate responses. Finally, data collection occurred in July 2020, which was in the middle of a global pandemic, in which telehealth had increased in many professions. |

| Article Title | Author, year, journal | Country | Research purpose | Participants | Behavior type | Research type | Research method | Data analysis | Research limitation |
|---|---|---|---|---|---|---|---|---|---|
| | | | | | | | | between the dependent variables and the MTM components, Pearson's product–moment correlations were performed. The following analyses were conducted to determine the utility of MTM in explaining intention to initiate and sustain of telehealth practice among veterinarians in two individual models. | Therefore, study results may not be applicable to normal, non pandemic times. |
| Why people follow a gluten-free diet? An application of health behaviour models | Vilma Xhakollari. (2021).Appetite | Britain | To understand factors affecting adherence to GFD by celiac and non-celiac people through the application of behavioural theories, Integrative Model (IM) and Multi Theory Model (MTM). | 308 | Gluten-free diet. | A Cross-Sectional Study | Analyses were conducted for a sample of 308 subjects, majority females, celiac and non-celiac. Adherence to GFD was measured considering two scales, self-declared adherence and scored adherence, in order to discern possible inconsistencies between what subjects believe and what they really do. Subsequently, adherence to GFD was modelled by considering constructs of MTM and IM. | Data were analysed using R Core Team (2019) 4.0.3. Firstly, descriptive statistics allowed to understand the general profile of the participants. Secondly, correlation tests were applied to understand if the model's constructs were associated with each other. An Ordered logit (OL) model was used to test the IM and MTM theoretical models using the survey data. The OL statistical model was chosen because of the type of dependent variable (adherence to GFD), measured using an ordinal scale, and the assumed relationship between the dependent and independent variables. | Despite its contribution, this research presents limitations in terms of sample representativeness and measurement of adherence to GFD and DASS. Most of the participants in the study are females. Nevertheless, according to many studies, females are mostly affected by CD and concerned about body shape and diets . Furthermore, the authors could not report a response rate since they do not have an instrument for measuring the number of people who saw the post on social media and the number of people who received the leaflets. However, in total, 535 people started the ques tionnaire, and 308 completed it. The study measured the reported adherence to GFD similarly to other studies by considering two measurements: by asking participants directly and indirectly by scoring their adherence to GFD through a scale used in other studies in Italy. Some studies, though, have measured adherence through clinical analysis, and this may be a viable alternative for future research |

| Article Title | Author, year, journal | Country | Research purpose | Participants | Behavior type | Research type | Research method | Data analysis | Research limitation |
|---|---|---|---|---|---|---|---|---|---|
| | | | | | | | | | conducted by researchers with the appropriate background. Finally, the DASS applied in this study is a reduction version of the original one. |
| Investigation of intention to initiate and maintain bedside elevation behavior among nurses in neurological intensive care unit | Li S S, Wu J J. (2021). Journal of Nursing Science | China | To investigate the intention to initiate and maintain the bedside elevation behavior of nurses in the neurological intensive care unit, in order to provide reference for the clinical development of behavioral intervention pathways. | 716 | Bedside elevation behavior of nurses in the neurological intensive care unit. | A Cross-Sectional Study | A total of 716 NICU nurses from 11 3A general hospitals in Chongqing were recruited by convenient sampling method， then they were investigated using a self-made questionnaire of the initiation and sustenance intention on the head-of-bed elevation behavior and the general information questionnaire. | SPSS23.0 software was used for descriptive statistical analysis of the data. The counting data were expressed by frequency and percentage, and the measurement data were expressed by mean ± standard deviation. | / |
| Evaluation scale of postural support behavior for preterm infants by nurses in neonatology department and | Meng J W. (2024). Nursing Research | China | To develop the postural support behavior assessment scale for neonatal nurses in preterm infants and verify its reliability and validity. | 203 | Neonatal nurse postural support behavior for preterm infants. | A Cross-Sectional Study | Guided by the multi-theoretical model of health behavior change, the draft of postural support behavior assessment scale for neonatal nurses was developed through literature review, the draft items were revised through expert group discussion, and the postural support behavior assessment scale for neonatal nurses | SPSS 26.0 statistical software was used for data analysis. Quantitative data conforming to normal distribution was expressed as mean standard deviation (x s), and qualitative data was expressed as frequency or percentage (%). The effective recovery rate was used to indicate the enthusiasm of experts. Expert authority is represented by expert authority coefficient (Cr), which is the arithmetic average of expert familiarity coefficient and judgment basis coefficient. The critical ratio method and correlation coefficient method were | At present, there is no standard for measuring postural support behavior of preterm infants for nurses, so this study could not test the calibration validity. In addition, this study only verified the reliability and validity of the scale, and did not explore the status quo of nurses' postures support behavior for preterm infants and related influencing factors. In the follow-up study, a large sample survey is needed to understand the status quo of nurses' posture support behavior for preterm infants, formulate targeted intervention |

| Article Title | Author, year, journal | Country | Research purpose | Participants | Behavior type | Research type | Research method | Data analysis | Research limitation |
|---|---|---|---|---|---|---|---|---|---|
| its reliability and validity | | | | | | | for preterm infants was developed through expert letter consultation and pre-investigation. Convenience sampling method was used to evaluate 203 clinical nurses in neonatal intensive care unit (NICU) and neonatology ward to test the reliability and validity of the scale. | used to screen the items. The content validity index (scale level CVI,S-CVI) was used to evaluate the content validity. Exploratory factor analysis was used to evaluate the structural validity. Cronbach's α coefficient, broken half reliability and retest reliability were used to evaluate reliability. | measures and health education programs, and promote clinical nurses to start and continue posture support for preterm infants. |
| Conceptualization of college students' COVID-19 related mask-wearing behaviors using the Multi-Theory Model of health behavior change | Robert E. Davis. (2021). HEALTH PROMOTION PERSPECTIVES | America | This study aimed to conceptualize mask-wearing behavior among students using the Multi-theory Model (MTM) of behavior change. | 28000 | College students' COVID-19 related mask-wearing behaviors. | A Cross-Sectional Study | In October 2020, students (n = 595) enrolled in a large public southeastern US university were recruited to participate in a cross-sectional survey, using a valid and reliable instrument. Univariate, bivariate, and multivariate techniques described mask-wearing behavior and differentiated theoretical drivers of mask-wearing between individuals compliant and noncompliant with guidelines. | Data analyses for the current study were conducted using IBM SPSS Statistics version 24.0 (IBM Corp. Armonk, NY, USA). Prior to analysis, participants exhibiting large amounts of missing data (i.e. those who provided ≤ the initial demographic items of the survey instrument) were removed (n=64). Subsequent missing data was handled using listwise deletion. For comparative purposes, the sample was split into those in compliance with mask-wearing guidelines, and those reporting non-compliance. Univariate statistics were calculated to reflect characteristics of the study sample as well as descriptors for MTM variables. Correlational analysis was used to examine bivariate relationships between MTM study variables. Additionally, Welch's t tests were used to detect statistically significant differences in MTM variables between those adhering to guidelines and those who were | / |

| Article Title | Author, year, journal | Country | Research purpose | Participants | Behavior type | Research type | Research method | Data analysis | Research limitation |
|---|---|---|---|---|---|---|---|---|---|
| | | | | | | | | not. Because of the small number of participants reporting non-adherence (4.5% of total sample), bootstrapping consisting of 1000 random samples with replacement was used for point estimation. Finally, multiple regression modeling was used to explain initiation and sustenance of mask-wearing among those complying with guidelines. Using G*Power version 3.1, a power analysis was conducted to determine the simple size required to conduct multiple regression modeling. Alpha was set at 0.05, power at 0.80, predictors set at 6, with effect size of 0.15 (medium). The MTM assumes 3 constructs as predictors of both initiation and sustenance models. For power analyses, 6 predictors were included to account for potential addition of covariates. Results of the power analysis dictated a minimum sample of 98, which we increased by 10% (to 108 minimum) to account for potential incomplete data. Demographic covariates were not included within regression models due to their lack of significant bivariate relationship with outcome variables. Similar modeling was not conducted among those exhibiting non-compliance with guidelines due to sample size restrictions. | |
| A Theory-Based | Manoj Sharma, | America | The current study aims to determine recent trends in | 428 | Unvaccinated. | A Recent Evidence | A sample of 428 unvaccinated African Americans were recruited | All analyses were conducted using IBM SPSS v.26,and SAS 9.3.Descriptive and exploratory analyses were utilized to | / |

| Article Title | Author, year, journal | Country | Research purpose | Participants | Behavior type | Research type | Research method | Data analysis | Research limitation |
|---|---|---|---|---|---|---|---|---|---|
| Analysis of COVID-19 Vaccine Hesitancy among African Americans in the United States: A Recent Evidence | Kavita Batra. (2021). HEALTHCARE | | COVID-19 vaccination rates and to test the MTM model in predicting the initiation of COVID-19 vaccines among vaccine-hesitant Blacks. | | | | through a web-based survey using a 28-item psychometric valid questionnaire. | investigate data distribution, normality, missing values, and outliers. All assumptions of the statistical tests were assessed. Categorical variables were compared among vaccine hesitant and non-hesitant groups by Chi-square analyses. The follow-up contingency table analysis(post-hoc) was conducted to obtain p-values corresponding to multi level variables. The observed p-values were Bonferroni-corrected in multiple comparisons to prevent type 1 errors. The values of adjusted residuals (or Z scores)were used to generate Bonferroni-corrected p values. Effect sizes were reported wherever appropriate. Continuous variables, such as age, advantages, disadvantages, participatory dialogue, behavior confidence, and changes in the physical environment were compared among groups using independent-samples t-tests or Welch's t test where equal variance could not be assumed. A square root transformation was applied to the non-normally distributed variables, which were later back-transformed for the ease of interpretation. A bivariate Pearson's correlation was also conducted to investigate the relationships between the MTM constructs. We also calculated the proportion of perceived advantages and disadvantages among hesitant and non-hesitant group. Hierarchical regression modelling was performed to | |

| Article Title | Author, year, journal | Country | Research purpose | Participants | Behavior type | Research type | Research method | Data analysis | Research limitation |
|---|---|---|---|---|---|---|---|---|---|
| | | | | | | | | determine the increment in variation (by R-square change)accounted for through addition of predictors over a set of models. | |
| Examining the Gambling Behavior of University Students: A Cross-Sectional Survey Applying the Multi-Theory Model (MTM) of Health Behavior Change in a Single Institution | Sidath Kapukotuwa .(2023).HEALTHCARE | America | This cross-sectional study aimed to identify and explain the initiation and sustenance of quitting gambling among university students who had participated in gambling during the past month and those who had not using a novel fourth-generation multi-theory model (MTM) of health behavior change. | 1474 | Gambling Behavior of University Students. | A Cross-Sectional Study | Data were collected from a sample of 1474 university students at a large southwestern university in the U.S. between January 2023 and February 2023, utilizing a validated 39-item survey | All data were analyzed using SAS version 9.4 and R Statistical Software version 4.3.0. R: A language and environment for statistical computing. R Foundation for Statistical Computing, Vienna, Austria.). A first-order, multi-factor model was used in the confirmatory factor analysis for initiation and sustenance models. The confirmatory factor analysis (CFA) for construct validation was performed using the R package lavaan. We used Weighted Least Squares with Mean and Variance adjustments (WLSMV) designed for ordinal data in implementing the CFA. The robust estimates of the comparative fit index (CFI), root mean square error of approximation (RMSEA), and the standardized root mean square residual (SRMR) were used to diagnose the fit for the presented model. We used the following cutoff criteria recommended by Hu and Bentler (1999) to assess the acceptable fit: CFI values above 0.95, RMSEA values below 0.06, and SRMR values below 0.08. The internal consistency of the subscales and the entire scale was tested using Cronbach's alpha values, using 0.70 as the lower threshold | Firstly, it was conducted solely at one large university in the southwestern region of the United States. Consequently, the generalizability of the findings to other universities or populations may be limited. Secondly, the study's reliance on self-reported information is a limitation, as it introduces potential biases such as recall bias, dishonesty, and acquiescence bias. Participants may have difficulty accurately recalling their gambling behavior or may provide socially desirable responses. However, it is important to note that self-reported data remain the primary method for collecting information on attitudes and behaviors related to health behavior. Thirdly, the lack of test–retest reliability assessment prevented us from examining the instrument's consistency over time. However, this limitation also presents an opportunity for future research to explore the stability of the instrument by conducting test–retest studies. Fourthly, we asked the students about their past 30-day behavior, which is not necessarily representative of typical behavior. Fifthly, we used an incentive through participation in a random number draw, which has the potential of being |

| Article Title | Author, year, journal | Country | Research purpose | Participants | Behavior type | Research type | Research method | Data analysis | Research limitation |
|---|---|---|---|---|---|---|---|---|---|
| | | | | | | | | for acceptable values. Convergent validity was measured using the average variance extracted (AVE). The AVE values for each subscale should be at least 0.5 to establish convergent validity. The model's reliability was measured using the McDonald's omega values using a lower threshold of 0.7. Descriptive statistics for continuous variables were presented using mean and standard deviation, while categorical variables were summarized using frequencies and percentages. Our dependent variables were the likelihood of intention and sustenance All data were analyzed using SAS version 9.4 (SAS Institute Inc., Copyright © 2016 SAS Institute Inc. | considered gambling itself. This may have affected the results by increasing the likelihood of those engaged in gambling participating more. We used this measure to increase participation in our survey. Sixthly, we used only one questionnaire in assessing the explanatory potential of MTM. Lastly, it is important to note that due to the cross-sectional design of the study, causal relationships could not be established. |
| Efficacy testing of the SAVOR (Sisters Adding Fruits and Vegetables for Optimal Results) intervention among African American women: a | LaVonne Brown. (2020). Health Promot Perspect | America | In this study, a fourth-generation multi-theory model (MTM) of health behavior change was used to design and evaluate a Sisters Adding Fruits and Vegetables for Optimal Results (SAVOR) intervention for AA women. | 54 | Sisters Adding Fruits and Vegetables for Optimal Results. | A randomized controlled trial | The study utilized a randomized controlled trial (RCT) with measurements taken at pretest, posttest (after the three-week intervention) and follow-up (at the end of eight weeks). SAVOR (n=26) was compared to an equivalent knowledge-based intervention (n=28). Process evaluation was done for program fidelity and satisfaction. A validated 38-item self-reported | All data were analyzed using SPSS, version 25.0. Descriptive statistics for demographic and study variables at pretest, posttest, and follow-up were computed in the form of frequencies and percentages for categorical variables and means and standard deviations for metric variables. Differences between demographic variables and study variables between experimental and comparison groups at pretest were analyzed using the chi-square test for categorical variables and two-tailed F-test for metric variables. Mean differences in scores for study variables between pretest, posttest and follow up for experimental and comparison were | First, the data were collected through self-reports which have the potential for several biases such as acquiescence bias recall bias, dishonesty, exaggeration, etc. which may skew the results. However, this is the only method for collecting data about attitudes or the constructs of MTM. For recording the behavior observations could have been used but were not feasible due to constraints of resources. Future studies can utilize observations. Second, while the sample size was enough for an ORBIT IIb trial, a larger sample could enhance the power and ability to conduct subgroup analyses. Third, an efficacy trial has the potential |

| Article Title | Author, year, journal | Country | Research purpose | Participants | Behavior type | Research type | Research method | Data analysis | Research limitation |
|---|---|---|---|---|---|---|---|---|---|
| randomized controlled trial | | | | | | | questionnaire was used to measure changes in MTM constructs and past 24-hour consumption of fruits and vegetables. | compared using repeated-measures ANOVA test. Sphericity assumed within-subject effects tested using the Mauchly's test were reported. The significance level (α) was set at $P < 0.05$. For the initiation model that tested the intention for starting fruits and vegetable consumption, since participatory dialogue showed significant difference at pretest between experimental and comparison group, repeated measures analysis of covariance (ANCOVA) was applied with it being the covariate. | to sometimes overestimate the intervention's effect size when implemented for practice in a clinical setting, which was a limitation. Finally, a per-protocol analysis method for data analysis was used as opposed to intention-to-treat analysis which could introduce bias due to compromised randomization. However, the attrition in the trial was very small so the effect would have been minimal and is justified for pragmatic trials. |
| Effect of the fourth generation multi-theory model intervention on the quality of life in Iranian postmenopausal women: A randomized controlled trial | Nooshin Yoshany. (2021). Post Reproductive Health | Iran | This study was conducted to determine the effect of specific educational interventions on the quality of life among postmenopausal women. | 80 | Education interventions in the quality of life of postmenopausal women in Iran. | A randomized control trial | This randomized controlled trial was conducted on 80 menopausal women who met the inclusion and exclusion criteria and were selected through the multi-stage stratified random sampling method. The participants were randomly allocated to either the control or intervention group (40 per group). The intervention comprised 5 × 45-min educational sessions based on the Multi-Theory Model on the predetermined days of the week. The scores of the quality of life level were collected at | The collected data were analyzed using SPSS software package (version 22). In addition to descriptive statistics indices (central and dispersion indices), parametric or nonparametric analytical statistics tests were utilized after examining the normality or abnormality of the data by the Smirnov–Kolmogorov test. Furthermore, the independent test was used to compare demographic variables in the control and intervention groups. Moreover, analysis of variance on repeated measures and Bonferroni's pairwise comparison test was employed to investigate the changes in the scores of quality of life at pretest, posttest, and follow-up in both intervention and control groups. An analysis of covariance was also used to compare the mean scores of quality of life change in the intervention and control groups, | / |

| Article Title | Author, year, journal | Country | Research purpose | Participants | Behavior type | Research type | Research method | Data analysis | Research limitation |
|---|---|---|---|---|---|---|---|---|---|
| | | | | | | | baseline, immediately, and 3 months after the intervention using the Menopause-Specific Quality of Life questionnaire (MENQOL). The control group received a health advice. | as well as compare the intervention in the two groups with the pretest results. | |
| Effect of Health Promotion Interventions on Small Portion Size Consumption Behavior among College Students | Atul Gupta. (2023).INDIAN JOURNAL OF PUBLIC HEALTH | India | The study was done to see the effect of health promotion intervention on small portion size consumption behavior using multitheory model (MTM). | 150 | Health promotion intervention on small portion size consumption behavior. | A quasi-experimental study | A quasi-experimental study was conducted among students of age groups 18 - 21 years in two different colleges from North India between 2019 to 2020. About 150 participants in the intervention group as well as control group were selected and health promotion intervention in the form of motivational group counseling, one-to-one counseling, Power Point presentations, lectures, and messages were given to participants in intervention group. Difference in difference of proportions for meal consumption behavior and the difference in the difference of means | Statistical Package for the Social Sciences (SPSS) Version 24.0 (manufactured by the SPSS Inc., 233 South Wacker Drive, 11th floor, Chicago, IL 60606-6412, US). Univariate analyses done for quantitative data were represented as numbers, percentages, and means ± standard deviation. Chi-square test was used to find the association between the outcome and dependent variables. The odds ratio was used to measure the strength of association. Difference in difference of proportions for meal consumption behavior and the difference in the difference of means for body mass index, waist–hip ratio and for constructs of MTM for portion size consumption behavior were calculated. Paired t-test was used to test the significance between the continuous variables before and after the intervention. The linear regression model was used to test the association between the variables.95% confidence intervals were used to represent the lower and upper bounds of values. A two-tailed P < 0.05 was | / |

| Article Title | Author, year, journal | Country | Research purpose | Participants | Behavior type | Research type | Research method | Data analysis | Research limitation |
|---|---|---|---|---|---|---|---|---|---|
| | | | | | | | for body mass index, waist–hip ratio and for constructs of MTM for portion size consumption behavior were calculated. Paired t-test was used to test the significance between the continuous variables. | considered statistically significant for all analyses. | |
| Effectiveness of Tobacco Cessation Counselling and Behavioural Changes using Multi theory Model | Vijay Kumar. (2021).Indian Journal of Dental Research | India | The aim of the present study was to evaluate the effectiveness of tobacco cessation counselling and behavioural changes using Multi Theory Model (MTM). | 100 | Tobacco Cessation Counselling and Behavioural Changes. | A Follow-Up Study | A 28 item questionnaire multi-theory model (MTM) for health behaviour was administered at baseline, 2 weeks, 6 weeks and 12 weeks after providing standardized tobacco cessation counselling (TCC) intervention at baseline. | Dropout analysis with hot-deck imputation of data was done for participants with loss of follow up. Repeated measure ANOVA used to find the mean score difference of behaviour changes in from baseline to 12 weeks. Multiple comparison was done with Bonferroni adjustment. Scores per question were calculated for each construct of the behaviour initiation and behaviour sustenance. Independent t test was used to find mean difference between initiation and sustenance. Level of addiction was determined by Fagerstrom scores, which was divided into <6-low level of addiction and ≥6- high level of addiction. Two way ANOVA used to find out significant difference in level of addiction and behaviour change among participants. A $p < 0.05$ was considered as significant. | Though dropouts were present in the subsequent follow ups, Imputation analysis was done to minimize the bias due to dropouts. Non-randomised trial without control was one of the important limitation of the study. Inherent biases of questionnaire study was present. Randomized control trials will be needed to evaluate the effectiveness of MTM model as guiding principle in tobacco cessation. |
| Effects of an educational intervention based on the | Mohammad Ali Morowatisharifabad. | Iran | The purpose of this study was to design an educational program based on the multi-theory model (MTM) to deal with | / | Improving the quality of life among postmenopausal | A protocol | In designing this study, four phases are considered. In the first phase, the questionnaire of menopausal symptom acceptance behaviors will | We will apply the $\chi^2$ test, independent t-test, and paired t-test to analyze our data using SPSS version 21. | / |

| Article Title | Author, year, journal | Country | Research purpose | Participants | Behavior type | Research type | Research method | Data analysis | Research limitation |
|---|---|---|---|---|---|---|---|---|---|
| multi-theory model on promoting the quality of life in postmenopausal women: A protocol | (2020). INTERNATIONAL JOURNAL OF ENVIRONMENTAL RESEARCH AND PUBLIC HEALTH | | complications of the menopausal period and improve the women's quality of life. | | women. | | be designed based on the MTM using literature review and a panel of experts' viewpoints. The validity and reliability of the questionnaire will be confirmed at this stage. In the second phase, a descriptive study will be conducted by administering the questionnaire designed in the first phase along with the Menopause-Specific Quality of Life questionnaire. The third phase includes the curriculum design based on the findings of the descriptive study, investigations of various studies, and viewpoints of the experts" panel. Therefore, the main components of the intervention will be identified. These com-l ponents will determine the influential constructs of the MTM according to the descriptive research. Later, the related interventions and messages will be produced and designed from | | |

| Article Title | Author, year, journal | Country | Research purpose | Participants | Behavior type | Research type | Research method | Data analysis | Research limitation |
|---|---|---|---|---|---|---|---|---|---|
| | | | | | | | different sources. Intervention strate-l will include group discussion, lectures, confidence-building skills, movie screenings, role play, preparation of daily activities booklets for postmenopausal women, and training classes for husbands and children to improve social support for women. The interventions, contents, and messages designed with the presence of health professionals and members of the target community will be pre-tested by examining factors such as audience perception of the message, appropriateness of the education to the audience's literacy and culture, as well asl attractiveness, credibility, and acceptance of the materials. Finally, the fourth phase will be the implementation of the pre-test/post-test educational | | |

| Article Title | Author, year, journal | Country | Research purpose | Participants | Behavior type | Research type | Research method | Data analysis | Research limitation |
|---|---|---|---|---|---|---|---|---|---|
| | | | | | | | intervention using the intervention and control groups. | | |
| Effects of Multi-Theory Model based Behavior Change Intervention with Staircase Approach on Sedentary Lifestyle among Community-dwelling Older Adults: a Randomized Controlled Trial | Subinuer Tuerdi. (2024). research square | China | An experimental study will be conducted to verify whether a sedentary lifestyle modification intervention based on the MTM and the staircase approach is more effective than conventional education in reducing sedentary time and improving physical activity levels, functional capacity, and quality of life among community-dwelling older people. | 56 | Sedentary Lifestyle. | A Randomized Controlled Trial | This community-based, parallel-arm, assessor-blinded randomized controlled trial aims to estimate the effect of Multi-Theory Model based behavior change intervention with staircase approach on sedentary lifestyle among community-dwelling older adults. A total of 56 participants will be enrolled in this study and randomly assigned to the intervention group (participants will receive multi-theory model-based stepped sedentary lifestyle change intervention) and the control group (participants will receive conventional behavioral change advice). Endpoints will be collected at baseline (T1), immediately after the end of intervention (T2), week 12 (T3) and week 18 (T4). The primary endpoint is the change in | Data analyses follow intention-to-treat principles, using generalized estimating equation (GEE) models to assess differential changes in the outcome variables between the two groups from baseline to 12 weeks (group*time interaction), with adjustment for potential covariates as appropriate (i.e., baseline group differences at 2-sided P < 0.25). Effect sizes for continuous outcome variables were estimated using Cohen's d statistic based on between group mean differences from baseline T1 to T2, T3, and T4 endpoints with cutoffs set at 0.2 (small), 0.5 (medium), and 0.8 (large). Each GEE model includes the main effects of group (intervention vs control), time (T2, T3, T4 vs. baseline), and two-way interaction effects (group x time). The group difference in the change from baseline to 6 weeks, 12 weeks, or 18 weeks between the two groups is verified when the two-way interaction effects (group x time) was statistically significant. All statistical analyses are two sided and performed using IBM SPSS 25.0 software. values < 0.05 is considered statistically significant. | / |

| Article Title | Author, year, journal | Country | Research purpose | Participants | Behavior type | Research type | Research method | Data analysis | Research limitation |
|---|---|---|---|---|---|---|---|---|---|
| | | | | | | | self-reported sedentary time (min/day). Secondary endpoints include the changes in different domain sedentary time, sedentary behavior characteristics including the longest continuous sedentary time and the prevalence of prolonged sedentary bouts(≥30min), step count, time spent in light-intensity physical activity and moderate-to-vigorous physical activity, MCPAQ score, SPPB score, anthropometric parameters, blood pressure, SF-36score, and adverse events. | | |
| The effect of empowerment program to reduce Sugar Consumption based on the Multi-Theory Model on Body | Hamid Joveini, Masoumeh Hashemian.( 2022). BMC Womens Health | Iran | This study was conducted to determine the effect of empowerment program to reduce sugar consumption based on the MultiTheory Model (MTM) on Body Mass Index (BMI) and abdominal obesity in women aged 30–60 in Joven. | 400 | Iranian women's Body Mass Index and abdominal obesity empowerment program on reducing sugar consumption. | A Randomized Controlled Trial | This quasi-experimental study was conducted on the Joven city, Khorasan Razavi province, Iran country from October 2020 to August 2021. Sampling was performed as a multi-stage cluster. First, a descriptive study was performed among 400 women, and | At first, we checked the normality of the variables with the Kolmogorov Smirnov test. Independent t-test was used to examine the difference between the mean of quantitative demographic variables and the main research variables in the intervention and control groups. Chi-square test was used to check the difference in the ratio of qualitative variables in the intervention and control groups. Finally, we used the Generalized Estimating Equations (GEE) test to check the | / |

| Article Title | Author, year, journal | Country | Research purpose | Participants | Behavior type | Research type | Research method | Data analysis | Research limitation |
|---|---|---|---|---|---|---|---|---|---|
| Mass Index and abdominal obesity in Iranian women | | | | | | | then 128 people who were eligible to enter the interventional phase of the study were selected. In the control group, 63 people and in the intervention group, 65 people were eligible to enter the study. The educational intervention was performed in five 60-minute sessions for groups of 12 people. The instruments included the demographic questionnaire, sugar consumption checklist and researcher-made questionnaire based on MTM constructs. Before the intervention, one, three and six months after the intervention, the questionnaire was completed by both intervention and control groups also measurement of waist circumference and BMI were performed using standard instruments. | mean difference of the main variables over time (Before the intervention(t1), 1 month after the intervention(t2), 3 months after the intervention(t3) and 6 months after the intervention(t4) by group (intervention and control). Then, multivariable linear regression by ENTER technique was also used to investigate the association between the start and maintain behavior with MTM constructs. The data analysis was done with SPSS 17 and the level of confidence in all the tests was considered to be 95%. | |
| The evaluation | Traci Hayes. | America | The purpose of this study was to | 48 | Intervention to | A | The randomized controlled trial | All data were analyzed using IBM SPSS version 25. | An efficacy study has the potential to overestimate the |

| Article Title | Author, year, journal | Country | Research purpose | Participants | Behavior type | Research type | Research method | Data analysis | Research limitation |
|---|---|---|---|---|---|---|---|---|---|
| of a fourth-generation multi-theory model (MTM) based intervention to initiate and sustain physical activity | (2019).Health Promot Perspect | | determine the efficacy of an intervention based on the fourth generation, multi-theory model (MTM) of health behavior change for initiating and sustaining physical activity among African American women when compared to a first generation, knowledge-based intervention. | | initiate and maintain physical activity. | randomized-controlled study | (RCT) utilized a pre-test, post-test and 6-week follow up evaluation with an experimental (n=25) group and a comparison group (n=23). Process evaluation for satisfaction and program fidelity was conducted along with impact evaluation for changes in MTM constructs, intent to initiate and sustain physical activity, minutes of physical activity, body mass index (BMI), waist circumference and blood pressure in hypertensives. | Descriptive statistics for sociodemographic variables (age, employment and income) and the MTM constructs. Chi square test compared the differences between categorical variables such as education and employment for the experimental and comparison groups. Two-tailed t test was be used to detect any baseline differences in age between the groups. Means scores over 3-time points for the same individuals were measured using repeated measures analysis of variance (ANOVA) test. Sphericity assumed within subject effects tested using the Mauchly's test for sphericity were used. The significance level (α) was set at $P < 0.05$. Repeated measures ANOVA was conducted to discern statistically significant differences in the means (from before to after to 6-week follow-up in the interventions) for each theory construct, minutes of PA, and the anthropometric and clinical measures between the experimental (based on MTM) and comparison (knowledge-based) groups. Repeated measures analysis of covariance (ANCOVA) was applied for covariates of participatory dialogue and emotional transformation which were found to be significantly different between the 2 groups at pre-test. | intervention's effect when implemented for practice in a clinical setting. This limitation can be addressed by future replication and effectiveness studies. The study met the sample requirement of 20 participants in each group; however, the sample was relatively small. A larger sample would improve the study power. The sample consisted of only one racial group and one gender. The results may not be generalizable especially across different groups. Therefore , it should be implemented among various racial and ethnic groups as well as with men. Future replication studies and effectiveness trials will have to be undertaken. The study relied on self-reports of PA. The participants recalled and recorded the number of PA minutes from the previous week. Future studies can use objective measures such as the use of accelerometers in recording PA. Finally, the survey brevity may have limited its ability to probe the constructs in their entirety. |
| Water Pipe Smoking | Saeed Bashirian , | Iran | The aim of this study was to determine of efficacy of an | 94 | Teenage boys cut down on smoking. | A randomized controlled trial | Overall, 94 male adolescent students (grades 10, 11) smoked water pipe | Data were analyzed by SPSS software (ver. 22 (Chicago, IL, USA), paired sample t-test (Comparison of mean score of | / |

| Article Title | Author, year, journal | Country | Research purpose | Participants | Behavior type | Research type | Research method | Data analysis | Research limitation |
|---|---|---|---|---|---|---|---|---|---|
| Reduction in the Male Adolescent Students: An Educational Intervention Using Multi-Theory Model | Hamid Abasi.(2019 ).Journal of Research in Health Sciences | | educational intervention based on Multi-Theory Model (MTM) to reduce WPS in the male adolescent students in Iran. | | | | (WP) in the past month (current WP smokers) were selected, allocated randomly in two groups (47 students in intervention group and 47 students in control group), in two different schools in 2018 in Hamadan City, western Iran. Data were collected utilizing a valid and reliable questionnaire based on MTM constructs and demographic variables. Educational intervention was designed in five 45-min sessions. Two groups were followed-up three-months after completion of intervention. | structures in each group), independent-sample t-test (Comparison of the mean score of structures between groups), Chi-square (to compare qualitative variables), Friedman test (testing change in the frequency of WPS in groups) and in this study, P<0.05 was considered significant. | |
| Effects of dietary guidance based on MTM model on taste changes in patients with gastrointestinal | Guo, Q. (2022). Nursing practice and research | China | To investigate the effect of dietary guidance based on multi-theory model (MTM) on taste change in patients with gastrointestinal malignancies undergoing chemotherapy. | 125 | Changes of taste in patients with gastrointestinal malignancies undergoing chemotherapy | Original study, intervention study, control group | A total of 125 patients with gastrointestinal malignancies treated in the oncology department of a Grade A hospital in Chongqing from January to December 2021 were selected as the study objects. Among them, 1 patient dropped out (condition worsened), 2 patients lost | SPSS 23.0 statistical software was used for data processing, and the measurement data were expressed as "mean ± standard deviation". t test was used for comparison of mean between groups when variances were homogeneous, and t 'test was used for comparison of mean between groups when variances were uneven. P<0.05 was considered to be statistically significant. | / |

| Article Title | Author, year, journal | Country | Research purpose | Participants | Behavior type | Research type | Research method | Data analysis | Research limitation |
|---|---|---|---|---|---|---|---|---|---|
| malignancies undergoing chemotherapy | | | | | | | follow-up after discharge, and 122 patients with gastrointestinal malignancies were enrolled. According to the principle of comparability of basic data between groups, 59 cases were divided into control group and 63 cases were divided into observation group. The control group was guided by the conventional diet of oncology department, and the observation group was guided by the diet guidance strategy based on the MTM model. The Kano chemotherapy-related Taste Change Scale and Core Quality of Life Questionnaire (QLQ-C30) were used to evaluate the difference in taste change and quality of life between the two groups at the 8th week after the intervention. | | |
| Behavior change | Zhang Saisai.(2024 | China | To explore the effect of behavior change intervention based on | / | Fatigue behavior after stroke | A Cross-Sectional Study | The control group received routine nursing, and after admission, the | SPSS26.0 software was used to perform t test, $\chi^2$ test, rank sum test and repeated measurement analysis of variance. The | Behavior change intervention in post-stroke fatigue patients based on multiple theoretical models |

| Article Title | Author, year, journal | Country | Research purpose | Participants | Behavior type | Research type | Research method | Data analysis | Research limitation |
|---|---|---|---|---|---|---|---|---|---|
| intervention in post-stroke fatigue patients based on multiple theoretical models | ).Journal of Nursing Science | | multiple theoretical models on patients with post-stroke fatigue. | | | | responsible nurse provided health education such as disease-related knowledge, diet guidance, medication, rehabilitation and exercise knowledge to provide psychological support for patients. After discharge, patients were instructed to pay attention to the department's wechat public account, and wechat groups of patients and their families were established to send health education knowledge regularly. From hospital admission to 3 months after discharge, the responsible nurse will keep in touch with the patient by phone or wechat, guide the patient's lifestyle, and complete the follow-up of 1 month and 3 months after discharge. The experimental group received a behavior change program based on multiple theoretical models on the basis of routine care. | test level α=0.05. | |

| Article Title | Author, year, journal | Country | Research purpose | Participants | Behavior type | Research type | Research method | Data analysis | Research limitation |
|---|---|---|---|---|---|---|---|---|---|
| Impact of a mobile health intervention based on multi-theory model of health behavior change on self-management in patients with differentiated thyroid cancer: protocol for a randomized controlled trial | Yang Jiang.(2024) Frontiers in Public Health | China | In this study, we used a microblogging platform as an intervention vehicle and mobile patient-doctor interactive health education as a means of intervention, with the aim of improving the health behaviors of DTC patients as well as the corresponding clinical outcomes. | patients over 18 years of age with differentiated thyroid cancer who were given radioactive iodine-131 therapy as well as endocrine therapy after radical surgery for thyroid cancer | Self-management in patients with differentiated thyroid cancer | A randomized controlled trial | The intervention group will receive the MTM-mHealth model intervention and the control group will receive usual care. The intervention will last for 3months. Questionnaires and physical examinations will be conducted at baseline and at 3 and 6months of follow-up to check for changes in self-management behaviors and TSH control. | Statistical analysis was performed using SPSS Statistics 26.0 software. | Impact of a mobile health intervention based on multi-theory model of health behavior change on self-management in patients with differentiated thyroid cancer: protocol for a randomized controlled trial |
| Exploring the Influencing Factors of COVID-19 Vaccination Willingness among Young | Yue Su, Jia Xue. (2023) .International Journal of Environmental Research | China | This study aims to explore the influencing factors related to COVID-19 vaccine willingness among young adults in China. | / | COVID-19 vaccine willingness among young adults in China. | A semi-structured interviews | Using semi-structured interviews, this study explored the factors that would motivate young adults with vaccine hesitancy to get the COVID-19 vaccine. | Thematic analysis is a traditional qualitative method that is widely used in the health sciences. Based on the interview transcripts, we performed thematic analysis to code interview data manually. In this research, we followed the thematic analysis steps suggested by Braun and Clarke, which included familiarizing ourselves with our data generating initial codes, searching for themes, reviewing themes, | / |

| Article Title | Author, year, journal | Country | Research purpose | Participants | Behavior type | Research type | Research method | Data analysis | Research limitation |
|---|---|---|---|---|---|---|---|---|---|
| Adults in China | and Public Health | | | | | | | defining and naming themes, and producing the report. The analysis of transcribed interviews was guided by MTM framework. To enhance the rigor of the analysis, the initial code development and subsequent theme generation and definition were discussed between two researchers (the first author and the second author) until a final agreement was reached. To demonstrate the meanings of the themes more clearly, we selected concrete examples that participants expressed to characterize each theme. However, such qualitative studies are also considered to rely on the understandings of individuals. Moreover, when the time required for interviews is long and the corresponding transcripts are large, repeated reading and examination of transcripts and rounds of discussions among researchers can consume a great deal of cognitive resources. Considering the long periods of interviews in this study could increase the possibility of oversights occurrence, we used topic modeling to complement thematic analysis and provide insights from a novel perspective. | |
| Male students' experiences on predictors of waterpipe | Saeed Bashirian. (2019). Tobacco | Iran | This study aimed to explain the experiences of high school students in Iran on predictors of WPS reduction based on a | 34 | Iranian boys cut down on hookah smoking. | A qualitative study in Iran | This study was a qualitative study of directed content analysis that was conducted in high school male students in Hamadan, Iran, in 2017. | The methods of qualitative research, which is presented in 2005 by the Hsieh and Shannon analysis, is a directed content analysis method or theory based content analysis method21. In the directed content analysis method, initial | First, this study is qualitative and limited to grade 8–12 high school students in Hamadan. Being qualitative it provides a snapshot in time and does not generalize to all community adolescents. Second, our study was not implemented among |

| Article Title | Author, year, journal | Country | Research purpose | Participants | Behavior type | Research type | Research method | Data analysis | Research limitation |
|---|---|---|---|---|---|---|---|---|---|
| smoking reduction: A qualitative study in Iran | Prevention & Cessation | | multi-theory model (MTM) of health behaviour change. | | | | In this study,34 students who had smoked waterpipe (WP) in the last month were recruited through snowball sampling that was continued until data saturation. | coding begins with an established theory or results. This type of analysis aims to validate the theory or develop a conceptual framework. The theory chosen in this type of study can help in focusing the research question. On the other hand, theory can help in predicting interesting variables or relationships between variables. As a result, it is also useful in determining how the initial encodings occur and the relationships between the codes. | female students, because of challenges of getting permission to enter girls' high schools. Thus, implementation of the program to cover female students could give better estimates of WP use and associated factors among all Iranian adolescents. Third, the present study's design is limited in being able to draw a definitive conclusion regarding efficacy. |
| Establishment and validity test of occupational exposure risk perception scale for staff in disinfection supply center | Zhu Q. (2023). Journal of Nursing Science | China | The scale of occupational exposure risk perception for staff in disinfection supply center was developed and its reliability and validity were tested. | 16 | Occupational exposure risk for workers in disinfection supply centres | Literature analysis, qualitative interview | The structure of the scale was determined by the theory of risk perception and the multi-theory model of health behavior change. The item pool of the scale was compiled by literature analysis and qualitative interviews and the first draft of the scale was formed by Delphi expert consultation. The reliability and validity of the seale were tested by questionnaire survey. | SPSS2 3.0 software was used for statistical analysis. The counting data were described by frequency and component ratio, and the sampling data conforming to normal distribution were described by mean number W standard difference. | / |
| Exploring Yoga Behaviors among College | Chia-Liang Dai. (2023) .International | America | This paper studies the yoga behavior of college students based on a multi-theoretical model of health behavior | 79 | Yoga behavior of college students. | A qualitative research | Students were asked to submit the yoga journals by answering the following open-ended question: describe your experiences of yoga | Directed content analysis along the seven constructs of the multi-theory model (MTM) of health behavior change was utilized on the collected data. The constructs for initiation of yoga behavior in MTM are advantages, disadvantages, | First, the participants in the study were predominantly female (91%). This limits our understanding of yoga practice behavior for all genders. Additionally, data from 56% of the class were collected. While the aim of |

| Article Title | Author, year, journal | Country | Research purpose | Participants | Behavior type | Research type | Research method | Data analysis | Research limitation |
|---|---|---|---|---|---|---|---|---|---|
| Students Based on the Multi-Theory Model (MTM) of Health Behavior Change | Journal of Environmental Research and Public Health | | change. | | | | practice in this class. Students were informed that the submission of the yoga journals was voluntary and that the submission or not would not affect their grades. There was no specific requirement regarding the length or format of the paper submission. Students turned in their yoga journals to the Web Campus Learning Management Platform used by the university. A doctoral-level graduate assistant who had completed the qualitative research methodologies class and served as a research assistant in other research projects handled the raw data. | behavioral confidence, and changes in the physical environment, while the constructs in the sustenance model are emotional transformation, practice for change, and changes in the social environment. Two researchers completed the initial coding independently. The initial coding process was carried out by two researchers who were not part of the class, thus maintaining anonymity. The results of the initial coding were discussed and reconciled among the three researchers. The first coder's research centered on using lifestyle-based behavior approaches, especially yoga, to promote personal health and wellness. The second coder conducted intervention research targeting mental health and wellness issues. The third coder was an expert in the theory of health behavior change as well as complementary and alternative medicine. Three coders have been trained in conducting qualitative research, have published qualitative studies, and were familiar with the constructs of MTM of health behavior change. | qualitative research is not generalizability, and we did obtain data saturation, the results do point to limited transferability. Demographic data of the participants such as age and race/ethnicity were not collected which is another limitation of the current study. |
| Multi-Theory Model (MTM) and change in childbearing behavior: A | Masoumeh Abbasi Shavazi,Manoj Sharma.(202 | Iran | To explore the effectiveness and usefulness of multi-theoretical models in encouraging couples to improve their fertility. | / | childbearing behavior | A perspective | / | / | / |

| Article Title | Author, year, journal | Country | Research purpose | Participants | Behavior type | Research type | Research method | Data analysis | Research limitation |
|---|---|---|---|---|---|---|---|---|---|
| perspective | 4)Journal of Research Development in Nursing and Midwifery | | | | | | | | |
| A phenomenological qualitative study based on the multi-theory model (MTM) of health behavior change to identify factors and ways to design interventions for quitting gambling among older | Laurencia BONSU.(2024)Journal of Health and Social Sciences (JHSS) | America | This qualitative, phenomenological study examines gambling cessation behavior using the multi-theory model (MTM) of behavior change and its initiation and sustenance stages. Directed content analysis categorizes and interprets participants' narratives to reveal the complex relationships that affect gambling behavior. | Ten | Smoking behavior of the elderly | A phenomenological qualitative study | Ten participants were engaged in in-depth interviews to gain comprehensive insights into their gambling experiences until data saturation occurred. | The study utilized directed content analysis, guided by the MTM, to discern patterns, motivations, and factors influencing gambling behavior. | This qualitative phenomenological research focuses only on older adults in a metropolitan area. Due to its qualitative nature, the study only offers a limited understanding of a specific moment and cannot be generalized to all older adults in all communities. Another limitation is that the participants are predominantly males, which limited our understanding of gambling quitting behavior in females. Furthermore, the current study's methodology is constrained in its ability to definitively establish effectiveness due to its phenomenological nature. Though the study has been presented within the framework of COnsolidated criteria for REporting Qualitative research (COREQ, Additional File 1,data triangulation has not been applied to test the validity of the convergence of data from other sources. |

| Article Title | Author, year, journal | Country | Research purpose | Participants | Behavior type | Research type | Research method | Data analysis | Research limitation |
|---|---|---|---|---|---|---|---|---|---|
| adults | | | | | | | | | |
