## Supplemental Table 3 for "Application of multi-theory model(MTM)health behavior change: A scoping review": S3_File.pdf

### File 2. Information for intervention articles.

| Article Title | Intervention technique | Intervention duration |
|---|---|---|
| Efficacy testing of the SAVOR (Sisters Adding Fruits and Vegetables for Optimal Results) intervention among African American women: a randomized controlled trial | The experimental arm received the MTM-based three-week intervention while the comparison arm received an equivalent knowledge-based intervention delivered didactically. A knowledge-based intervention was chosen due its traditional and practical method to deliver pathology of fruit and vegetable consumption and related data associated with its effects on the human body. | Eleven weeks. |
| Effect of the fourth generation multi-theory model intervention on the quality of life in Iranian postmenopausal women: A randomized controlled trial | The intervention group received the educational program based on the MTM, whereas the control group received only the routine care. Based on the values obtained from the descriptive-analytical and pilot phase of the intervention, the first type error of 5%, test power of 80%, the sample size was calculated to be 40 cases per group. | Three months. |
| Effect of Health Promotion Interventions on Small Portion Size Consumption Behavior among College Students | About 150 participants in the intervention group as well as control group were selected and health promotion intervention in the form of motivational group counseling, one- to- one counseling, Power Point presentations, lectures, and messages were given to participants in intervention group. Difference in difference of proportions for meal consumption behavior and the difference in the difference of means for body mass index, waist–hip ratio and for constructs of MTM for portion size consumption behavior were calculated. Paired t- test was used to test the significance between the continuous variables. Differential interventions for each group were planned for 6 months and divided into two phases. For the initial 2 months, intervention in the form of motivational group counseling, one- to- one counseling, classroom trainings in the form of PowerPoint presentation, and lectures were given to all the participants. In the next 4 months, health promotion messages were sent individually to all participants of the intervention group via WhatsApp, after every 15- day interval. | Six months. |
| Effectiveness of Tobacco Cessation Counselling and Behavioural Changes using Multi theory Model | Each participant was given standardised 10- minute intervention with power point presentation and face to face interview. Doubt clearing sessions for participants was done separately on the same day. Therapeutic measures like oral prophylaxis, restorations and referral for speciality treatment were administered as needed. Multi theory model, a 28 items questionnaire [Supplement 1] with 6 point Likert scale was used for assessment of change in behaviour at baseline before intervention. The participants were recalled at 2 weeks. 6 weeks and 12 weeks and assessment of their tobacco habit and change in behaviour were done through same questionnaire followed by brief counselling on tobacco cessation. | Six months. |
| Effects of an educational intervention based on the multi-theory model on promoting the quality of life in postmenopausal women: A protocol | The method of instruction will be selected according to the content type and audience accessibility, and will include group discussion, lectures, confidence-building skills, movie screenings, role play, preparation of daily activities booklets for postmenopausal women, and training classes for husbands and children. The research team will finalize these educational methods according to their priorities. | / |
| Effects of Multi-Theory Model based Behavior Change Intervention with Staircase Approach on Sedentary Lifestyle among Community-dwelling Older Adults: a | This community-based, parallel-arm, assessor-blinded randomized controlled trial aims to estimate the effect of Multi-Theory Model based behavior change intervention with staircase approach on sedentary lifestyle among community-dwelling older adults. A total of 56 participants will be enrolled in this study and randomly assigned to the intervention group (participants will receive multi-theory model-based stepped sedentary lifestyle change intervention) and the control group (participants will receive conventional behavioral change advice). Endpoints will be collected at baseline (T1), immediately after the end of intervention (T2), week 12 | 6-week intervention + 12-week follow-up. |

| Article Title | Intervention technique | Intervention duration |
|---|---|---|
| Randomized Controlled Trial | (T3) and week 1 (T4). The primary endpoint is the change in self-reported sedentary time (min/day). Secondary endpoints include the changes in different domain sedentary time, sedentary behavior characteristics including the longest continuous sedentary time and the prevalence of prolonged sedentary bouts(≥30min), step count, time spent in light-intensity physical activity and moderate-to-vigorous physical activity, MCPAQ score, SPPB score, anthropometric parameters, blood pressure, SF-36score, and adverse events. | |
| The effect of empowerment program to reduce Sugar Consumption based on the Multi-Theory Model on Body Mass Index and abdominal obesity in Iranian women | Before the intervention, one, three and six months after the intervention, the questionnaire was completed by both intervention and control groups also measurement of waist circumference and BMI were performed using standard instruments. The obtained data were analyzed by SPSS 17. | Ten months. |
| The evaluation of a fourth-generation multi-theory model (MTM) based intervention to initiate and sustain physical activity | The study orientation and enrollment along with the interventional sessions and 6-week follow-up were conducted at a recreational center on the campus of a midsize Historically Black University in the Southern United States. The participants in the experimental arm (n=25) engaged in activities based on the fourth generation MTM constructs to influence behavior adoption and behavior sustenance. The MTM-based study participants attended three 60-minute sessions over a 3-week period. The MTM intervention was highly interactive, consisting of bi-directional conversations about the advantages and disadvantages of PA, participating in demonstrations of moderate-impact exercises and movements, 10 to 15 minutes workout sessions, instruction on gym equipment, affective learning activities, participation with regard to self-monitoring of the behavior and mobilization of social support. The study participants in the comparison arm received standard first-generation knowledge-based instruction (n=23). The comparison intervention relied on a didactic approach consisting of lectures and Power Point presentations. The experimental and comparison sessions were monitored by an independent observer who verified content and recorded class times to ensure compliance with the study protocol. Participants completed a satisfaction survey during the follow-up session. | Five months. |
| Water Pipe Smoking Reduction in the Male Adolescent Students: An Educational Intervention Using Multi-Theory Model | Designed educational intervention according to the analysis of pre-test results was implemented for the experiment group in five training sessions 45 min for 1 week. All of educational sessions were held at the school amphitheater. Pamphlets and a booklet were given to the students after each educational session. First session was focus group discussion that showed photo clips. The focus group discussions were conducted in 10-member groups about advantages and disadvantages of reducing WPS. Second session was focused on students' behavioral confidence toward reducing WPS. Third session was focused on emotional transformation that influenced students' positive and directing emotions to reduce WPS through showing of video clips. The fourth session was held with the aim of increasing students' self-monitoring and setting goal of reducing WPS by keeping diary. The fifth session focused on changing the social environment. Moreover, students' posters designed by health educator about harms effects of WPS, installed in main halls at the school. Parallel to these educational sessions, we created a social network on Telegram and added all of students in intervention group to this channel. This channel included image file of pamphlet, booklet and poster that we designed for them. Control group received a pamphlet about healthy nutrition during adolescence period. Finally, three months after educational intervention, the questionnaire was completed by both groups again. | More than three months. |
| Effects of dietary guidance based on MTM model on taste changes in patients with gastrointestinal malignancies undergoing chemotherapy | The control group was guided by the conventional diet of oncology department, and the observation group was guided by the diet guidance strategy based on the MTM model. The Kano chemotherapy-related Taste Change Scale (CiTAS) and Core Quality of Life Questionnaire (QLQ-C30) were used to evaluate the difference in taste change and quality of life between the two groups at the 8th week after the intervention. The control group was guided by the routine diet of oncology department. Provide dietary guidance to eligible patients during perichemotherapy, including explaining CITA-related knowledge to patients and their caregivers before | / |

| Article Title | Intervention technique | Intervention duration |
| --- | --- | --- |
| | chemotherapy, including the cause, clinical manifestations, duration and coping methods of CITA; During chemotherapy, targeted dietary guidance was given according to patients' taste changes; After the end of chemotherapy, the patient was provided with discharge guidance, and the changes of taste and nutritional intake were continuously followed up. Observation group based on the control group, MTM was used as the theoretical basis to formulate dietary guidance for patients with gastrointestinal malignancies undergoing chemotherapy. | |
| Behavior change intervention in post-stroke fatigue patients based on multiple theoretical models | The control group received routine nursing, and after admission, the responsible nurse provided health education such as disease-related knowledge, diet guidance, medication, rehabilitation and exercise knowledge to provide psychological support for patients. After discharge, patients were instructed to pay attention to the department's wechat public account, and wechat groups of patients and their families were established to send health education knowledge regularly. From hospital admission to 3 months after discharge, the responsible nurse will keep in touch with the patient by phone or wechat, guide the patient's lifestyle, and complete the follow-up of 1 month and 3 months after discharge. The experimental group received a behavior change program based on multiple theoretical models on the basis of routine care. | More than three months |
| Impact of a mobile health intervention based on multi-theory model of health behavior change on self-management in patients with differentiated thyroid cancer: protocol for a randomized controlled trial | The intervention group will receive the MTM-mHealth model intervention and the control group will receive usual care. The intervention will last for 3months. Questionnaires and physical examinations will be conducted at baseline and at 3 and 6months of follow-up to check for changes in self-management behaviors and TSH control. | from March 2023 to March 2024 |
